# Beyond Kidney Volume: 3D Mapping of Parenchymal, Cortical and Medullary Thickness with Disease, Anthropometry and Kidney Adiposity

**DOI:** 10.64898/2026.10.19.26365903

**Authors:** Marjola Thanaj, Nicolas Basty, Brandon Whitcher, Marili Niglas, Camilo Bradford-Bell, Ramprakash Srinivasan, Anna S Li, Rachel Lennon, Madeleine Cule, Maciej Tomaszewski, Jimmy D Bell, E Louise Thomas

## Abstract

**Introduction:** Kidney volume is associated with renal function, but conventional whole-organ measurements may miss regional changes in kidney substructures. We used magnetic resonance imaging (MRI) to test whether kidney substructure volumes and three-dimensional (3D) thickness phenotypes are associated with disease, adiposity, and incident outcomes.

**Methods:** Automated abdominal MRI segmentations from 70,270 UK Biobank participants were used to quantify left and right kidney cortex, medulla, and parenchyma volumes and mesh-based thickness maps. Mass univariate regression generated statistical parametric maps linking regional thickness to anthropometric traits and disease (CKD, type-2 diabetes [T2D], hypertension). Bidirectional causal mediation analyses were performed using kidney substructure volumetric, thickness PCs and kidney adiposity. Longitudinal changes were assessed in 2,610 participants with repeat imaging. Cox proportional hazards models tested the associations between kidney volumes and principal component (PC) scores of thickness and incident CKD, T2D, hypertension, cardiovascular disease (CVD), and mortality.

**Results:** CKD was associated with thinning of the parenchyma and most pronounced in the medulla (87.7% left, 72.8% right), whereas associations in the cortex were mixed. In contrast, T2D and hypertension were predominantly associated with increased thickness, most extensively in the cortex (T2D: 84.39% left, 96.35% right; hypertension: 94.8% left, 99.9% right). Age-disease interactions were associated with reduced thickness across all conditions. Smaller medulla volume and lower cortical thickness mediated the association between CKD and renal sinus fat. Kidney substructure volume was inversely associated with incident CKD across all substructures and both kidney sides (p<0.05), but positively associated with incident hypertension and mortality (p<0.05). Thickness PC scores also predicted outcomes independently of volume, with PC3 in the left cortex (HR 0.91 [0.84– 0.98]), reflecting inferior pole/hilar thickness, associated with incident CKD.

**Conclusions:** MRI-derived kidney substructure volumes and 3D thickness maps provide complementary, spatially-resolved markers of kidney morphology, that are mechanistically linked to renal adiposity and future disease risk.

**Translational Statement / Lay Summary:** This study used MRI scans from the UK Biobank to examine kidney structure in much greater detail than is possible with standard measurements of kidney size or volume. Instead of treating the kidney as a single organ, we measured tissue thickness at thousands of locations across its three main layers: the cortex (outer layer), medulla (inner layer), and parenchyma (the main functional tissue). Thinning of these regions can indicate tissue loss, while thickening may reflect swelling or other structural changes. We found that ageing, body composition, fat around and within the kidney, and conditions such as chronic kidney disease, type 2 diabetes, and hypertension were each associated with distinct patterns of structural change thickening across different parts of the kidney. These detailed 3D thickness maps reveal changes that are not captured by simple measurements of kidney size and could help identify early signs of kidney disease before more obvious damage occurs

## Introduction

Chronic kidney disease (CKD), type 2 diabetes (T2D), and hypertension are major causes of global morbidity and mortality, and each can alter kidney structure before, during, or after measurable functional decline^1,2^. Clinical assessment typically relies on estimated glomerular filtration rate (eGFR), albuminuria, diagnostic codes, and blood pressure, while large imaging studies use global measures such as kidney length, total kidney volume, or cortical thickness^3,4^. In the UK Biobank, body-surface-area-adjusted kidney volume is associated with eGFR, muscle volume, and incident CKD risk, and left-right volume asymmetry with higher CKD risk^5^, but a single summary number may obscure local change across the cortex, medulla, parenchyma, and perirenal or sinus regions.

The cortex and medulla have distinct roles: the cortex contains most glomeruli and proximal tubules and is central to filtration, whereas the medulla concentrates urine and maintains osmotic gradients, and ageing, nephron loss, diabetes, hypertension, and obesity may affect them differently^4,6^. Cortical volume declines from middle age, while medullary volume is comparatively preserved^3,4^, and larger cortical volume is linked to elevated glomerular pressure independent of nephron number^7^. CKD is associated with reduced kidney size and cortical thinning, while early diabetes and obesity are associated with renal hypertrophy and hyperfiltration^8,9^. Structural change is therefore unlikely to be uniform across the kidney or disease stage, and local thickness variation may provide more informative phenotypes than whole-organ volume alone.

Large-scale MRI and automated segmentation make spatially detailed kidney phenotyping feasible at population scale^10^. Statistical parametric mapping and mass univariate regression (MUR), long used in neuroimaging, cardiovascular and abdominal imaging, map associations across aligned 3D surfaces while controlling for multiple testing^11–16^. These vertex-wise techniques have rarely been applied to kidney substructures, despite well-documented cortical and medullary changes in CKD, diabetes, and hypertension^3^.

Perirenal and renal sinus fat sit close to the kidney and may reflect local metabolic, vascular, or mechanical stress distinct from BMI or waist-to-hip ratio. Perirenal fat thickness has been linked to reduced GFR^17^, and renal sinus fat to intrarenal haemodynamic abnormalities independent of visceral fat^18^. Their relationship with local kidney thickness across CKD, T2D, and hypertension remains uncharacterised at population scale.

Building on our earlier surface-to-surface analysis of whole-kidney shape in the UK Biobank^15^, this study analyses the cortex and medulla separately and bilaterally, as they are functionally distinct and the kidney is anatomically asymmetric. Our exploratory aims were to (i) map associations between substructure thickness/volume and anthropometric traits, CKD, T2D, and hypertension, and (ii) quantify longitudinal change using repeat imaging. Our hypothesis-driven aims were to (iii) investigate interrelationships between kidney phenotypes, disease, and kidney adiposity, and (iv) test whether substructure volumes and thickness-derived principal components (PCs) were independently associated with incident CKD, T2D, hypertension, cardiovascular disease (CVD), and all-cause mortality.

## Methods

### Data

Full details regarding the UK Biobank abdominal MRI protocol have previously been reported^19^. Briefly, we used neck-to-knee Dixon MRI acquisitions comprising six overlapping series. Data were obtained under UK Biobank application number 44584, with ethical approval (REC 11/NW/0382) and informed consent from all participants. Further information: www.ukbiobank.ac.uk.

### Disease Definition

CKD was defined as eGFR below 60 ml/min/1.73 m² and a diagnosis or procedure code for chronic kidney disease, dialysis, or kidney transplantation. T2D was defined using diagnosis and self-report codes. Hypertension was defined as antihypertensive medication use, mean blood pressure >=140/90 mmHg at the imaging visit, or prior diagnosis. Incident outcomes after imaging were defined from hospital, primary care, self-report, death and first-occurrence records, with censoring at first event, death, or last follow-up (Supplementary Table S1).

### Image processing

Kidney image-derived phenotypes (IDPs) were obtained automatically from the neck-to-knee 2-point Dixon MRI (details in Supplementary Material). A 3D U-Net pre-trained to jointly segment 28 organs/tissues was fine-tuned per kidney substructure using soft Dice loss, with manual annotations from 100 participants representative of the UK Biobank population for the cortex, medulla, and vessels. Segmentation quality was confirmed by visual inspection by experienced analysts.

### Estimates of Perirenal and Renal Sinus Fat

We quantified perirenal and renal sinus fat from abdominal MRI. Fat maps were derived from the fat fraction (fat/fat+water) channels, with voxels binarised at 0.7 to define fat-rich tissue. Visceral adipose tissue (VAT) segmentations from our previously published automated pipeline^10^, were used as an anatomical constraint to isolate perirenal fat compartments. Binary closure was applied to the kidney masks to fill potential holes. The mask was dilated by five voxels and then a renal shell was defined by subtracting the original mask from the dilated mask. Renal sinus fat was fat within the closed kidney mask while perirenal fat was fat in the shell after subtracting renal sinus fat. The perirenal fat ratio was perirenal fat volume divided by renal shell volume.

### Image Registration and Mesh Construction

Preprocessing ensured that all kidney substructures were spatially contained within the kidney (see Supplementary Material). Organ templates were constructed following our previously published procedure^14^. For the cortex and parenchyma, template surfaces were built by averaging segmentations from 20 participants selected to span the cohort’s anthropometric range, whereas for the medulla, a single-subject template was used, whose thinner, more variable boundaries are more sensitive to averaging artefacts. Templates were registered to each participant via rigid, affine, and non-rigid registration (Image Registration Toolkit, IRTK^14^), yielding surface meshes with ∼4,000 anatomically consistent vertices per participant. At each vertex, cortical thickness was defined as the local depth of the cortical rim, measured as the the Euclidean distance from the outer cortex surface to the cortico-medullary boundary, medullary thickness as the depth of the medulla from its outer surface to the hilar vessel/sinus surface, and parenchymal thickness as the full depth of renal tissue from the capsule to the central vessel/sinus surface (Supplementary Material, Supplementary Figure S1).

### Quality Control

This study included 84,405 participants at initial imaging and 3,084 with a follow-up scan ∼2.3 years later. We removed participants with missing clinical, anthropometric, or biological data (n=9,966) and those with cortical volume <50 ml or medullary volume <15 ml (n=1,085), leaving 70,270 participants at baseline. For follow-up, 474 datasets were excluded for missing anthropometric variables, leaving 2,610 participants (Supplementary Figure S2).

### Mass Univariate Regression Analysis

Associations between vertex-wise kidney thickness and covariates were assessed using mass univariate regression analysis^20^. We applied threshold-free cluster enhancement (TFCE)^21^ and permutation testing (100 permutations) to derive the p-values associated with each regression coefficient following adjustment for relevant covariates with the correction for multiple comparisons via the Benjamini-Hochberg false discovery rate (FDR) procedure^22^ (Supplementary Material).

To determine which factors were associated with substructure thickness, we fitted a linear regression model at each vertex with age, sex, ethnicity, BMI, waist-to-hip ratio (WHR), alcohol and smoking status, dominant hand grip strength (HGS), fluid intake, nonsteroidal anti-inflammatory drugs (NSAIDs) taken for pain, Townsend deprivation index, urinary sodium and urea, aspartate aminotransferase:alanine aminotransferase (AST:ALT) ratio, mid-thigh intermuscular adipose tissue (IMAT)^23^ and CKD, T2D, and hypertension as predictors, with FDR correction (Supplementary Material), adjusting also for time since the initial assessment visit (9.9 ± 2.4 years before imaging). Age×disease and BMI/WHR×T2D interactions assessed accelerated change, fat-depot×disease interactions were modelled separately, and analyses were repeated by sex. All continuous variables were standardised to one SD.

### Causal Mediation Analysis

Bidirectional causal mediation analyses were performed for each kidney substructure and side, to examine how CKD, T2D and hypertension relate to perirenal fat ratio and renal sinus fat, decomposing effects into direct and indirect (mediated) components. Substructure volume and the first ten thickness PC scores were tested with renal adiposity as the outcome, as fat accumulation is a shared downstream consequence of structural kidney change and metabolic disease progression^24,25^. We evaluated: (i) disease-substructure volume-adiposity; (ii) disease-PC score-adiposity; (iii) substructure volume-disease-adiposity; and (iv) PC score-disease-adiposity. This tested both plausible causal directions between disease and kidney phenotype with a common downstream endpoint. This tested both plausible causal directions between disease and kidney phenotype with a common downstream endpoint.

All models were adjusted for the same covariates as in the linear regression models, with continuous variables standardised before analysis. The average causal mediation effect (ACME), average direct effect (ADE), and total effect were estimated using nonparametric bootstrap resampling (5,000 simulations) using the R package *mediation*^26^. Only mediation models for which the ACME, ADE, total effect, and proportion mediated were all statistically significant (p-values<0.05) were considered in the results.

### Longitudinal Analysis

In 2,610 participants with a second imaging visit (after 2.3 years), Wilcoxon rank-sum tests and vertex-wise linear mixed-effects models from the *lme4* package^27^, with participant ID as a random effect were fitted using both time points^28^. Fixed effects were age at first imaging, sex, ethnicity, BMI, WHR, alcohol intake, smoking, dominant HGS, fluid intake, NSAID use, thigh IMAT volume, Townsend deprivation index, CKD, T2D and hypertension, assessing main effects of imaging visit and disease and their interaction. P-values, computed using the *lmerTest* package^29^ were corrected for multiplicity, and FDR < 0.05 was determined significance. All continuous fixed effects were standardised.

### Survival Analysis

Cox proportional hazards models, adjusted for relevant covariates, assessed disease risk after the imaging visit. To reduce dimensionality of the 3D mesh-derived phenotypes, we applied robust sparse PCA using the R package *sparsepca*^30^ (Supplementary Material) and extracted PC scores characterising kidney thickness variation per substructure and side with the first ten modes presented as −3 SD, mean and +3 SD.

Models were run per outcome: a volume model (age, sex, ethnicity, BMI, WHR, alcohol intake, smoking, dominant HGS, fluid intake, NSAID use, thigh IMAT, Townsend deprivation index and substructure volume) and a volume + PCs model adding the PC scores. CKD, T2D, CVD, hypertension and all-cause mortality were defined from hospital, primary care, self-report and death records (Supplementary Table S1). Follow-up was censored at first event, death, or last follow-up (31 May 2024), with a median of 4.8 years, excluding participants with a prior event. Hazard ratios (HRs) with 95% confidence intervals (CIs) and the concordance index (c-index) were reported, with FDR < 0.05 defining significance.

## Results

### Segmentation Performance

The deep learning model achieved a mean Dice similarity coefficient of 0.91/0.90 (left/right cortex), 0.86/0.84 (left/right medulla), and 0.81/0.80 (left/right vessels) on a held-out 20% test set, with visual quality control confirming successful segmentation in all cases.

### Study Population Characteristics

Of 70,270 participants (34,044 men, 36,226 women), 85% were White; mean age was 66.20 ± 7.97 years, BMI 26.56 ± 4.43 kg/m², and WHR 0.89 ± 0.09 (Supplementary Table S2). We identified 1,773 with CKD (2.5% cases; 51% men), 3,972 with T2D (5.7%; 65% men), and 27,838 with hypertension (40%; 57% men).

### Associations with Anthropometric Traits and Disease

We quantified kidney volumes and vertex-wise thickness for each substructure and performed linear regression and mass univariate regression to generate SPMs, adjusting for relevant covariates (Tables 1-2, Figures 1-2, Supplementary Figures S3-S5).

**Figure 1.**
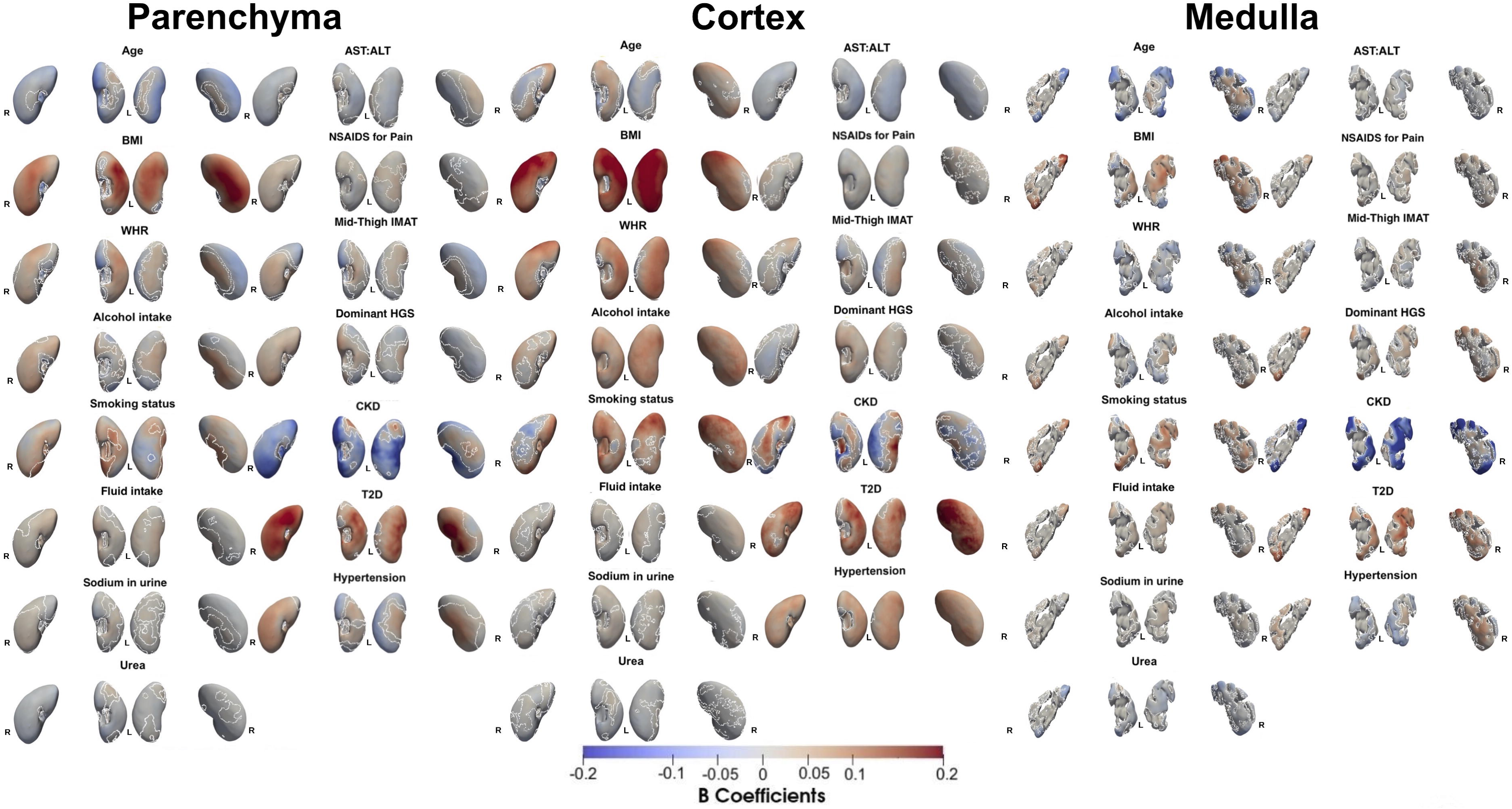
Three-dimensional statistical parametric maps (SPMs) show, for each covariate (age, sex, BMI, waist-to-hip ratio (WHR), alcohol intake, smoking status, fluid intake, sodium in urine, urea, AST:ALT ratio, NSAID use, mid-thigh IMAT, dominant hand grip strength, chronic kidney disease (CKD), type-2 diabetes (T2D), and hypertension), the local strength of association with thickness at each vertex of the kidney parenchyma, cortex, and medulla surface meshes, from mass univariate regression models fitted in the full cohort (N=70,270). For each covariate, the anterior and posterior surfaces of the left (L) and right (R) kidney are shown (anterior views, left plots; posterior views, right plots). White contour lines mark the boundary of statistically significant regions (p < 0.05 after correction for multiplicity). Red indicates a positive (thicker) association and blue a negative (thinner) association. Colour intensity reflects the standardised regression coefficient (*β^*, in units of SD).

**Figure 2.**
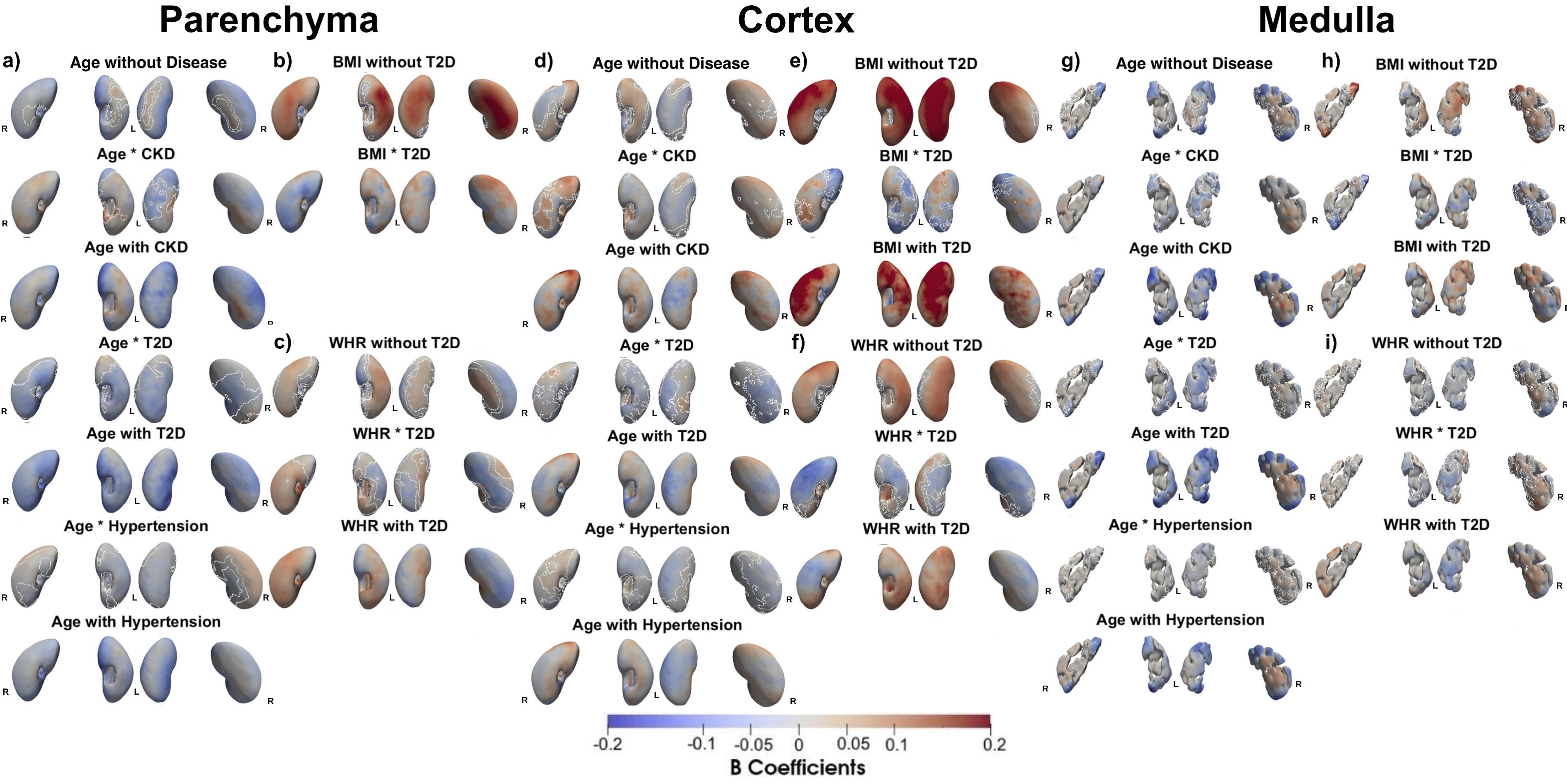
Three-dimensional statistical parametric maps (SPMs) show, for the kidney parenchyma (a-c), cortex (d-f), and medulla (g-i), the local rate of change in thickness for the left (L) and right (R) kidney (anterior views, left plots; posterior views, right plots), in the full cohort (N=70,270). Panels (a, d, g) show the rate of change as a function of age in participants without chronic kidney disease (CKD), type-2 diabetes (T2D), or hypertension, together with the additional change associated with CKD, from the sum of the age and age×CKD regression coefficients, with T2D, from the sum of the age and age×T2D coefficients, and with hypertension, from the sum of the age and age×hypertension coefficients. Panels (b, e, h) show the rate of change as a function of body mass index (BMI) in participants without and with T2D, from the sum of the BMI and BMI×T2D coefficients. Panels (c, f, i) show the rate of change as a function of waist-to-hip ratio (WHR) in participants without and with T2D, from the sum of the WHR and WHR×T2D coefficients. Red indicates a positive (thicker) association and blue a negative (thinner) association. Regression coefficients (*β̂*) are shown with units in standard deviations for each covariate.

**Table 1.**
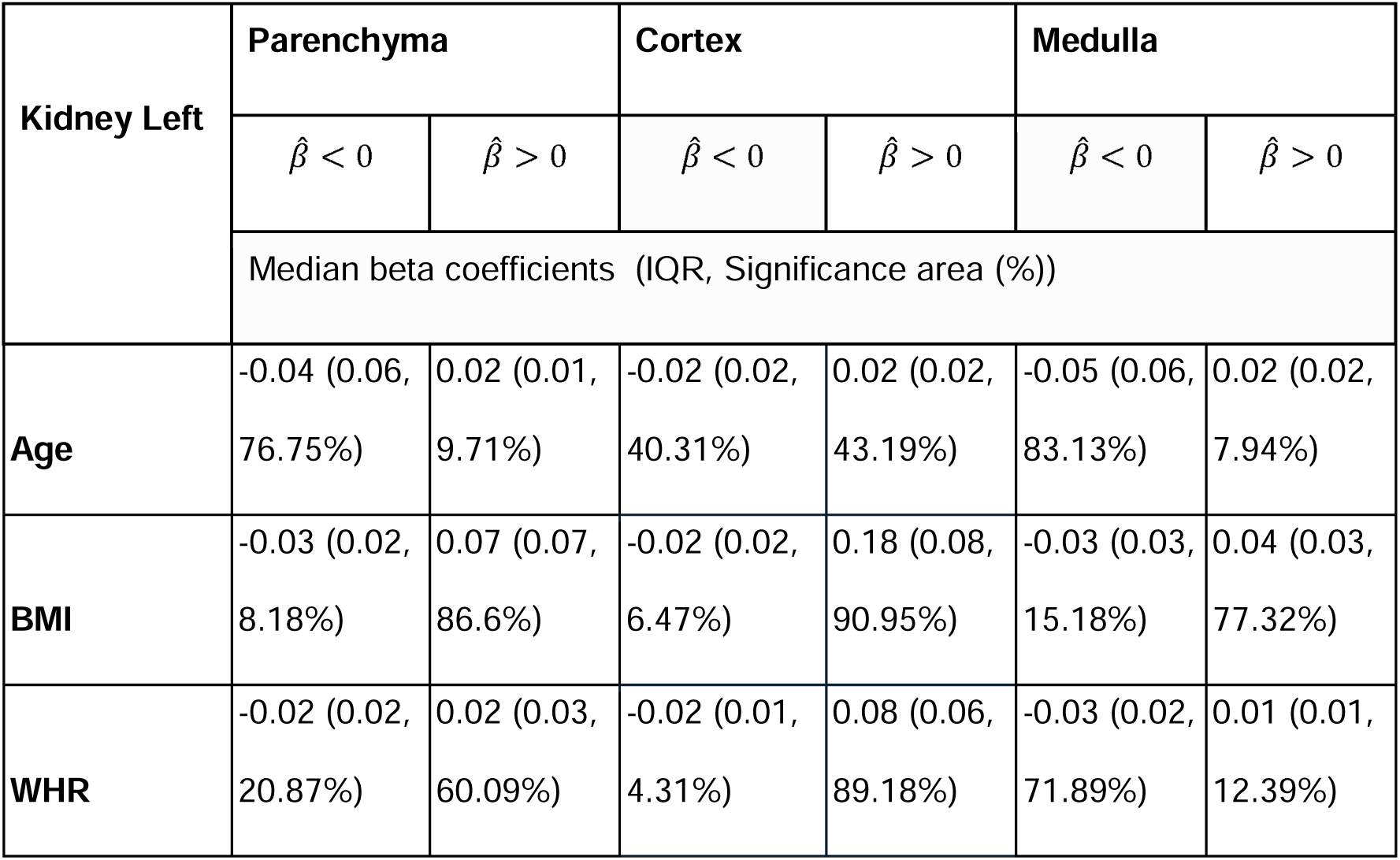
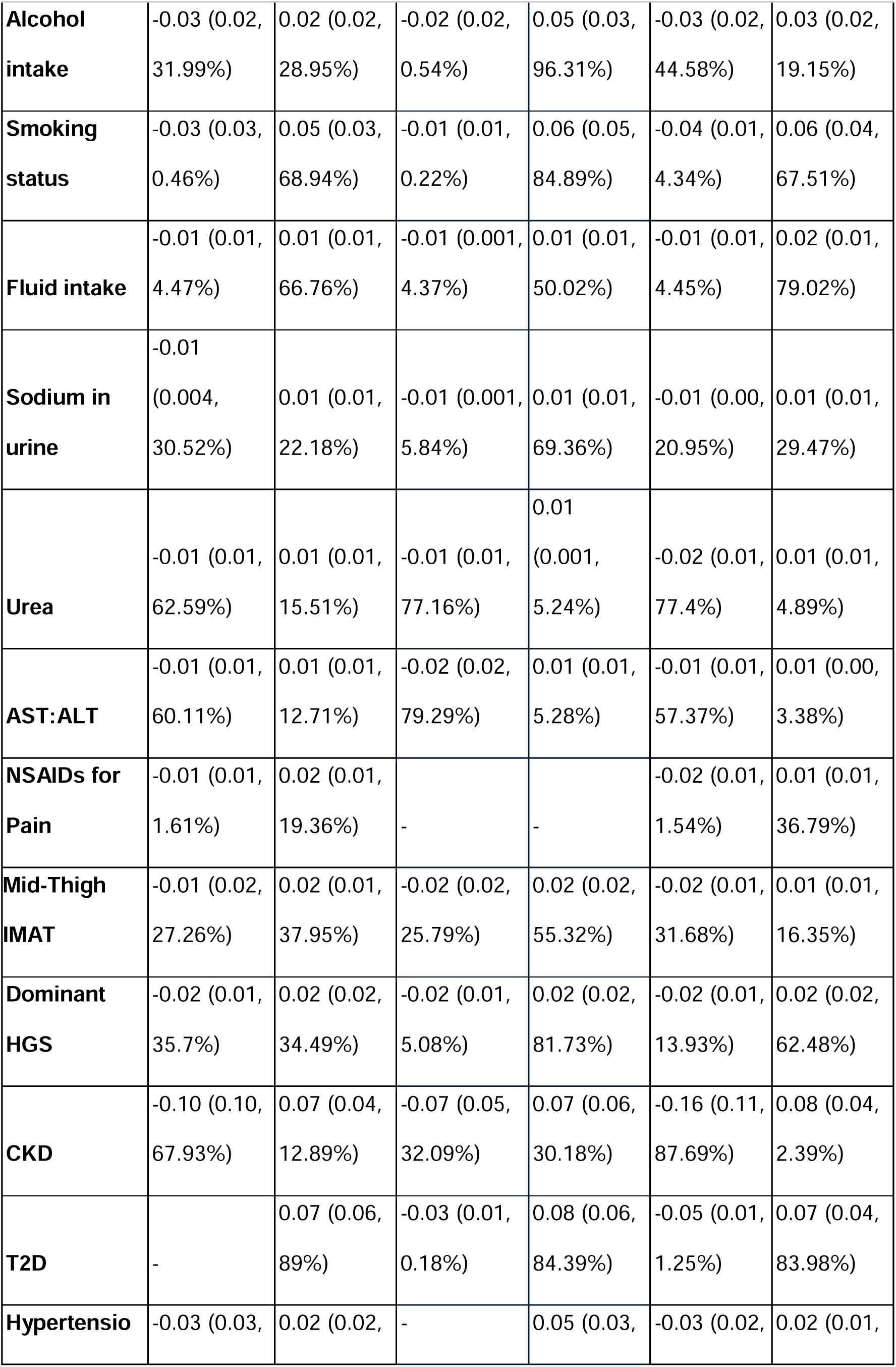
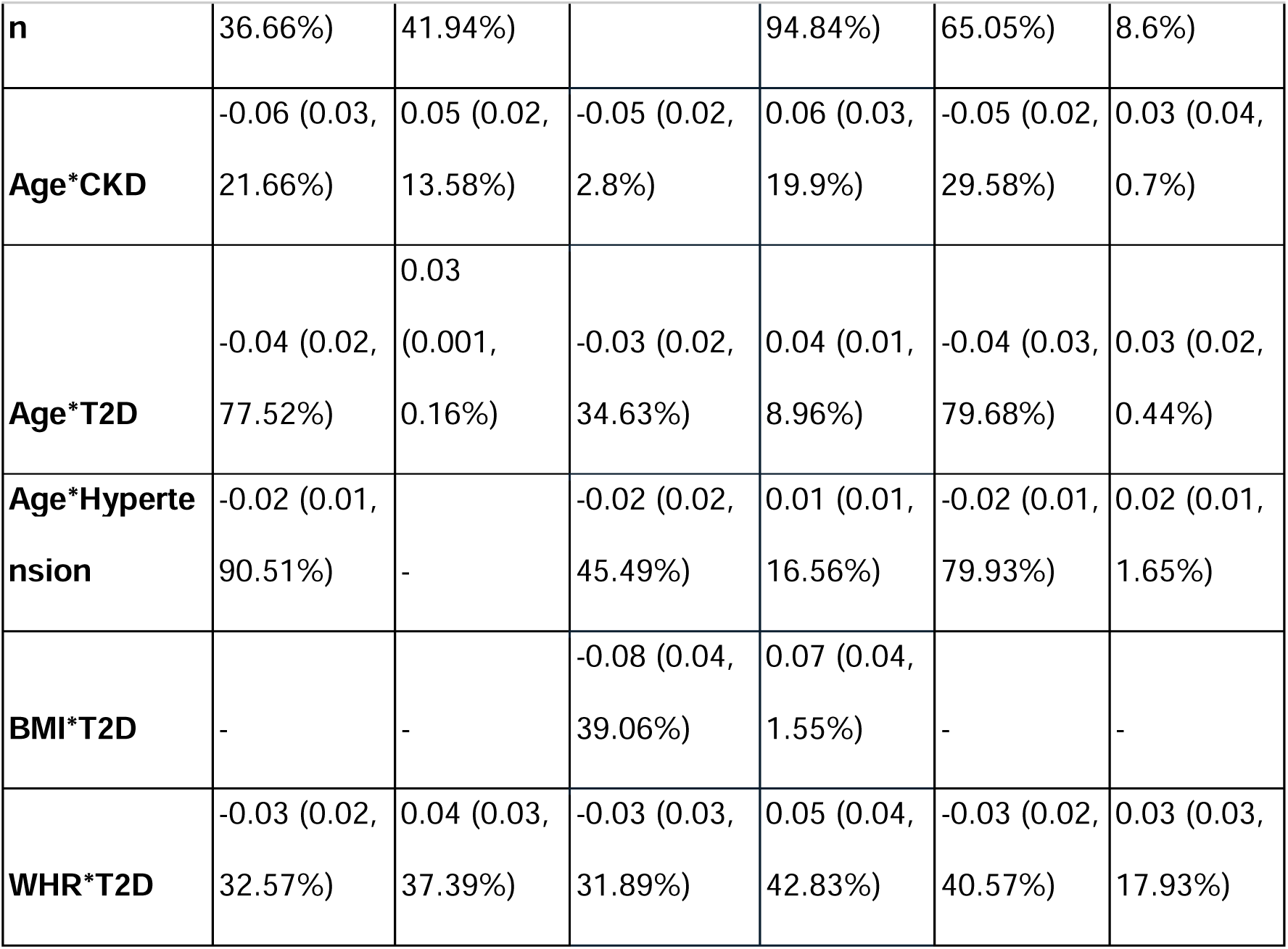
Significance areas for covariates in the MUR model for the anthropometric variables (N=70,270) of the model for the left kidney substructures (parenchyma, cortex and medulla). The total area has been split into areas of positive, negative associations. The standardised regression coefficients (*β^*) are presented as median (interquartile range - IQR) across all vertices of the left kidney substructure’s surface and the significance areas as a percentage (%) of the vertices with statistically significant associations.

**Table 2.**
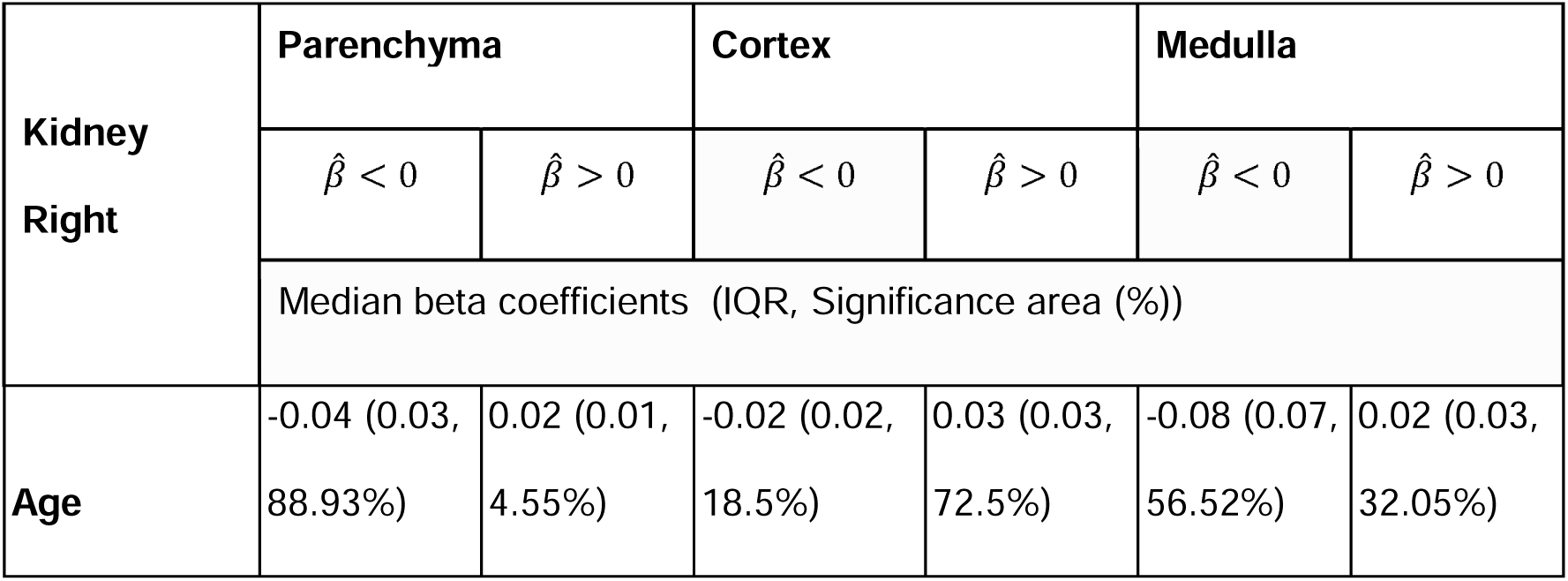
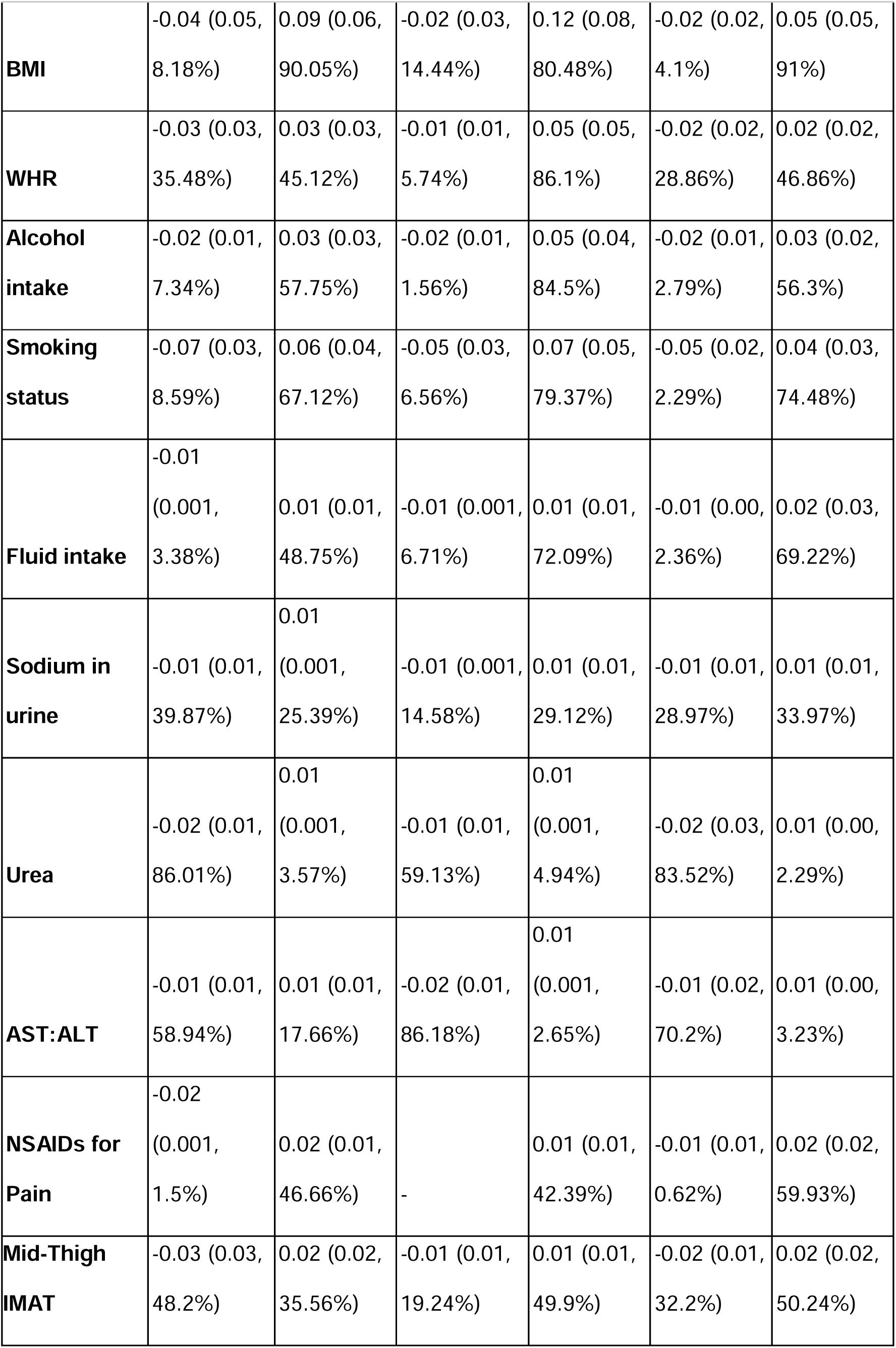
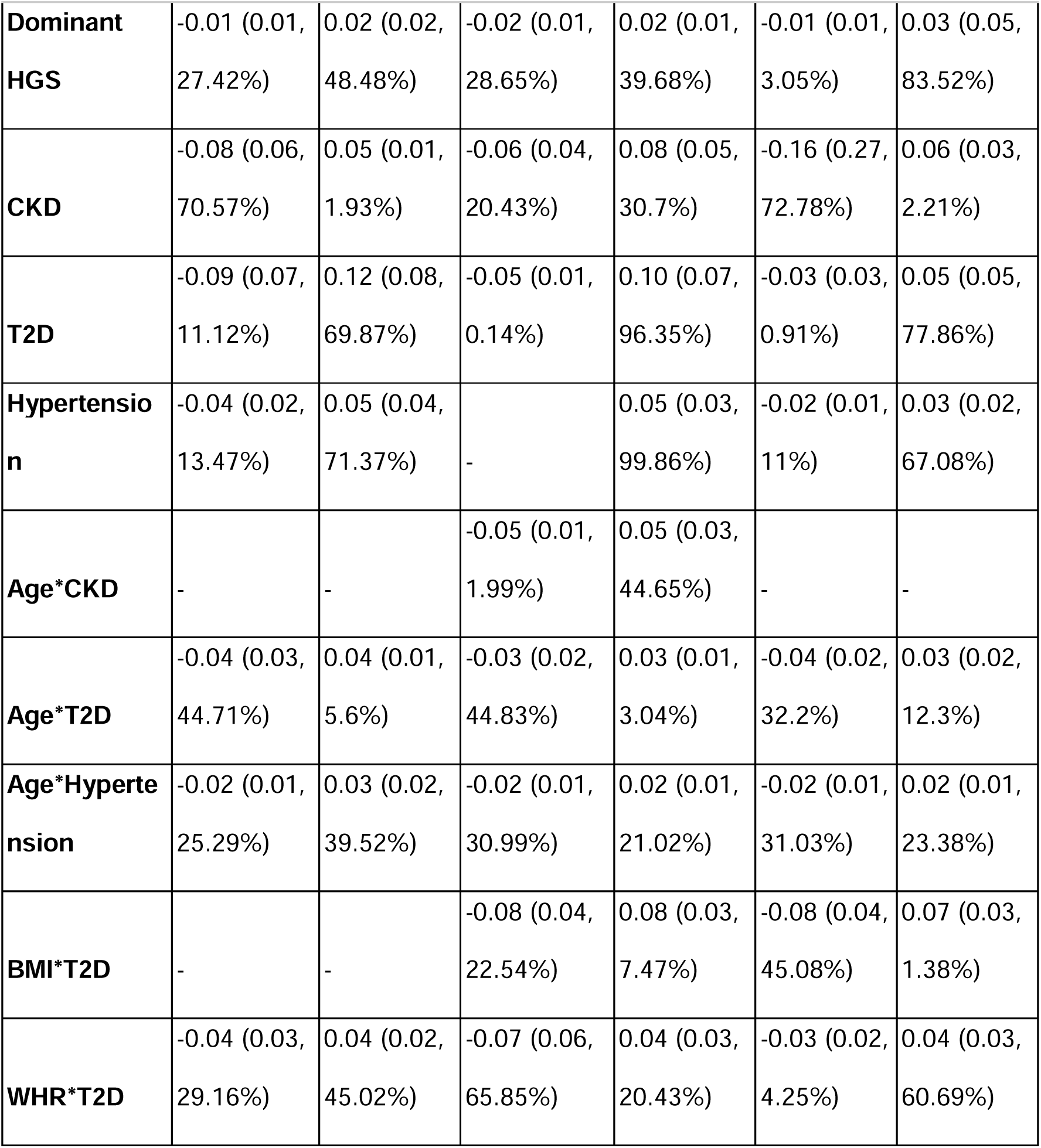
Significance areas for covariates in the MUR model for the anthropometric variables (N=70,270) of the model for the right kidney substructures (parenchyma, cortex and medulla). The total area has been split into areas of positive, negative associations. The standardised regression coefficients (*β^*) are presented as median (interquartile range - IQR) across all vertices of the right kidney substructure’s surface and the significance areas as a percentage (%) of the vertices with statistically significant associations.

Kidney thickness was negatively associated with age at the inferior and superior poles of both kidneys (covering 56.5-88.9% of parenchymal/medullary vertices) and positively associated with age anteriorly and posteriorly. BMI and WHR were positively associated with thickness most extensively in the cortex (80-91% of vertices). Alcohol intake and smoking were also positively associated with cortical thickness (79-96% of vertices). NSAID use covered 36.8% (left medulla) and 59.94% (right medulla). Sodium in the urine showed a positive association with thickness reaching 69.4% (left cortex) and 25.4% (right parenchyma), while urea showed a reduced association reaching 77.4% (left medulla) and 86.01% (right parenchyma).

To aid interpretation, standardised coefficients were converted to unstandardised (raw) mm using the median SD of vertex thickness with 2.74 mm for left parenchyma, 5.00 mm for right parenchyma, 1.61 mm for left cortex, 1.82 mm for right cortex, 2.30 mm for left medulla and 0.46 mm for right medulla. CKD associations were predominantly negative in parenchyma and medulla (67.9-87.7% of vertices; −0.07 to −0.40mm), but showed a more balanced, heterogeneous pattern in the cortex (20.4-32.1% negative vs 30.2-30.7% positive), unlike the uniformly thinner parenchyma and medulla. T2D showed the opposite pattern, with positive associations covering 89%, 84.4%, 84%, 69.9%, 96.4%, and 77.9% of left parenchyma, left cortex, left medulla, right parenchyma, right cortex, and right medulla, corresponding to 0.19, 0.13, 0.16, 0.60, 0.18, and 0.02 mm. Hypertension was also positively associated with cortex thickness, covering 94.8% (left) and 99.9% (right) of the surfaces, corresponding to 0.08 mm (left), 0.09 mm (right) (Supplementary Table S3).

In the left kidney, the age effect (parenchyma −0.04, cortex −0.02, medulla −0.05) was amplified in participants with CKD (age+age×CKD: parenchyma −0.10, cortex −0.07, medulla −0.10, covering 2.8-29.6% additional vertices), with T2D (parenchyma −0.08, cortex - 0.05, medulla −0.09), and with hypertension (parenchyma −0.06, cortex −0.04, medulla - 0.07). In the right kidney, the age×CKD interaction reached significance only in the cortex (age+age×CKD: −0.07, covering 1.99% additional vertices), while age×T2D (parenchyma - 0.08, cortex −0.05, medulla −0.12) and age×hypertension (parenchyma −0.06, cortex −0.04, medulla −0.10) were significant across all three substructures. The BMI×T2D interaction was significant only in the cortex, with an additional negative association covering 39.06% (left) and 22.54% (right) of vertices, superimposed on a positive association with BMI (90.95% left, 80.48% right cortex). A smaller negative association was also present in the interaction with the right medulla (45.1% of vertices). The WHR×T2D interaction showed mixed-direction associations across all three substructures bilaterally, with negative associations covering 4.3-65.9% and positive associations covering 17.9-60.7%, compared with a modest WHR main effect (Table 1, Table 2, Figure 2).

Left renal sinus fat was strongly negative in parenchyma (β=-0.15, 98.4% of vertices) and medulla (β=-0.12, 94.8%), and positive in cortex (β=0.08, 77.2%). CKD interactions reinforced the negative pattern (parenchyma −0.06, 87.5%), while T2D and hypertension attenuated it (parenchyma 0.05/62.3% and +0.04/73.1%). Right renal sinus fat was mixed in parenchyma (47.8% negative vs 47.4% positive) and predominantly positive in cortex and medulla (β=0.08/80.1%, β=0.07/70.2%); CKD interactions shifted associations further positive, whereas T2D and hypertension interactions were negative in the cortex (−0.05/70.0% and −0.03/59.6%). Perirenal fat ratio was positive in the right cortex (β=0.17, 74.3%) and negative in the right medulla (β=-0.10, 69.3%); CKD interactions were negative on both sides, whereas the hypertension cortical interaction reversed direction (left cortex +0.03, 54.5%) (Supplementary Tables S4-S7 and Figure S6).

### Associations by Sex

Sex-stratified analyses broadly recapitulated the full-cohort pattern (Supplementary Figures S7-S8, Tables S8-S9). BMI showed very strong positive cortical associations in men (99.11% of left and 92.8% of right cortical vertices); WHR showed more extensive negative parenchymal/medullary associations in men than women (left parenchyma 35.58% vs 21.09%), whereas women showed slightly more extensive positive cortical WHR associations (83.98% vs 81.73% left). CKD medullary associations were slightly stronger in men (left: −0.17 vs −0.16; right: −0.19 vs −0.18), T2D cortical associations were larger in women (right cortex 0.13 vs 0.10), and hypertension associations were similar between sexes.

### Longitudinal Changes in Kidney Thicknesses

Among 2,610 participants with a second imaging visit, we observed small but statistically significant changes in thickness between visits (Supplementary Figure S9). In the linear mixed-effects models, the left kidney showed significant associations across 17.08%/14.81% (parenchyma; negative/positive), 11.02%/6.75% (cortex; negative/positive), and 8.16%/3.01% (medulla; negative/positive) of vertices. The right kidney showed smaller effects, up to 9.69%/6.72% (medulla, negative/positive). No significant imaging-visit interaction with CKD, T2D, or hypertension was observed in either kidney (Figure 3, Supplementary Tables S10-S11). Substructure volume also decreased significantly in the parenchyma, medulla, and left cortex (Supplementary Figures S10-S11).

**Figure 3.**
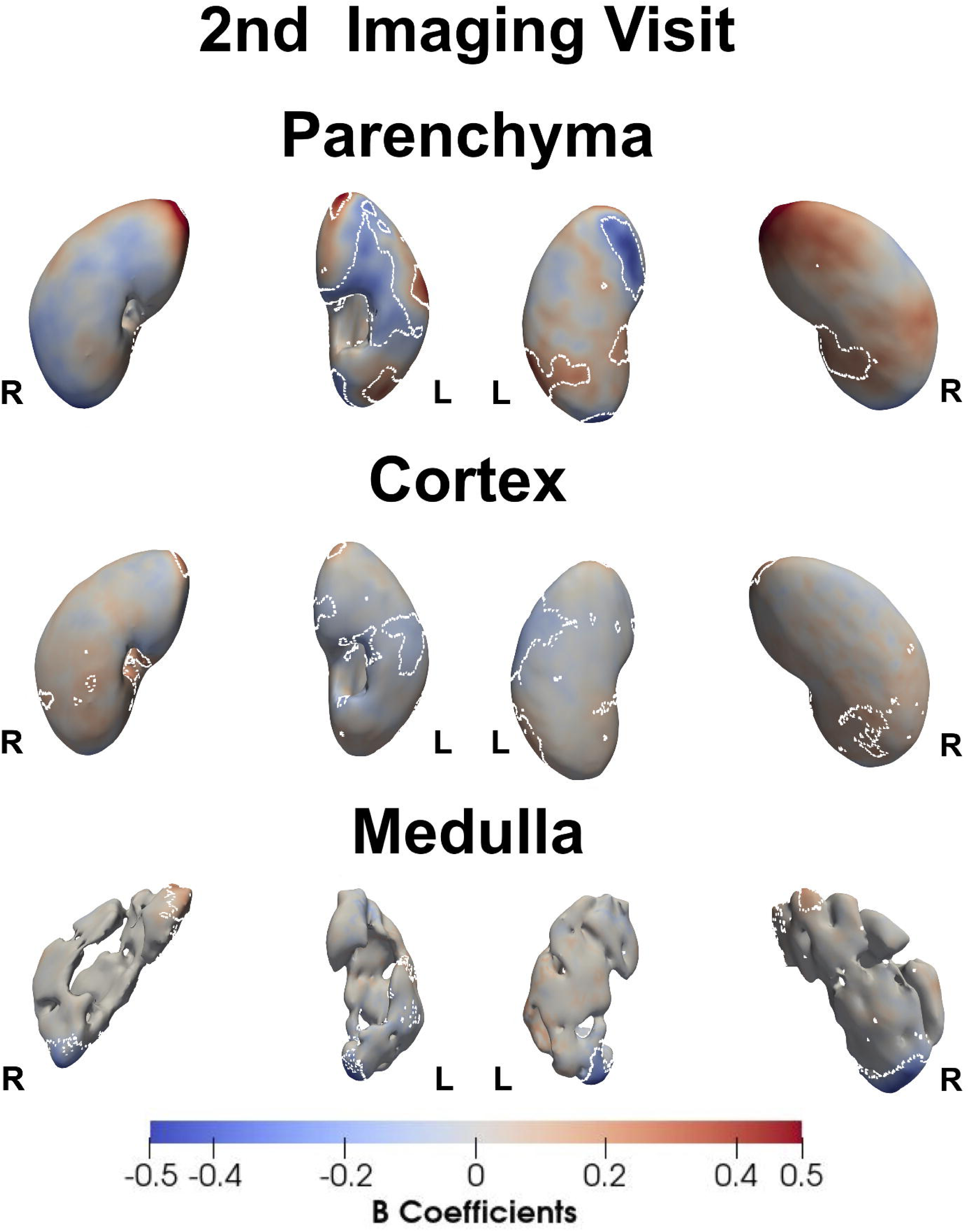
Three-dimensional statistical parametric maps (SPMs) show the local strength of association between kidney substructure (parenchyma, cortex, medulla) thickness and imaging visit (baseline vs. follow-up, approximately 2.3 years later), from linear mixed-effects models in the longitudinal cohort (N = 2,610), for the left and right kidney (anterior views, left plots; posterior views, right plots). White contour lines mark the boundary of statistically significant regions (FDR-corrected p < 0.05); red indicates a positive (thicker) association at follow-up and blue a negative (thinner) association. Colour intensity reflects the standardised regression coefficient (*β̂*, in units of SD).

### Mediation effects in the association of kidney phenotyping, disease and kidney adiposity

SPCA captured over 50% of thickness variation across the first ten PCs in all substructures (Supplementary Figure S12, Video S1). Mediation pathways with significant direct and indirect effects (p<0.05), high proportion of mediation and direction-consistent are highlighted (Supplementary Data SD1).

Smaller medulla volume was associated with CKD prevalence bilaterally and with higher renal sinus fat, with significant indirect effects (ACME: right 0.004, p<0.001; left 0.014, p<0.001) mediating 6.3% (right, p=0.003) and 21.6% (left, p<0.001) of the association, alongside significant direct effects (ADE: right 0.058, p=0.004; left 0.052, p=0.015) (Supplementary Figure S13).

Cortical thickness PC-based mediation revealed further effects. Hypertension was significantly associated with PC1, which predominantly reflects global cortical thickness, thinner-to-thicker across nearly the entire right cortex, and reverse pattern on the left. While PC1 was significantly associated with perirenal fat ratio, yielding ACME = 0.026 (right, p < 0.001) and 0.014 (left, p < 0.001) and mediating 39.2% and 24.1% (both p<0.001; Supplementary Figure S14). The CKD–PC1–renal sinus fat pathway was more modest but significant (ACME = 0.015 right, p<0.001; 0.008 left, p=0.009), mediating 24.3% (right, p=0.004) and 12.6% (left, p=0.009; Supplementary Figure S15). T2D was significantly associated with PC3, representing the variation at the inferior pole and medial border near the hilum, with little change at the superior pole, which in turn was associated with perirenal fat ratio (ACME = −0.007 right, −0.011 left, both p<0.001), mediating 20.2% (right, p=0.001) and 16.2% (left, p<0.001; Supplementary Figure S16). CKD was also significantly associated with PC4, reflecting localised foci along the medial, hilar aspect, one superior, one central, which was associated with renal sinus fat (ACME = 0.004 right, p=0.011; 0.012 left, p<0.001), mediating 7.0% (right, p=0.013) and 18.4% (left, p=0.002) (Supplementary Figure S17). As a sensitivity analysis, we reversed the ordering (disease - kidney fat - cortical PC1, PC3 and PC4, Supplementary Data SD2). In the right kidney, perirenal fat ratio mediated hypertension–PC1 (ACME 0.020, ADE 0.111; 15.5%, p<0.001) and T2D–PC3 (−0.008, −0.069; 10.9%, p=0.001), and renal sinus fat mediated CKD–PC1 (0.010, 0.090; 9.9%, p=0.001) and CKD–PC4 (−0.007, −0.066; 9.0%, p=0.014). In the left kidney, only T2D–PC3 via perirenal fat ratio was significant (0.009, 0.082; 10.2%, p<0.001).

### Survival Analysis

We assessed whether volume and thickness had predictive value for incident disease diagnosed after the imaging visit, fitting a model per outcome using kidney volume, and the first 10 thickness PC scores, separately per kidney (Figures 4-5, Supplementary Figures S18-S34).

**Figure 4.**
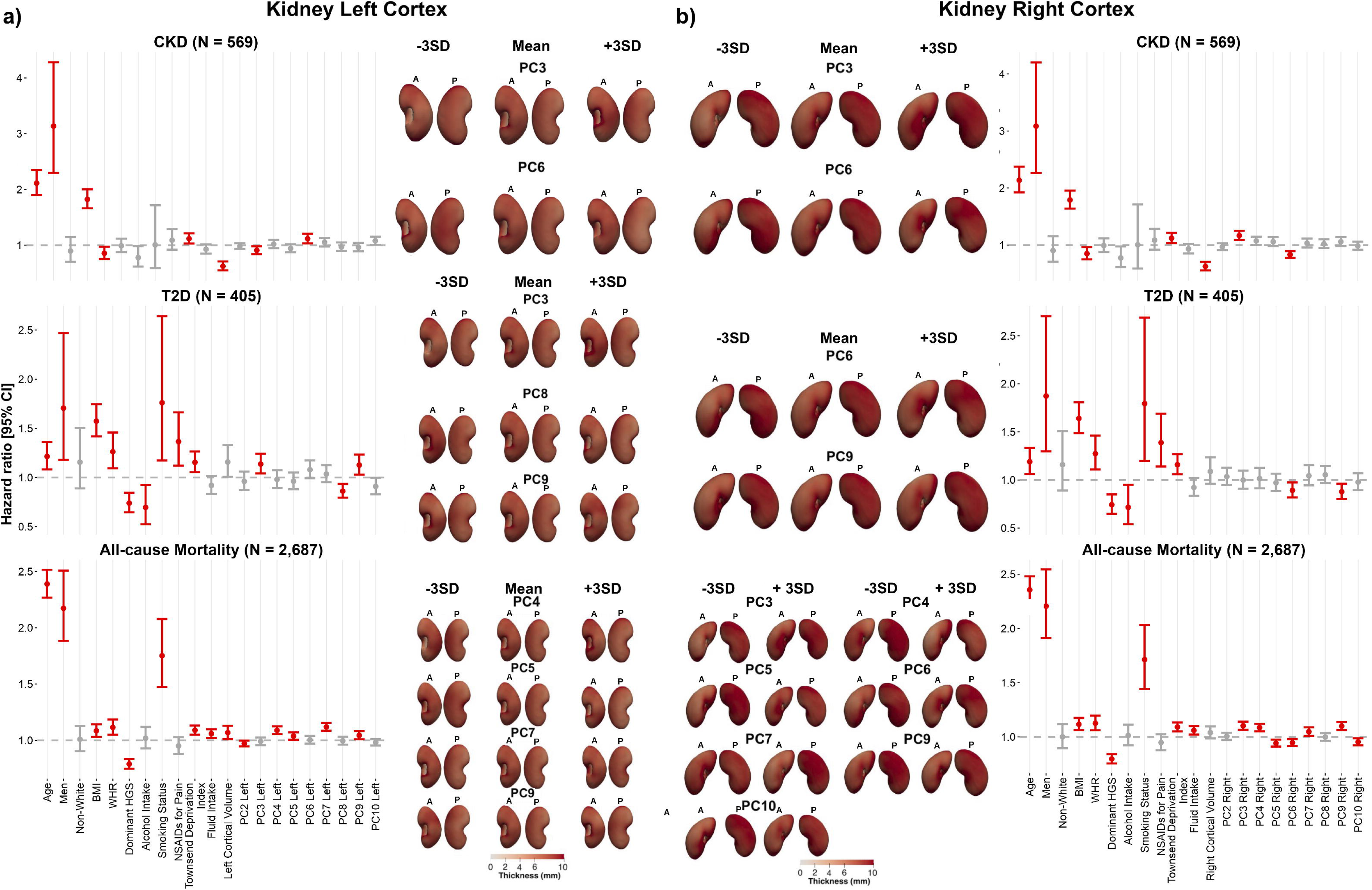
Kidney cortex thickness principal components (PCs) and their associations with incident disease. Forest plots show hazard ratios (HR) and 95% confidence intervals (CI) for incident chronic kidney disease (CKD), type-2 diabetes (T2D), and all-cause mortality for the a) left and b) right kidney cortex, from Cox models adjusted for age, sex, ethnicity, body mass index, waist-to-hip ratio, dominant hand grip strength, alcohol intake frequency, smoking status, NSAID use for pain, Townsend deprivation index, fluid intake, kidney cortical volume, and the first ten thickness principal component (PC) scores (excluding PC1, which reflects overall cortical thickness). Significant associations (FDR-adjusted p < 0.05) are shown in red and non-significant associations in grey. For each outcome, the PC modes significantly associated with that outcome are visualised as 3D kidney renders at −3 SD, mean, and +3 SD of that mode, in anterior (A) and posterior (P) view, coloured by thickness (mm).

**Figure 5.**
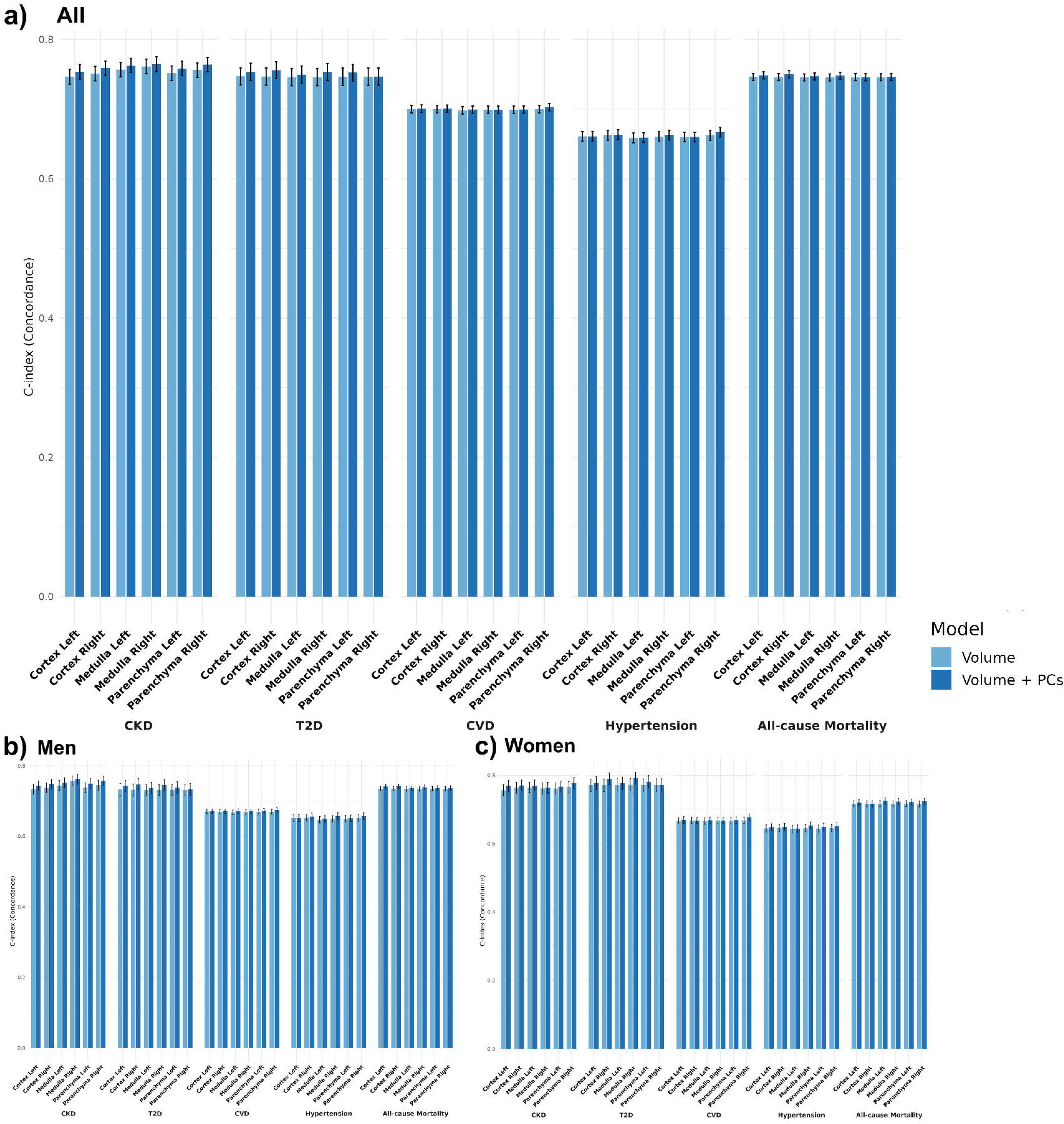
Performance for incident disease between kidney volume alone and kidney volume plus thickness principal components (PCs). Bar plots show the concordance index (C-index, with 95% CI) of Cox proportional hazards models for incident chronic kidney disease (CKD), type-2 diabetes (T2D), cardiovascular disease (CVD), hypertension, and all-cause mortality, for each kidney substructure (cortex, medulla, parenchyma) and side (left, right), comparing a model with kidney volume alone (light blue) against a model with kidney volume plus the first ten thickness PC scores (dark blue). Results are shown for (a) the full cohort, (b) men, and (c) women.

Kidney volume was inversely associated with incident CKD across all substructures and sides (hazard ratio (HR) 0.55-0.62, all p<10⁻ ¹²), consistently in both sexes (Supplementary Figures S18-S23). In the left cortex, several thickness PC scores were independently associated with incident CKD (Figure 4); PC3 (HR 0.91, 95% confidence interval (CI) [0.84-0.98], p=0.015) reflected thickness variation at the inferior pole and medial/hilar border. Volume was a weaker predictor of incident T2D (significant only in the left cortex, HR 1.15 [1.01-1.30]); PC9 (HR 1.13, [1.03-1.23], p=0.009; medial/hilar cortical thickness) and PC8 (HR 0.86, p<0.001) were the most significant PC scores for T2D (Figure 4). PC7 (left, HR 1.12 [1.09-1.15], p=5.1×10⁻¹³; hilar concavity and inferior-pole thickness) and PC3 (right, HR 1.10 [1.07–1.14], p=1.6×10⁻; inferior pole and hilar border) were associated with incident all-cause mortality. The largest c-index gains from adding PCs to volume were for CKD in the left cortex (0.747 to 0.754) and T2D in the right cortex (0.747 to 0.756) (Figure 5).

## Discussion

We mapped local kidney substructure thicknesses at population scale, testing whether 3D parenchymal, cortical, and medullary thickness mapping could reveal disease-, adiposity-, and anthropometry-specific signatures that volume alone conceals. Statistical parametric mapping has previously localised phenotype-disease associations in abdominal^13–15^, neuroimaging^11^ and cardiac imaging^12^. Here, we assessed regionally distinct, disease-specific kidney remodelling, adiposity-mediated pathways, and incident disease prediction (CKD, T2D, hypertension, mortality) beyond what global volume alone could capture.

Thickness declined with age at the renal poles, consistent with reports that cortical volume falls while medullary volume is relatively preserved in healthy renal ageing^3,4^. CKD, T2D and hypertension amplified this decline, suggesting these diseases compound rather than simply co-occur with structural renal ageing. BMI and WHR were positively associated with cortical thickness, in line with hyperfiltration-related hypertrophy in early metabolic disease^7,9,31,32^. CKD was associated with thinning, most pronounced in the parenchyma and medulla, consistent with reduced kidney volume in lower kidney function^3,8,33^, with focal cortical increases consistent with compensatory hypertrophy of residual nephrons amid atrophy^34^. T2D and hypertension showed the opposite, predominantly positive association, consistent with compensatory hypertrophy rather than atrophy at this disease stage^6,9,32,35^. The NSAID-medullary association spatially corresponds to the renal papilla and medulla, the site of analgesic-induced papillary necrosis^34^, raising the possibility that our maps are sensitive to subclinical analgesic nephrotoxicity.

Perirenal fat has been linked to reduced GFR and altered renal haemodynamics^17,36^, and renal sinus fat to intrarenal haemodynamic abnormality independent of visceral adiposity^18^, potentially via paracrine signalling, mechanical compression, or shared cardiometabolic pathways^37,38^. Perirenal fat ratio was independently associated with thickness and modified by CKD, T2D, and hypertension, supporting a local, kidney-specific contribution to risk. Renal sinus fat showed a left-right contrast, consistent with asymmetric venous drainage^39^. CKD interactions reinforced baseline fat-thickness associations, consistent with compounding mechanical/paracrine injury, whereas T2D and hypertension interactions often attenuated or reversed them, particularly in the cortex, reflecting concurrent hyperfiltration-related hypertrophy, consistent with disease-stage-dependent fat remodelling^24^ and afferent-arteriolar mediation of perirenal fat effects^25^.

Over a median 2.3-year follow-up, thickness changes were small but significant, with no interaction by disease, suggesting these associations reflect pre-existing structural differences rather than short-term remodelling.

Causal mediation analysis extended these adiposity findings mechanistically. Smaller medulla volume partly mediated the association between CKD and higher renal sinus fat bilaterally. Cortical thickness PCs mediated a substantial share of the hypertension-perirenal fat and CKD-renal sinus fat associations, with CKD-cortical thickness pathway reflecting hilar/medial thinning, and renal sinus fat. These findings position local cortical and medullary thickness, rather than volume alone, as an intermediate on the causal path between disease and kidney fat accumulation, consistent with mechanical and paracrine mechanisms proposed for perirenal and sinus fat^37,38^. Given that renal sinus fat is increasingly implicated as a modifiable cardiorenal risk factor^17,18,36^, spatially resolved thickness phenotyping may help identify which patients’ kidney-fat relationships are structurally, rather than purely adiposity-driven.

Kidney volume was consistently inversely associated with incident CKD, replicated across substructure, side, and sex, whereas parenchymal and cortical volumes were risk factors for hypertension and CVD, indicating that kidney size is not a single biological axis. This reflects protective nephron mass in CKD but hyperfiltration-associated hypertrophy in hypertension and CVD^9,32^. Thickness-derived PC scores added incremental discrimination beyond volume, with the highest performance for CKD and T2D. Cortical thickness variation concentrated at the inferior pole and medial/hilar border was independently associated with incident CKD. This spatial pattern may reflect the higher density of medullary nephrons and vascular convergence near the renal hilum. This indicates that substructure-level thickness captures disease-relevant morphology that a single global size metric cannot.

Strengths include the large imaging cohort, automated substructure segmentation, bilateral analysis, spatially resolved thickness maps, local fat-depot quantification, sex-stratified and interaction models, and linkage to five incident outcomes. Limitations include the observational design and UK Biobank’s healthy-volunteer selection bias^40^, predominantly European ancestry, and exclusion of more severely affected individuals, although risk-factor associations in UK Biobank have previously been shown to be broadly generalisable ^41,42^. Disease definitions combined multiple coding sources and inevitably include some heterogeneity in timing and severity; medication use and reverse causation cannot be fully excluded, and albuminuria, a key component of CKD staging and an independent cardiovascular risk marker^43^, was not available for the full cohort. Furthermore, the lack of renal histology is a well-known gap in imaging-based kidney phenotyping^44^, in particular given the insights it provides into age-and disease-related structural involution of the kidney^45^. Follow-up (median 4.8 years) was adequate for CKD, CVD, hypertension and mortality but yielded comparatively few T2D events, limiting power for those outcomes. With longer follow-up, external replication, and multimodal imaging, these 3D phenotypes have clear potential for clinical utility as risk markers.

## Conclusions

In conclusion, 3D kidney substructure thickness mapping reveals disease- and adiposity-specific structural signatures that volume alone conceals. CKD was marked by parenchymal and medullary thinning with a mixed cortical pattern and reduced volume, whereas T2D and hypertension showed predominantly thicker remodelling and larger volume, with perirenal and renal sinus fat acting locally on thickness. Thickness-derived principal components consistently improved disease prediction beyond volume, particularly for CKD and T2D, establishing population-scale 3D kidney phenotyping as a potentially clinically relevant tool for characterising kidney structural disease.

## Declarations

### Ethics approval and consent to participate

The data resources used in this study have approval from ethics committees. Full anonymised images and participants metadata from the UK Biobank cohort was obtained through UK Biobank Access Application number 44584. The UK Biobank has approval from the North West Multi-Centre Research Ethics Committee (REC reference: 11/NW/0382), and obtained written informed consent from all participants prior to the study. All methods were performed in accordance with the relevant guidelines and regulations as presented by the relevant authorities, including the Declaration of Helsinki https://www.ukbiobank.ac.uk/learn-more-about-uk-biobank/about-us/ethics.

### Consent for publication

Not applicable.

### Availability of data and materials

The data that support the findings of this study are available from the UK Biobank (https://www.ukbiobank.ac.uk), but restrictions apply to the availability of these data, which were used under license for the current study, and so are not publicly available. Data are however returned by us to the UK Biobank where they will be fully available on request.

### Author Contributions

J.D.B., E.L.T. and M.T. conceived the study. J.D.B., N.B., M.N., B.W., E.L.T., and M.T. designed the study. M.T., N.B. and B.W. implemented the methods and performed the data analysis. M.T. defined the disease and physiological condition categories. C.B-B. performed the kidney substructure annotations and R.S. performed the deep learning segmentation models. M.T. and N.B. performed the image analysis and M.T. performed the statistical analysis. M.T., R.S., R.L., A.S.L., N.B., B.W., M.N., M.Tom., J.D.B., and M.C. drafted the manuscript. All authors read and approved the manuscript.

### Declaration of Generative AI and AI-assisted technologies in the writing process

During the preparation of this work, the authors used Claude and Grammarly to rephrase text to improve readability and reduce the overall word count. After using these tools, the authors reviewed and edited the content as needed and take full responsibility for the content of the publication.

## Acknowledgements

We thank Alex Chekholko for providing technical support that expedited our work. This research has been conducted using the UK Biobank Resource under Application Number 44584.

## Abbreviations

CKD: chronic kidney disease;
T2D: type 2 diabetes;
CVD: cardiovascular disease;
BMI: body mass index;
WHR: waist-to-hip ratio;
HGS: hand grip strength;
eGFR: estimated glomerular filtration rate;
NSAIDs: nonsteroidal anti-inflammatory drugs;
AST: aspartate aminotransferase;
ALT: alanine aminotransferase;
IMAT: intermuscular adipose tissue;
MUR: mass univariate regression;
TFCE: threshold-free cluster enhancement;
SPM: statistical parametric map;
SSA: statistical shape analysis;
PCA: principal component analysis;
PC: principal component;
ACME: average causal mediation effect;
ADE: average direct effect;
HR: hazard ratio;
CI: confidence interval;
IQR: interquartile range;
FDR: false discovery rate.

