## Supplementary_Material for "Beyond Kidney Volume: 3D Mapping of Parenchymal, Cortical and Medullary Thickness with Disease, Anthropometry and Kidney Adiposity"

**Phenotype Definitions**

Anthropometric measurements including age, body mass index (BMI), waist and hip circumferences, and systolic and diastolic blood pressure were taken at the UK Biobank imaging visit, and ethnicity was defined based on the self-reported ethnic background at the initial assessment visit (field: 21000). For the purpose of our analysis we categorised ethnic background as follows: 0 for ”White” and 1 for any other ethnic background (due to small numbers of non-white participants in this dataset (3.1%)). Sex was self-reported and included those recorded by the NHS and those obtained at the initial assessment visit (field: 31).

Biological samples for serum creatinine (field: 30700) were measured in $millimole/L$, sodium in urine (field: 30530) in $millimole/L$ and urea (field: 30670) in $millimole/L$ units were taken on the initial assessment visit. Estimated glomerular filtration rate (eGFR) was calculated based on the CKD-EPI creatinine equation (2009)^1^ as follows:

$eGFR=141 \times min(Scr/\kappa, 1)^{\alpha} \times max(Scr/\kappa, 1)^{-1.209} \times0.993^{Age} [\times1.018 if female],$ $[\times1.159 if black]$.

where $Scr$ is serum creatinine in converted into $mg/dL$ units, $\kappa$ is 0.7 for females and 0.9 for males, $\alpha$ is −0.329 for females and −0.411 for males, $min$ is the minimum of Scr/κ or 1, and $max$ is the maximum of Scr/κ or 1. In the aforementioned equation, ethnicity for “Black” was defined based on the continental genetic ancestry for “African” ancestry ([https://pan.ukbb.broadinstitute.org](https://pan.ukbb.broadinstitute.org/docs/technical-overview)) and if missing from the self-reported ethnic background (field: 21000) for “Black or Black British”. It should be noted that the UK Biobank initial assessment visit preceded the imaging visit by 9 ± 1.7 years.

Questionnaire information from the UK Biobank imaging visit was used to determine alcohol intake frequency (field: 1558) and smoking status (field: 20116) and fluid intake. For our analysis, we categorised alcohol intake frequency as 1 for “Daily or almost daily” and 0 otherwise, smoking status as 1 for ”Current” and 0 for “Previous” and “Never” and fluid intake as the sum of tea (field: 1488), water (field: 1528) and coffee (field: 1498) intake in cups or glasses/day.

Pain-related phenotypes were derived using touchscreen fields collected at the imaging visit assessing pain experienced in the last month in specific body sites: neck/shoulder pain (field: 3404), back pain (field: 3571), hip pain (field: 3414), knee pain (field: 3773), and general pain throughout the body (field: 2956). Pain type(s) experienced in the last month (field: 6159) was used to exclude participants reporting only headache, facial pain, stomach/abdominal pain, "prefer not to answer" or "none of the above", such that the derived pain phenotype captured musculoskeletal/general pain specifically^2^. Self-reported medication use for pain relief was captured from medication for pain relief, constipation, heartburn (field: 6154, categories "Ibuprofen" and "Aspirin") and treatment/medication code (field: 20003)^3,4^.

Pain diagnoses were also identified from GP clinical records using a pre-specified list of Read v2/CTV3 codes spanning five musculoskeletal diagnostic categories: spine disorders, arthropathy, soft tissue disorders, muscle disorders, and bone disorders. Nonsteroidal anti-inflammatory drug (NSAIDs) prescriptions were identified from GP prescription records by matching the drug name field against: ibuprofen, aspirin, diclofenac, naproxen, ketoprofen, piroxicam, ketorolac, indometacin, celecoxib, etoricoxib, nefopam, diflunisal, etodolac, fenoprofen, flurbiprofen, mefenamic acid, meloxicam, nabumetone, oxaprozin, sulindac, and UK brand names Disprin, Nurofen, Naprosyn, Voltarol, Celebrex, Brufen, Calprofen, Fenbid, Ibugel, and Ibuleve^3^. NSAIDs for pain treatment was defined based on both self-reported (touchscreen questionnaire) and linked primary care (GP) data and was coded as 1 if they met the criteria for either the self-reported definition or the GP-linked pain-treatment-with-NSAIDs definition, and 0 otherwise.

**Disease Definitions**

We selected diseases known to be associated with kidney health, and those previously associated with changes in kidney volume^5^. In part, this was also guided by the number of patients available in the UK Biobank with diseases of interest. Disease categories of interest, including CKD, T2D and hypertension were selected based on their influence on body composition and organ health, as well as their frequency within the UK Biobank. They were defined at the time and before the imaging visit. A summary of the codes corresponding to the considered disease traits are provided in supplementary Table S1.

For the definition of CKD, we calculated eGFR using the CKD-EPI equation (see methods section) and baseline cohort data. In a previous study^6^, mild to moderate CKD was defined by an eGFR between 15 and 60 ml/min/1.73 m² or by the International Classification of Disease (ICD) codes N18.1-N18.5. We define CKD by eGFR below 60 ml/min/1.73 m², and the ICD10 and self-reported codes for chronic kidney disease, kidney dialysis and kidney transplantation operation codes (<https://archive.datadictionary.nhs.uk/DD%20Release%20July%202022/Covid19PRA/Chronic_Kidney.html>). The codes for T2D were selected based on the ICD10 and self-reported codes for type-2 diabetes^7^. Hypertension was defined as self-report of hypertensive medication use, or mean blood pressure ≥ 140/90 mmHg^8^ at the imaging visit, or a prior diagnosis of hypertension. The codes for hypertension diagnosis were selected based on the ICD10 and the self-reported codes for hypertension. Hypertensive medication codes were selected from the medication fields, reporting as regularly taking blood pressure medication. Codes for CVD were selected based on the general presence of cardiac disease as previously described^9^.

CKD, CVD, T2D and hypertension diagnosed or reported after the imaging visit was defined based on ICD10 codes from the various sources, the date of first diagnosis made available as “first occurrence” data (Category 1712) and based on self-reported codes described in supplementary Table S1, defined after the imaging visit. All-cause mortality events occurring after the imaging visit were similarly identified from the date of death field, with participants censored at their date of death or at the end of mortality follow-up, whichever occurred first.

### **Image Processing**

### *Deep Learning Architecture*

### We segmented the kidney and other abdominal structures from neck-to-knee Dixon MRI using a 3D U-Net-style encoder-decoder with skip connections (five encoding levels, four decoding levels) ^10^. Each encoding stage applied two 3×3×3 convolutions with instance normalisation and ReLU activation, followed by 2×2×2 max-pooling; channel depth doubled at each level, from 32 filters initially to 512 at the bottleneck. Each decoding stage upsampled via transposed convolution, concatenated the corresponding skip connection, and applied a further two convolutional layers. Five channels (fat, water, in-phase, out-of-phase, and a body mask) formed the network input, zero-padded from the native 370×174×224 voxels to 384×176×224 to cover the full field of view. A single 1×1×1 convolution produced a two-class (background/foreground) output per structure. The shared trunk comprised approximately 22 million trainable parameters, plus ~65,000 per structure-specific head, implemented in PyTorch and PyTorch Lightning^11^.

### *Loss Function*

### Segmentation quality was optimised using the soft Dice loss, which directly maximises the overlap between predicted and ground-truth masks^12^. For a binary segmentation task it is defined as:

$$L_{Dice}= 1-\frac{2\sum_{i=1}^{N} p_{i}g_{i}+\epsilon}{\sum_{i=1}^{N} p_{i}^{2}+\sum_{i=1}^{N} g_{i}^{2}+\epsilon}$$

### Here, $p_{i}$ is the predicted probability and $g_{i}$ is the ground-truth label at voxel i (summed over all N voxels), and ϵ (1×10⁻⁶) is a small constant added for numerical stability. Probabilities were taken after softmax, allowing the loss to be optimised by gradient descent; multi-task pre-training combined this with binary cross-entropy in equal weight across all 28 organs, while fine-tuning used the Dice loss alone^13^.

### *Training Strategy*

### Training used a two-stage transfer-learning strategy to scale efficiently to many structures. First, a shared encoder-decoder trunk with 28 independent output heads was pre-trained to jointly segment major organs and tissues, supervised by the combined Dice and binary cross-entropy loss, equally weighted and aggregated across all organs and training samples. Second, each organ-specific model was initialised by copying the pre-trained trunk and randomly initialising a new classification head, then fine-tuned using the Dice loss alone; this transfer-learning step enabled effective training despite limited structure-specific annotation. Training data per organ comprised 100 manually annotated volumes, split 80/20 between training and validation.

### *Data Augmentation and Training Details*

### Elastic deformation was the principal augmentation used to improve generalisation and robustness to anatomical variability: a coarse random displacement field, sampled from a Gaussian distribution (σ=0.02) and upsampled to full resolution by trilinear interpolation, was applied jointly to each image and its mask, preserving anatomical plausibility while adding spatial variation. Intensities were normalised channel-wise using pre-computed training-set standard deviations. Rotation and flipping were not used, since the neck-to-knee Dixon acquisition maintains a consistent anatomical orientation across participants. Models were optimised with Adam (initial learning rate 4×10⁻³, exponential decay γ=0.99 per epoch) under FP16 mixed precision to reduce memory use and speed computation. Pre-training ran for up to 150 epochs with early stopping on validation loss; organ-specific fine-tuning typically converged within 30 epochs, with validation performed every half-epoch. Fine-tuning ran for 30 epochs with checkpoints saved at each validation step (twice per epoch), and the checkpoint with the lowest validation loss was deployed for each organ; ties, matched to two decimal places, were broken in favour of the earliest iteration to limit overfitting and favour parsimony once performance plateaued.

### *Post-Processing and Quality Control*

### Final binary masks were obtained by applying softmax to the two-channel output, taking the foreground probability map, and thresholding at 0.5; connected-component analysis then retained only the largest connected region per structure (the two largest for bilateral structures). Quality control combined systematic manual review of three subsets per organ: the 50 largest masks by volume, the 50 smallest, and a random sample of 50. Segmentation errors identified during review (under- or over-segmentation, or erroneous inclusion of adjacent structures) were documented and added to the training annotations, and models were retrained iteratively until they met visual QC criteria across the full range of anatomical variation in the population.

### *Training Infrastructure*

### Training was performed on a compute node with dual NVIDIA GeForce RTX 4090 GPUs (24 GB each) and an 8-core Intel Xeon W-2145 CPU (3.70 GHz base), using data parallelism across both GPUs (batch size 1 per GPU, effective batch size 2, given the size of the 3D volumes) and BF16 mixed precision to fit models, activations, and gradients in available GPU memory while preserving numerical stability. Both full checkpoints (optimizer state and training metadata) and lightweight weights-only checkpoints were written at each validation step (twice per epoch) and synchronised automatically to Google Cloud Storage via a PyTorch Lightning callback, giving permanent storage and straightforward integration with cloud-based inference. This supported distributed training experiments and deployment at scale across the UK Biobank imaging cohort.

**Image Registration and Mesh Construction**

A critical step in our analysis involved calculating the regional kidney thickness at each vertex of the 3D surface mesh. Preprocessing was applied to ensure that all substructures were spatially contained within the kidney. More specifically, for each kidney, the largest connected component of the cortex segmentation was retained to remove small, spurious surrounding tissues, and the resulting cortex boundary was closed and dilated to define a vessel-containment shell. Voxels associated with vessels lying outside this shell, most often reflecting segmentation leakage into the renal pelvis or perirenal vasculature, were removed, and only vessel voxels within it were retained. The retained vessel mask was then combined with the cortex and medulla segmentations to build two composite label maps, a cortex-vessel map and a medulla-vessel map, which served as the inner reference surfaces for the parenchymal, cortical, and medullary thickness distance calculations described below.

The process for organ template construction follows the procedure in^14^. For the cortex and parenchyma, template surfaces were built by averaging segmentations from 20 participants selected to span the anthropometric range of the cohort, providing a stable, representative reference shape for these comparatively large and consistently segmented compartments. We then constructed 3D surface meshes from the template image and all participants' segmentations using the marching cubes algorithm. For the medulla, whose boundaries are thinner, more variable in shape, and more sensitive to inter-subject averaging artefacts given its smaller volume and closer proximity to the renal vasculature, a single, randomly selected participant meeting strict segmentation quality criteria was instead used as the reference template. This single-subject approach avoided the blurring of fine medullary boundary detail that multi-subject averaging can introduce in small, high-curvature structures.

The registration process we employed has been previously outlined in^14^. In brief, we initially used rigid registration to remove the position and orientation difference between all participant-specific surfaces and template surfaces. We then registered the template to the participant's segmentations via affine and non-rigid registration. The template mesh was then propagated to each participant mesh using the deformation fields obtained from the non-rigid registration. Hence, all surface meshes are parameterised with the same number of vertices (approximately 4,000) ensuring each vertex was anatomically accurate and consistent across all participants while preserving the size and shape information for subsequent analysis. All the steps for the template-to-subject registration were performed using the Image Registration Toolkit (IRTK) (<https://biomedia.doc.ic.ac.uk/software/irtk/>).

To determine the regional kidney thickness at each vertex in the 3D model, parenchymal thickness was defined to be the Euclidean distance between the outer parenchymal surface and the vessel segmentation, cortical thickness the distance between cortex and medulla segmentations, and medullary thickness the distance between medulla and vessel segmentations, for each left and right kidney across the population (Supplementary Figure S1).

**Mass Univariate Regression Analysis**

Associations between vertex-wise thickness and covariates were estimated with a linear model, expressed as follows:

$Y=X\beta+\epsilon$,

where, Y is a $n_{s}$ × $n_{v}$ matrix containing $n_{s}$ subjects from a sample of the population under study and $n_{v}$ is the number of voxels in the mesh, X is the $n_{s}$ × p design matrix of p known covariates (including the intercept) and the relevant variables for each subject. X is related to Y by the vector of the estimated regression coefficients $\hat{\beta}$. Finally $\epsilon$ is a $n_{s}$ × $n_{v}$ matrix which is independent and identically distributed across the subjects and is assumed to be a zero-mean Gaussian process^15^. Mass univariate regression (MUR) analysis was performed using a refined version of the R package *mutools3D**^16^*. For each vertex, the model above was fitted to estimate the regression coefficient β for each covariate of interest, adjusting for the remaining covariates in the design matrix. Vertex-wise significance was assessed using threshold-free cluster enhancement (TFCE), a technique that boosts sensitivity to spatially contiguous signal without requiring an arbitrary cluster-forming threshold. TFCE-derived p-values were obtained empirically via permutation testing, with the covariate of interest permuted across subjects 100 times to build a null distribution against which the observed statistic at each vertex was compared. These vertex-wise p-values were then corrected for multiple comparisons across all vertices and models using the Benjamini-Hochberg false discovery rate (FDR) procedure^14,17^.

The resulting regression coefficients $\hat{\beta}$, together with their TFCE-derived, FDR-corrected p-values, were mapped at each vertex onto the 3D kidney mesh, providing spatially-resolved associations across the kidney anatomy. The 3D mesh construction and phenotype mapping pipeline for kidney thickness is outlined in Supplementary Figure S1.

Covariates for the mass univariate regression models were selected based on prior evidence of association with kidney structure, function, or measurement accuracy. Age, sex, ethnicity, BMI, and WHR are established determinants of kidney size and structure^18^. Alcohol intake^19^, smoking status^20^, and fluid intake^21^ were included for their known effects on renal haemodynamics and hydration status. Dominant hand grip strength^22^ and mid-thigh IMAT were included as proxies for skeletal muscle mass, since muscle mass influences creatinine-based renal function estimates and has itself been linked to kidney structural and functional change^23,24^. NSAIDs taken for pain treatment were included given their recognised nephrotoxic potential, particularly affecting the renal medulla^25^. Urinary sodium and urea^26^, and the AST:ALT ratio^27^, were included as markers of renal tubular handling, hydration, and hepatorenal status, respectively. CKD, T2D, and hypertension were included as the primary diseases of interest.

**Statistical Shape Analysis**

Statistical shape analysis (SSA) is a technique defined by the variation of the size and shape (represented using a surface mesh) across participants. Principal component analysis (PCA) is a widely explored dimensionality reduction approach applied in SSA for constructing 3D statistical shape models^28,29^. The principal directions of variation, also called modes of variation, could be represented by eigenvectors calculated from PCA. This technique finds a new coordinate system that describes the input data so that the greatest variance of the data lies on the first coordinate (the first principal component mode), the second-largest variance of the data that is orthogonal to the first component mode on the second coordinate, and so on. In this study, we initially performed PCA on the coordinate system and the thicknesses of the full cohort (N=70,270), to explore whether these coefficients for the principal components known as shape parameters can characterise variations in kidney thicknesses across the population.

Supplementary Tables

| **Trait** | **ICD-10** | **OPCS-4** | **Self-Reported Code (20002)** | **Operation Code**  **(20004)** | **UK Biobank Field** |
| --- | --- | --- | --- | --- | --- |
| **CKD** | N18 | M01, M011-M015,  M018-M019, X401-X406, X408-X409 | 1193 | 1195, 1580, 1581, 1582 | - |
| **T2D** | E11 | - | 1220, 1223 | - | - |
| **Hypertension** | I10-I13, I15, O10 | - | 1065, 1072 | - | 6153, 6177 |
| **CVD** | I00-I25, I30-I52 |  |  |  |  |
| **All-cause mortality** |  |  |  |  | 40020, 40000 |

**Table S1.** Summary of the codes used to define disease.

|  | **Full Cohort**  (N = 70,270) | **Men**  (N = 34,044) | **Women**  (N = 36,226) |
| --- | --- | --- | --- |
| **Demographics** | | | |
| **Age (years)** | 66.20 ± 7.97 | 66.81 ± 8.04 | 65.63 ± 7.87 |
| **White Ethnicity** | 60,071 (85%) | 29,292 (86%) | 30,779 (85%) |
| **Weight (kg)** | 75.5 ± 15.1 | 83.1 ± 13.3 | 68.4 ± 13.1 |
| **Height (m)** | 1.70 ± 0.09 | 1.77 ± 0.07 | 1.63 ± 0.07 |
| **BMI (kg/m²)** | 26.56 ± 4.43 | 26.99 ± 3.92 | 26.16 ± 4.82 |
| **WHR** | 0.89 ± 0.09 | 0.95 ± 0.07 | 0.83 ± 0.07 |
| **Daily alcohol intake frequency** | 11,950 (17%) | 7,068 (21%) | 4,882 (13%) |
| **Current smoker** | 2,218 (3.2%) | 1,270 (3.7%) | 948 (2.6%) |
| **Daily fluid intake (cups/day)** | 7.72 ± 3.09 | 7.53 ± 3.17 | 7.89 ± 3.01 |
| **Sodium in urine (mmol/L)** | 74.18 ± 42.75 | 84.91 ± 44.10 | 64.10 ± 38.82 |
| **Urea (mmol/L)** | 5.30 ± 1.21 | 5.52 ± 1.21 | 5.08 ± 1.18 |
| **eGFR (mL/min/1.73m²)** | 97.95 ± 12.62 | 97.13 ± 12.48 | 98.73 ± 12.71 |
| **AST/ALT ratio** | 1.26 ± 0.42 | 1.15 ± 0.39 | 1.37 ± 0.43 |
| **NSAIDs for Pain Treatment** | 24,342 (35%) | 12,092 (36%) | 12,250 (34%) |
| **Townsend deprivation index** | -1.90 ± 2.73 | -1.94 ± 2.72 | -1.86 ± 2.74 |
| **Mid-thigh internal fat volume (mL)** | 110.73 ± 78.15 | 119.07 ± 87.23 | 102.90 ± 67.60 |
| **Dominant hand grip strength (kg)** | 30.05 ± 10.73 | 37.45 ± 9.36 | 23.09 ± 6.41 |
| **CKD** | 1,773 (2.5%) | 902 (2.6%) | 871 (2.4%) |
| **T2D** | 3,972 (5.7%) | 2,587 (7.6%) | 1,385 (3.8%) |
| **Hypertension** | 27,838 (40%) | 15,769 (46%) | 12,069 (33%) |
| **Kidney Characteristics** | | | |
| **Left kidney cortex volume (mL)** | 117.34 ± 26.35 | 133.94 ± 23.86 | 101.75 ± 17.59 |
| **Right kidney cortex volume (mL)** | 116.10 ± 24.89 | 131.27 ± 22.30 | 101.85 ± 17.76 |
| **Left kidney medulla volume (mL)** | 56.15 ± 12.71 | 60.49 ± 12.98 | 52.06 ± 10.98 |
| **Right kidney medulla volume (mL)** | 43.71 ± 10.82 | 47.77 ± 11.14 | 39.91 ± 8.96 |
| **Left kidney parenchyma volume (mL)** | 173.49 ± 37.08 | 194.43 ± 34.75 | 153.81 ± 27.07 |
| **Right kidney parenchyma volume (mL)** | 159.82 ± 34.35 | 179.04 ± 32.01 | 141.75 ± 25.53 |
| **Left perirenal fat (mL)** | 156.13 ± 53.87 | 188.55 ± 40.68 | 125.66 ± 46.45 |
| **Right perirenal fat (mL)** | 134.14 ± 52.15 | 166.57 ± 42.73 | 103.66 ± 40.52 |
| **Left renal sinus fat (mL)** | 8.09 ± 6.03 | 10.49 ± 6.34 | 5.85 ± 4.72 |
| **Right renal sinus fat (mL)** | 6.13 ± 5.99 | 8.61 ± 6.42 | 3.79 ± 4.42 |
| **Left perirenal fat ratio** | 0.74 ± 0.21 | 0.84 ± 0.15 | 0.64 ± 0.21 |
| **Right perirenal fat ratio** | 0.64 ± 0.21 | 0.76 ± 0.16 | 0.53 ± 0.18 |

**Table S2.** Summary statistics (mean ± standard deviation) of the participants demographics and kidney characteristics for continuous variables and counts for discrete variables in the full cohort (N=70,270).

|  | **Parenchyma** | | **Cortex** | | **Medulla** | |
| --- | --- | --- | --- | --- | --- | --- |
|  | $\hat{\beta}<0$ | $\hat{\beta}\boldsymbol{>0}$ | $\hat{\beta}<0$ | $\hat{\beta}\boldsymbol{>0}$ | $\hat{\beta}<0$ | $\hat{\beta}\boldsymbol{>0}$ |
|  | Median beta coefficients (IQR, Significance area (%)) | | | | | |
| **Left Kidney Thicknesses (mm)** | | | | | | |
| **CKD** | -0.27 (0.27, 67.93%) | 0.19 (0.11, 12.89%) | -0.11 (0.08, 32.09%) | 0.11 (0.10, 30.18%) | -0.37 (0.25, 87.69%) | 0.18 (0.09, 2.39%) |
| **T2D** | - | 0.19 (0.16, 89%) | -0.05 (0.02, 0.18%) | 0.13 (0.10, 84.39%) | -0.12 (0.02, 1.25%) | 0.16 (0.09, 83.98%) |
| **Hypertension** | -0.08 (0.08, 36.66%) | 0.05 (0.05, 41.94%) | - | 0.08 (0.05, 94.84%) | -0.07 (0.05, 65.05%) | 0.05 (0.02, 8.6%) |
| **Right Kidney Thicknesses (mm)** | | | | | | |
| **CKD** | -0.40 (0.30, 70.57%) | 0.25 (0.05, 1.93%) | -0.11 (0.07, 20.43%) | 0.15 (0.09, 30.7%) | -0.07 (0.12, 72.78%) | 0.03 (0.01, 2.21%) |
| **T2D** | -0.45 (0.35, 11.12%) | 0.60 (0.40, 69.87%) | -0.09 (0.02, 0.14%) | 0.18 (0.13, 96.35%) | -0.01 (0.01, 0.91%) | 0.02 (0.02, 77.86%) |
| **Hypertension** | -0.20 (0.10, 13.47%) | 0.25 (0.20, 71.37%) | - | 0.09 (0.05, 99.86%) | -0.01 (0.005, 11%) | 0.01 (0.01, 67.08%) |

**Table S3.** Significance areas for covariates of the MUR models for the disease states of the model for the left and right kidney (N=70,270). The total area has been split into areas of positive and negative associations. The unstandardised (raw) regression coefficients ($\hat{\beta}$) are presented as median (interquartile range - IQR, Significance area (%)) across all vertices of the left and right kidney surfaces for the vertices with statistically significant associations. The unstandardised (raw) regression coefficients are based on the standardised coefficients and the median standard deviation of the thicknesses across all vertices (SD for the thicknesses is 2.74 mm (kidney left parenchyma), 5.00 mm (kidney right parenchyma), 1.61 mm (kidney left cortex), 1.82 mm (kidney right cortex), 2.3 mm (kidney left medulla) and 0.46 mm (kidney right medulla)).

| **Kidney Left** | **Parenchyma** | | **Cortex** | | **Medulla** | |
| --- | --- | --- | --- | --- | --- | --- |
|  | $\hat{\beta}<0$ | $\hat{\beta}\boldsymbol{>0}$ | $\hat{\beta}<0$ | $\hat{\beta}\boldsymbol{>0}$ | $\hat{\beta}<0$ | $\hat{\beta}\boldsymbol{>0}$ |
|  | Median beta coefficients (IQR, Significance area (%)) | | | | | |
| **All** | | | | | | |
| **Renal Sinus Fat Left** | -0.15 (0.09, 98.41%) | 0.03 (0.04, 1.01%) | -0.05 (0.05, 20.75%) | 0.08 (0.06, 77.16%) | -0.12 (0.11, 94.82%) | 0.04 (0.04, 2.43%) |
| **Renal Sinus Fat Left * CKD** | -0.06 (0.05, 87.51%) | - | -0.04 (0.03, 49.15%) | 0.06 (0.06, 32.11%) | -0.06 (0.03, 67.81%) | 0.05 (0.01, 6.47%) |
| **Renal Sinus Fat Left * T2D** | -0.03 (0.01, 7.92%) | 0.05 (0.04, 62.31%) | -0.04 (0.03, 51.37%) | 0.03 (0.02, 31.25%) | -0.03 (0.01, 12.57%) | 0.03 (0.03, 67.14%) |
| **Renal Sinus Fat Left * Hypertension** | -0.02 (0.02, 15.85%) | 0.04 (0.04, 73.09%) | -0.02 (0.02, 41.38%) | 0.03 (0.02, 48.85%) | -0.02 (0.01, 26.28%) | 0.04 (0.03, 52.96%) |
| **Men** | | | | | | |
| **Renal Sinus Fat Left** | -0.12 (0.08, 93.19%) | 0.04 (0.03, 4.63%) | -0.05 (0.05, 18.33%) | 0.10 (0.07, 75.26%) | -0.13 (0.13, 49.54%) | 0.02 (0.02, 1.62%) |
| **Renal Sinus Fat Left * CKD** | -0.07 (0.05, 80.3%) | 0.01 (0.00, 0.02%) | -0.07 (0.04, 44.12%) | 0.08 (0.06, 26.87%) | -0.06 (0.03, 17.53%) | 0.01 (0.01, 0.15%) |
| **Renal Sinus Fat Left * T2D** | -0.04 (0.02, 7.39%) | 0.05 (0.03, 44.12%) | -0.04 (0.03, 40.27%) | 0.04 (0.02, 17.3%) | -0.03 (0.01, 7.31%) | 0.04 (0.03, 23.3%) |
| **Renal Sinus Fat Left * Hypertension** | -0.03 (0.01, 13.84%) | 0.03 (0.03, 60.19%) | -0.03 (0.03, 48.43%) | 0.03 (0.02, 19.52%) | -0.02 (0.01, 10.58%) | 0.04 (0.02, 25.73%) |
| **Women** | | | | | | |
| **Renal Sinus Fat Left** | -0.15 (0.11, 99.09%) | 0.04 (0.03, 0.64%) | -0.05 (0.04, 20.93%) | 0.08 (0.06, 74.29%) | -0.14 (0.11, 52.63%) | 0.05 (0.02, 1.1%) |
| **Renal Sinus Fat Left * CKD** | -0.05 (0.03, 56.99%) | 0.06 (0.02, 3.61%) | -0.07 (0.03, 6.33%) | 0.06 (0.05, 23.05%) | -0.06 (0.03, 27.38%) | 0.01 (0.00, 0.07%) |
| **Renal Sinus Fat Left * T2D** | -0.05 (0.02, 0.68%) | 0.06 (0.04, 67.51%) | -0.05 (0.03, 39.36%) | 0.05 (0.03, 31.35%) | -0.02 (0.00, 0.11%) | 0.04 (0.03, 36.05%) |
| **Renal Sinus Fat Left * Hypertension** | -0.03 (0.01, 8.08%) | 0.03 (0.03, 67.18%) | -0.03 (0.02, 30.48%) | 0.03 (0.02, 41.34%) | -0.02 (0.01, 6.32%) | 0.03 (0.03, 24.88%) |

**Table S4.** Significance areas for covariates in the MUR model for the anthropometric variables for the full cohort (N=70,270), men (N=34,044) and women (N=36,226) of the model for the left kidney substructures (parenchyma, cortex and medulla) showing the local rate of change as a function of the left renal sinus fat for kidney left thicknesses and their interactions with CKD, T2D and hypertension. The total area has been split into areas of positive, negative associations. The standardised regression coefficients ($\hat{\beta}$) are presented as median (interquartile range - IQR) across all vertices of the right kidney substructure’s surface and the significance areas as a percentage (%) of the vertices with statistically significant associations.

| **Kidney Right** | **Parenchyma** | | **Cortex** | | **Medulla** | |
| --- | --- | --- | --- | --- | --- | --- |
|  | $\hat{\beta}<0$ | $\hat{\beta}\boldsymbol{>0}$ | $\hat{\beta}<0$ | $\hat{\beta}\boldsymbol{>0}$ | $\hat{\beta}<0$ | $\hat{\beta}\boldsymbol{>0}$ |
|  | Median beta coefficients (IQR, Significance area (%)) | | | | | |
| **All** | | | | | | |
| **Renal Sinus Fat Right** | -0.08 (0.07, 47.81%) | 0.06 (0.07, 47.37%) | -0.04 (0.04, 14.27%) | 0.08 (0.07, 80.13%) | -0.05 (0.05, 25.52%) | 0.07 (0.08, 70.16%) |
| **Renal Sinus Fat Right * CKD** | -0.05 (0.03, 29.02%) | 0.07 (0.03, 47.91%) | -0.04 (0.04, 20.45%) | 0.05 (0.04, 42.97%) | -0.04 (0.02, 14.12%) | 0.05 (0.03, 41.45%) |
| **Renal Sinus Fat Right * T2D** | -0.03 (0.02, 31.09%) | 0.04 (0.03, 42.9%) | -0.05 (0.03, 69.98%) | 0.03 (0.03, 19.32%) | -0.03 (0.01, 5.95%) | 0.03 (0.02, 52.81%) |
| **Renal Sinus Fat Right * Hypertension** | -0.02 (0.02, 26.8%) | 0.04 (0.03, 56.28%) | -0.03 (0.03, 59.58%) | 0.03 (0.03, 22.54%) | -0.02 (0.01, 9.47%) | 0.02 (0.02, 57.97%) |
| **Men** | | | | | | |
| **Renal Sinus Fat Right** | -0.10 (0.09, 47.95%) | 0.06 (0.08, 44.4%) | -0.04 (0.04, 12.31%) | 0.09 (0.08, 81.93%) | -0.06 (0.04, 23.74%) | 0.08 (0.08, 70.13%) |
| **Renal Sinus Fat Right * CKD** | -0.06 (0.03, 11.05%) | 0.08 (0.05, 42.53%) | -0.07 (0.02, 8.22%) | 0.08 (0.05, 35.87%) | -0.06 (0.03, 3.63%) | 0.06 (0.04, 44.46%) |
| **Renal Sinus Fat Right * T2D** | -0.04 (0.02, 7.75%) | 0.04 (0.02, 37.31%) | -0.05 (0.03, 65.85%) | 0.04 (0.02, 10.09%) | -0.04 (0.01, 6.03%) | 0.04 (0.02, 39.24%) |
| **Renal Sinus Fat Right * Hypertension** | -0.03 (0.02, 8.43%) | 0.04 (0.03, 62.74%) | -0.04 (0.03, 53.55%) | 0.03 (0.02, 16.45%) | -0.02 (0.01, 3.74%) | 0.02 (0.02, 40.8%) |
| **Women** | | | | | | |
| **Renal Sinus Fat Right** | -0.05 (0.06, 44.73%) | 0.06 (0.05, 46.68%) | -0.03 (0.04, 16.18%) | 0.06 (0.06, 74.63%) | -0.05 (0.04, 27.95%) | 0.05 (0.06, 63.67%) |
| **Renal Sinus Fat Right * CKD** | -0.06 (0.03, 10.68%) | 0.08 (0.03, 21.1%) | -0.05 (0.02, 24.1%) | 0.05 (0.02, 2.75%) | -0.05 (0.01, 2.07%) | 0.05 (0.03, 14.56%) |
| **Renal Sinus Fat Right * T2D** | -0.05 (0.02, 24.73%) | 0.05 (0.03, 17.9%) | -0.05 (0.03, 46.66%) | 0.04 (0.02, 16.78%) | -0.04 (0.02, 2.61%) | 0.04 (0.02, 36.88%) |
| **Renal Sinus Fat Right * Hypertension** | -0.03 (0.03, 18.38%) | 0.04 (0.03, 49.02%) | -0.03 (0.03, 57.18%) | 0.03 (0.02, 9.89%) | -0.02 (0.01, 6.32%) | 0.02 (0.02, 36.44%) |

**Table S5.** Significance areas for covariates in the MUR model for the anthropometric variables for the full cohort (N=70,270), men (N=34,044) and women (N=36,226) of the model for the right kidney substructures (parenchyma, cortex and medulla) showing the local rate of change as a function of the right renal sinus fat for kidney right thicknesses and their interactions with CKD, T2D and hypertension. The total area has been split into areas of positive, negative associations. The standardised regression coefficients ($\hat{\beta}$) are presented as median (interquartile range - IQR) across all vertices of the right kidney substructure’s surface and the significance areas as a percentage (%) of the vertices with statistically significant associations.

| **Kidney Left** | **Parenchyma** | | **Cortex** | | **Medulla** | |
| --- | --- | --- | --- | --- | --- | --- |
|  | $\hat{\beta}<0$ | $\hat{\beta}\boldsymbol{>0}$ | $\hat{\beta}<0$ | $\hat{\beta}\boldsymbol{>0}$ | $\hat{\beta}<0$ | $\hat{\beta}\boldsymbol{>0}$ |
|  | Median beta coefficients (IQR, Significance area (%)) | | | | | |
| **All** | | | | | | |
| **Perirenal Fat Ratio Left** | -0.09 (0.07, 46.23%) | 0.07 (0.07, 49.72%) | -0.09 (0.09, 29.37%) | 0.13 (0.09, 68.05%) | -0.08 (0.09, 52.74%) | 0.05 (0.05, 39.43%) |
| **Perirenal Fat Ratio Left * CKD** | -0.05 (0.03, 70.47%) | 0.04 (0.01, 1.19%) | -0.06 (0.05, 58.2%) | 0.06 (0.03, 9.57%) | -0.06 (0.02, 33.52%) | 0.01 (0.01, 0.33%) |
| **Perirenal Fat Ratio Left * T2D** | -0.06 (0.04, 37.05%) | 0.04 (0.02, 10.27%) | -0.05 (0.04, 51.81%) | 0.05 (0.02, 18.15%) | -0.06 (0.03, 16.72%) | 0.05 (0.02, 11.94%) |
| **Perirenal Fat Ratio Left * Hypertension** | -0.02 (0.02, 42.04%) | 0.02 (0.01, 23.51%) | -0.02 (0.02, 25.89%) | 0.03 (0.04, 54.49%) | -0.02 (0.02, 58.4%) | 0.02 (0.01, 6.14%) |
| **Men** | | | | | | |
| **Perirenal Fat Ratio Left** | -0.06 (0.05, 43.73%) | 0.06 (0.06, 49.54%) | -0.07 (0.05, 29.29%) | 0.10 (0.11, 66.74%) | -0.05 (0.07, 30.28%) | 0.03 (0.03, 17.35%) |
| **Perirenal Fat Ratio Left * CKD** | -0.10 (0.07, 72.76%) | 0.09 (0.03, 1.59%) | -0.13 (0.13, 73.45%) | 0.09 (0.04, 1.83%) | -0.09 (0.03, 7.5%) | 0.08 (0.03, 9.59%) |
| **Perirenal Fat Ratio Left * T2D** | -0.06 (0.03, 55.74%) | 0.08 (0.02, 0.83%) | -0.08 (0.05, 89.52%) | 0.00 (0.00, 0.02%) | -0.05 (0.02, 1.03%) | 0.06 (0.02, 16.98%) |
| **Perirenal Fat Ratio Left * Hypertension** | -0.03 (0.01, 29.39%) | 0.02 (0.00, 0.95%) | -0.03 (0.02, 55.86%) | 0.02 (0.01, 13.17%) | -0.03 (0.01, 10.14%) | 0.02 (0.01, 3.2%) |
| **Women** | | | | | | |
| **Perirenal Fat Ratio Left** | -0.08 (0.08, 46.7%) | 0.07 (0.08, 48.85%) | -0.08 (0.08, 23.95%) | 0.14 (0.11, 72.93%) | -0.07 (0.10, 34.4%) | 0.06 (0.06, 16.39%) |
| **Perirenal Fat Ratio Left * CKD** | - | - | -0.07 (0.03, 20.47%) | 0.07 (0.04, 29.17%) | -0.07 (0.04, 15.58%) | 0.06 (0.06, 0.77%) |
| **Perirenal Fat Ratio Left * T2D** | -0.07 (0.04, 31.59%) | 0.07 (0.03, 16.52%) | -0.07 (0.05, 1.17%) | 0.08 (0.05, 52.26%) | -0.08 (0.04, 12.53%) | 0.04 (0.06, 0.44%) |
| **Perirenal Fat Ratio Left * Hypertension** | -0.03 (0.02, 48.71%) | 0.03 (0.02, 13.15%) | -0.03 (0.02, 15.39%) | 0.04 (0.04, 55.3%) | -0.03 (0.01, 31.61%) | 0.01 (0.02, 0.33%) |

**Table S6.** Significance areas for covariates in the MUR model for the anthropometric variables for the full cohort (N=70,270), men (N=34,044) and women (N=36,226) of the model for the left kidney substructures (parenchyma, cortex and medulla) showing the local rate of change as a function of the left perirenal fat ratio for kidney left thicknesses and their interactions with CKD, T2D and hypertension. The total area has been split into areas of positive, negative associations. The standardised regression coefficients ($\hat{\beta}$) are presented as median (interquartile range - IQR) across all vertices of the right kidney substructure’s surface and the significance areas as a percentage (%) of the vertices with statistically significant associations.

| **Kidney Right** | **Parenchyma** | | **Cortex** | | **Medulla** | |
| --- | --- | --- | --- | --- | --- | --- |
|  | $\hat{\beta}<0$ | $\hat{\beta}\boldsymbol{>0}$ | $\hat{\beta}<0$ | $\hat{\beta}\boldsymbol{>0}$ | $\hat{\beta}<0$ | $\hat{\beta}\boldsymbol{>0}$ |
|  | Median beta coefficients (IQR, Significance area (%)) | | | | | |
| **All** | | | | | | |
| **Perirenal Fat Ratio Right** | -0.10 (0.10, 46.51%) | 0.06 (0.06, 49.82%) | -0.07 (0.08, 23.71%) | 0.17 (0.17, 74.26%) | -0.10 (0.10, 69.26%) | 0.04 (0.05, 25.59%) |
| **Perirenal Fat Ratio Right * CKD** | -0.06 (0.04, 32.42%) | 0.06 (0.02, 16.3%) | -0.05 (0.03, 31.52%) | 0.05 (0.03, 19.85%) | -0.05 (0.03, 31.54%) | 0.04 (0.03, 23.92%) |
| **Perirenal Fat Ratio Right * T2D** | -0.05 (0.04, 40.79%) | 0.07 (0.05, 28.67%) | -0.05 (0.03, 41.65%) | 0.05 (0.03, 20.61%) | -0.04 (0.03, 16.15%) | 0.05 (0.03, 40.8%) |
| **Perirenal Fat Ratio Right * Hypertension** | -0.04 (0.03, 31.32%) | 0.04 (0.04, 52.34%) | -0.03 (0.03, 41.02%) | 0.02 (0.02, 42.23%) | -0.02 (0.02, 32.96%) | 0.02 (0.02, 54.23%) |
| **Men** | | | | | | |
| **Perirenal Fat Ratio Right** | -0.09 (0.08, 49.86%) | 0.06 (0.04, 42.97%) | -0.05 (0.05, 26.13%) | 0.13 (0.14, 68.87%) | -0.07 (0.08, 67.15%) | 0.03 (0.04, 24.5%) |
| **Perirenal Fat Ratio Right * CKD** | - | - | -0.08 (0.04, 41.3%) | 0.08 (0.03, 3.65%) | - | - |
| **Perirenal Fat Ratio Right * T2D** | -0.06 (0.03, 26.48%) | 0.06 (0.01, 0.18%) | -0.07 (0.04, 49.43%) | 0.06 (0.03, 5.8%) | - | - |
| **Perirenal Fat Ratio Right * Hypertension** | -0.03 (0.02, 28.06%) | 0.04 (0.02, 39.13%) | -0.03 (0.02, 50.9%) | 0.03 (0.03, 18.74%) | -0.03 (0.01, 26.61%) | 0.02 (0.01, 16.48%) |
| **Women** | | | | | | |
| **Perirenal Fat Ratio Right** | -0.09 (0.10, 47.66%) | 0.08 (0.09, 47.35%) | -0.06 (0.08, 19.44%) | 0.18 (0.16, 77.36%) | -0.11 (0.09, 61.63%) | 0.04 (0.05, 31.25%) |
| **Perirenal Fat Ratio Right * CKD** | -0.07 (0.02, 19.22%) | 0.07 (0.02, 7.03%) | -0.06 (0.03, 23.3%) | 0.07 (0.02, 7.55%) | -0.07 (0.03, 12.78%) | 0.06 (0.03, 9.8%) |
| **Perirenal Fat Ratio Right * T2D** | -0.07 (0.03, 15.65%) | 0.08 (0.06, 30.56%) | -0.05 (0.03, 23.61%) | 0.06 (0.04, 39.5%) | -0.07 (0.03, 6.93%) | 0.06 (0.04, 46.32%) |
| **Perirenal Fat Ratio Right * Hypertension** | -0.04 (0.05, 22.03%) | 0.04 (0.04, 59.11%) | -0.03 (0.03, 45.75%) | 0.03 (0.02, 28.51%) | -0.03 (0.01, 15.75%) | 0.03 (0.02, 48.71%) |

**Table S7.** Significance areas for covariates in the MUR model for the anthropometric variables for the full cohort (N=70,270), men (N=34,044) and women (N=36,226) of the model for the right kidney substructures (parenchyma, cortex and medulla) showing the local rate of change as a function of the right perirenal fat ratio for kidney right thicknesses and their interactions with CKD, T2D and hypertension. The total area has been split into areas of positive, negative associations. The standardised regression coefficients ($\hat{\beta}$) are presented as median (interquartile range - IQR) across all vertices of the right kidney substructure’s surface and the significance areas as a percentage (%) of the vertices with statistically significant associations.

| **Kidney Left** | **Parenchyma** | | **Cortex** | | **Medulla** | |
| --- | --- | --- | --- | --- | --- | --- |
|  | $\hat{\beta}<0$ | $\hat{\beta}\boldsymbol{>0}$ | $\hat{\beta}<0$ | $\hat{\beta}\boldsymbol{>0}$ | $\hat{\beta}<0$ | $\hat{\beta}\boldsymbol{>0}$ |
|  | Median beta coefficients (IQR, Significance area (%)) | | | | | |
| **Men** | | | | | | |
| **Age** | -0.04 (0.05, 78.83%) | 0.02 (0.01, 2.58%) | -0.02 (0.02, 34.71%) | 0.02 (0.02, 41.06%) | -0.06 (0.05, 87.87%) | 0.03 (0.01, 4.48%) |
| **BMI** | -0.02 (0.01, 2.42%) | 0.10 (0.10, 88.15%) | -0.03 (0.01, 0.26%) | 0.22 (0.11, 99.11%) | -0.04 (0.04, 21.83%) | 0.05 (0.06, 66.12%) |
| **WHR** | -0.03 (0.02, 35.58%) | 0.03 (0.02, 40.53%) | -0.03 (0.03, 7.94%) | 0.08 (0.07, 81.73%) | -0.04 (0.03, 78.39%) | 0.03 (0.02, 7.68%) |
| **Alcohol intake** | -0.04 (0.02, 12.87%) | 0.03 (0.02, 37.09%) | -0.01 (0.00, 0.1%) | 0.06 (0.03, 98.17%) | -0.03 (0.02, 40.83%) | 0.03 (0.01, 16.8%) |
| **Smoking status** | -0.06 (0.02, 2.4%) | 0.06 (0.04, 64.83%) | -0.07 (0.02, 2.05%) | 0.07 (0.05, 59.02%) | 0.00 (0.00, 0.04%) | 0.07 (0.04, 70.49%) |
| **Fluid intake** | -0.01 (0.00, 0.66%) | 0.01 (0.01, 74.84%) | -0.01 (0.01, 0.46%) | 0.01 (0.01, 34.67%) | -0.01 (0.01, 1.43%) | 0.02 (0.01, 83.2%) |
| **Sodium in urine** | -0.01 (0.01, 19.86%) | 0.01 (0.01, 15.39%) | -0.01 (0.00, 7.74%) | 0.01 (0.01, 39.99%) | -0.01 (0.00, 10.18%) | 0.01 (0.01, 25.28%) |
| **Urea** | -0.01 (0.01, 75.04%) | 0.01 (0.01, 2.3%) | -0.02 (0.01, 77.46%) | 0.01 (0.00, 1.11%) | -0.02 (0.02, 83.94%) | 0.02 (0.01, 2.43%) |
| **AST:ALT** | -0.02 (0.01, 58.72%) | 0.01 (0.01, 10.37%) | -0.02 (0.02, 71.92%) | 0.02 (0.01, 4.43%) | -0.01 (0.01, 36.09%) | 0.01 (0.00, 5.92%) |
| **NSAIDs for Pain** | -0.02 (0.01, 0.83%) | 0.02 (0.01, 28.93%) | - | - | 0.00 (0.01, 0.11%) | 0.02 (0.01, 32.08%) |
| **Mid-Thigh IMAT** | -0.02 (0.02, 39.93%) | 0.02 (0.01, 14.1%) | -0.02 (0.02, 61.58%) | 0.02 (0.02, 16.52%) | -0.02 (0.01, 47.41%) | 0.02 (0.01, 22.2%) |
| **Dominant HGS** | -0.01 (0.01, 25.36%) | 0.02 (0.01, 37.43%) | -0.02 (0.01, 6.1%) | 0.02 (0.01, 61.81%) | -0.02 (0.02, 7.57%) | 0.02 (0.02, 69.68%) |
| **CKD** | -0.12 (0.09, 82.8%) | 0.05 (0.05, 0.81%) | -0.10 (0.06, 28.12%) | 0.09 (0.07, 17.47%) | -0.17 (0.11, 89.6%) | 0.11 (0.04, 0.66%) |
| **T2D** | - | 0.09 (0.06, 86.56%) | -0.01 (0.00, 0.1%) | 0.08 (0.06, 76.63%) | -0.06 (0.01, 0.48%) | 0.08 (0.06, 84.9%) |
| **Hypertension** | -0.04 (0.03, 28.61%) | 0.03 (0.02, 49.13%) | -0.03 (0.02, 0.68%) | 0.05 (0.04, 88.17%) | -0.03 (0.02, 52.66%) | 0.02 (0.01, 15.62%) |
| **Age*CKD** | -0.08 (0.03, 16.72%) | 0.07 (0.03, 25.89%) | -0.07 (0.05, 0.46%) | 0.07 (0.03, 23.21%) | - | - |
| **Age*T2D** | -0.05 (0.03, 75.73%) | - | -0.04 (0.02, 48.11%) | 0.04 (0.01, 3.51%) | -0.04 (0.03, 67.14%) | - |
| **Age*Hypertension** | -0.02 (0.01, 84.29%) | - | -0.02 (0.02, 40.41%) | 0.02 (0.01, 7.66%) | -0.02 (0.02, 72.84%) | 0.02 (0.01, 0.15%) |
| **BMI*T2D** | - | - | - | - | - | - |
| **WHR*T2D** | -0.04 (0.02, 24.05%) | 0.04 (0.02, 18.71%) | -0.05 (0.04, 47.93%) | 0.05 (0.03, 24.29%) | -0.04 (0.03, 35.21%) | 0.05 (0.02, 14.59%) |
| **Women** | | | | | | |
| **Age** | -0.04 (0.06, 69.64%) | 0.02 (0.02, 15.25%) | -0.03 (0.03, 39.08%) | 0.03 (0.02, 42.26%) | -0.06 (0.06, 74.72%) | 0.02 (0.01, 9.52%) |
| **BMI** | -0.03 (0.03, 18.96%) | 0.07 (0.06, 75.77%) | -0.04 (0.03, 12.49%) | 0.17 (0.09, 85.07%) | -0.03 (0.03, 15.62%) | 0.04 (0.03, 76.3%) |
| **WHR** | -0.02 (0.01, 21.09%) | 0.03 (0.03, 50.6%) | -0.02 (0.01, 8.98%) | 0.08 (0.05, 83.98%) | -0.02 (0.02, 48.59%) | 0.02 (0.01, 20.73%) |
| **Alcohol intake** | -0.04 (0.03, 39.44%) | 0.03 (0.01, 8.5%) | -0.03 (0.02, 1.59%) | 0.05 (0.04, 87.17%) | -0.04 (0.02, 33.33%) | 0.04 (0.02, 16.8%) |
| **Smoking status** | -0.07 (0.03, 5.88%) | 0.06 (0.03, 44.26%) | -0.06 (0.02, 1.29%) | 0.09 (0.06, 88.07%) | -0.07 (0.02, 3.9%) | 0.07 (0.04, 43.22%) |
| **Fluid intake** | -0.01 (0.01, 13.72%) | 0.02 (0.01, 40.05%) | -0.01 (0.00, 14%) | 0.01 (0.01, 39.46%) | -0.01 (0.01, 7.79%) | 0.02 (0.01, 66.74%) |
| **Sodium in urine** | -0.01 (0.01, 25.16%) | 0.01 (0.01, 7.21%) | -0.01 (0.00, 3.51%) | 0.01 (0.01, 67.57%) | -0.01 (0.01, 3.27%) | 0.01 (0.01, 15.91%) |
| **Urea** | -0.01 (0.01, 45%) | 0.02 (0.01, 23.77%) | -0.01 (0.01, 59.43%) | 0.01 (0.01, 8.98%) | -0.02 (0.02, 68.83%) | 0.01 (0.01, 9.67%) |
| **AST:ALT** | -0.01 (0.01, 54.53%) | 0.01 (0.01, 16.4%) | -0.02 (0.01, 81.04%) | 0.01 (0.01, 8.22%) | -0.01 (0.01, 56.74%) | 0.01 (0.00, 3.53%) |
| **NSAIDs for Pain** | - | - | - | - | -0.01 (0.02, 1.21%) | 0.02 (0.01, 13.01%) |
| **Mid-Thigh IMAT** | -0.03 (0.02, 28.71%) | 0.02 (0.01, 44.98%) | -0.02 (0.01, 7.64%) | 0.03 (0.03, 66.58%) | -0.02 (0.01, 22.09%) | 0.02 (0.01, 44.21%) |
| **Dominant HGS** | -0.03 (0.02, 49.38%) | 0.03 (0.03, 23.67%) | -0.03 (0.01, 9.33%) | 0.03 (0.02, 68.23%) | -0.02 (0.02, 21.54%) | 0.03 (0.03, 40.06%) |
| **CKD** | -0.12 (0.10, 46.98%) | 0.12 (0.07, 19.88%) | -0.09 (0.04, 20.59%) | 0.10 (0.09, 27.22%) | -0.16 (0.12, 80.6%) | 0.11 (0.06, 3.31%) |
| **T2D** | -0.08 (0.02, 0.95%) | 0.08 (0.05, 55.66%) | -0.06 (0.01, 0.32%) | 0.11 (0.08, 85.44%) | -0.07 (0.02, 2.35%) | 0.07 (0.03, 52.48%) |
| **Hypertension** | -0.04 (0.02, 38.44%) | 0.02 (0.01, 13.15%) | 0.00 (0.00, 0.14%) | 0.05 (0.03, 92.22%) | -0.04 (0.03, 68.76%) | 0.03 (0.01, 2.43%) |
| **Age*CKD** | - | - | -0.07 (0.02, 11.02%) | 0.08 (0.04, 12.57%) | - | - |
| **Age*T2D** | -0.05 (0.02, 29.33%) | 0.06 (0.02, 1.89%) | -0.07 (0.02, 2.36%) | 0.06 (0.03, 19.88%) | -0.06 (0.03, 71.04%) | 0.06 (0.01, 1.58%) |
| **Age*Hypertension** | -0.02 (0.01, 66.58%) | 0.00 (0.00, 0.18%) | -0.03 (0.02, 32.66%) | 0.02 (0.01, 9.83%) | -0.02 (0.01, 53.84%) | 0.02 (0.01, 0.96%) |
| **BMI*T2D** | -0.11 (0.06, 1.31%) | 0.14 (0.06, 19.9%) | -0.12 (0.05, 29.9%) | 0.03 (0.07, 0.6%) | - | - |
| **WHR*T2D** | -0.04 (0.02, 19.72%) | 0.04 (0.03, 26.97%) | -0.04 (0.02, 18.07%) | 0.05 (0.04, 45.71%) | -0.04 (0.02, 18.3%) | 0.05 (0.03, 12.24%) |

**Table S8.** Significance areas for covariates in the MUR model for the anthropometric variables for men (N=34,044) and women (N=36,226) of the model for the left kidney substructures (parenchyma, cortex and medulla). The total area has been split into areas of positive, negative associations. The standardised regression coefficients ($\hat{\beta}$) are presented as median (interquartile range - IQR) across all vertices of the left kidney substructure’s surface and the significance areas as a percentage (%) of the vertices with statistically significant associations.

| **Kidney Right** | **Parenchyma** | | **Cortex** | | **Medulla** | |
| --- | --- | --- | --- | --- | --- | --- |
|  | $\hat{\beta}<0$ | $\hat{\beta}\boldsymbol{>0}$ | $\hat{\beta}<0$ | $\hat{\beta}\boldsymbol{>0}$ | $\hat{\beta}<0$ | $\hat{\beta}\boldsymbol{>0}$ |
|  | Median beta coefficients (IQR, Significance area (%)) | | | | | |
| **Men** | | | | | | |
| **Age** | -0.03 (0.03, 82.01%) | 0.02 (0.01, 7.12%) | -0.02 (0.02, 5.76%) | 0.03 (0.03, 79%) | -0.08 (0.07, 51%) | 0.03 (0.02, 34.99%) |
| **BMI** | -0.02 (0.03, 46.68%) | 0.02 (0.02, 44.87%) | -0.03 (0.03, 3.18%) | 0.12 (0.09, 92.8%) | -0.02 (0.01, 0.94%) | 0.07 (0.06, 95.14%) |
| **WHR** | -0.01 (0.02, 43.99%) | 0.01 (0.01, 34%) | -0.03 (0.01, 5.72%) | 0.05 (0.05, 83%) | -0.03 (0.02, 48.02%) | 0.02 (0.02, 32.92%) |
| **Alcohol intake** | -0.07 (0.08, 10.01%) | 0.07 (0.06, 53.12%) | -0.03 (0.00, 0.14%) | 0.04 (0.03, 83.55%) | -0.02 (0.01, 3.16%) | 0.03 (0.02, 45.26%) |
| **Smoking status** | -0.01 (0.01, 56.38%) | 0.01 (0.01, 9.02%) | -0.06 (0.06, 10.73%) | 0.08 (0.05, 71.12%) | -0.05 (0.02, 2.61%) | 0.05 (0.04, 68.13%) |
| **Fluid intake** | 0.00 (0.00, 22.81%) | 0.00 (0.01, 12.2%) | -0.01 (0.00, 0.21%) | 0.01 (0.01, 72.01%) | -0.01 (0.00, 3.77%) | 0.02 (0.02, 60.4%) |
| **Sodium in urine** | -0.02 (0.01, 24.63%) | 0.02 (0.02, 25.47%) | -0.01 (0.00, 10.85%) | 0.01 (0.00, 11.36%) | -0.01 (0.01, 13.03%) | 0.01 (0.01, 36.23%) |
| **Urea** | -0.01 (0.01, 22.21%) | 0.01 (0.01, 32.2%) | -0.01 (0.01, 50.64%) | 0.01 (0.00, 1.99%) | -0.02 (0.04, 72.05%) | 0.01 (0.01, 2.94%) |
| **AST:ALT** | 0.00 (0.00, 26.17%) | 0.01 (0.01, 44.42%) | -0.02 (0.02, 77.91%) | 0.02 (0.00, 0.29%) | -0.01 (0.01, 61.96%) | 0.01 (0.00, 2.79%) |
| **NSAIDs for Pain** | -0.07 (0.05, 12.84%) | 0.16 (0.09, 41.32%) | -0.01 (0.00, 0.27%) | 0.02 (0.01, 20.02%) | -0.02 (0.01, 0.69%) | 0.03 (0.02, 41.85%) |
| **Mid-Thigh IMAT** | -0.01 (0.01, 23.65%) | 0.02 (0.02, 47.05%) | -0.02 (0.01, 49.2%) | 0.02 (0.01, 11.67%) | -0.02 (0.02, 42.18%) | 0.02 (0.02, 29.04%) |
| **Dominant HGS** | -0.01 (0.02, 20.37%) | 0.03 (0.03, 40.03%) | -0.02 (0.01, 20.92%) | 0.02 (0.01, 26.29%) | -0.01 (0.01, 5.7%) | 0.03 (0.05, 76.04%) |
| **CKD** | -0.03 (0.04, 13.66%) | 0.05 (0.04, 47.09%) | -0.12 (0.06, 10.27%) | 0.09 (0.05, 34.35%) | -0.19 (0.28, 68.06%) | 0.08 (0.04, 4.07%) |
| **T2D** | -0.08 (0.09, 66.76%) | 0.03 (0.04, 11.59%) | -0.07 (0.02, 0.21%) | 0.10 (0.06, 91.8%) | -0.05 (0.02, 1.23%) | 0.05 (0.04, 64.54%) |
| **Hypertension** | -0.02 (0.03, 33.2%) | 0.03 (0.04, 44.67%) | - | 0.06 (0.03, 99.08%) | -0.02 (0.01, 15.35%) | 0.03 (0.02, 59.6%) |
| **Age*CKD** | - | - | -0.08 (0.02, 1.42%) | 0.07 (0.03, 34.68%) | - | - |
| **Age*T2D** | -0.06 (0.05, 37.35%) | 0.03 (0.05, 10.99%) | -0.04 (0.02, 28.53%) | 0.04 (0.02, 1.7%) | -0.04 (0.02, 39.56%) | 0.04 (0.02, 4.54%) |
| **Age*Hypertension** | -0.02 (0.01, 27.63%) | 0.02 (0.01, 15.46%) | -0.02 (0.01, 26%) | 0.02 (0.01, 7.79%) | -0.02 (0.01, 35.28%) | 0.02 (0.01, 16.15%) |
| **BMI*T2D** | - | - | - | - | - | - |
| **WHR*T2D** | -0.02 (0.02, 26.74%) | 0.02 (0.02, 22.97%) | -0.09 (0.06, 66.94%) | 0.05 (0.03, 13.99%) | -0.04 (0.02, 0.58%) | 0.04 (0.02, 48.64%) |
| **Women** | | | | | | |
| **Age** | -0.05 (0.04, 84.6%) | 0.02 (0.01, 4.72%) | -0.03 (0.03, 22.35%) | 0.03 (0.03, 63.52%) | -0.08 (0.07, 56.81%) | 0.03 (0.03, 30.2%) |
| **BMI** | -0.05 (0.06, 10.58%) | 0.09 (0.07, 87.28%) | -0.03 (0.03, 18.87%) | 0.13 (0.10, 77.42%) | -0.02 (0.02, 5.74%) | 0.05 (0.05, 86.24%) |
| **WHR** | -0.03 (0.02, 35.09%) | 0.03 (0.03, 46.82%) | -0.02 (0.02, 9.33%) | 0.06 (0.06, 77.83%) | -0.03 (0.02, 26.75%) | 0.02 (0.02, 49.84%) |
| **Alcohol intake** | -0.03 (0.02, 9.47%) | 0.05 (0.04, 45.37%) | -0.03 (0.02, 3.86%) | 0.07 (0.05, 71.94%) | -0.03 (0.01, 2.5%) | 0.03 (0.03, 55.68%) |
| **Smoking status** | -0.07 (0.01, 2.09%) | 0.08 (0.04, 64.46%) | -0.06 (0.01, 0.55%) | 0.09 (0.07, 75.86%) | -0.05 (0.03, 0.98%) | 0.06 (0.04, 49.4%) |
| **Fluid intake** | -0.01 (0.01, 0.62%) | 0.01 (0.01, 56.32%) | -0.01 (0.01, 9.78%) | 0.01 (0.01, 57.98%) | -0.01 (0.00, 3.92%) | 0.03 (0.03, 66.1%) |
| **Sodium in urine** | - | - | -0.01 (0.01, 11.14%) | 0.01 (0.01, 33.49%) | -0.01 (0.01, 38.22%) | 0.01 (0.01, 17.13%) |
| **Urea** | -0.02 (0.01, 86.03%) | 0.02 (0.01, 1.89%) | -0.01 (0.01, 39.48%) | 0.01 (0.00, 6.73%) | -0.02 (0.04, 78.73%) | 0.01 (0.00, 1.38%) |
| **AST:ALT** | -0.02 (0.01, 65.42%) | 0.01 (0.01, 6.32%) | -0.02 (0.01, 83.18%) | 0.01 (0.00, 3.75%) | -0.02 (0.02, 62.54%) | 0.01 (0.01, 2.94%) |
| **NSAIDs for Pain** | - | - | -0.02 (0.01, 0.98%) | 0.02 (0.01, 23.89%) | -0.02 (0.01, 0.94%) | 0.02 (0.01, 54.16%) |
| **Mid-Thigh IMAT** | -0.03 (0.02, 40.4%) | 0.02 (0.02, 27.75%) | -0.03 (0.02, 13.23%) | 0.03 (0.02, 73.58%) | -0.02 (0.01, 21.56%) | 0.02 (0.02, 38.8%) |
| **Dominant HGS** | -0.02 (0.01, 18.4%) | 0.04 (0.03, 57.94%) | -0.03 (0.02, 33.92%) | 0.03 (0.02, 35.81%) | -0.02 (0.00, 0.62%) | 0.04 (0.06, 82.72%) |
| **CKD** | -0.10 (0.05, 50.39%) | 0.08 (0.03, 2.15%) | -0.07 (0.03, 27.36%) | 0.10 (0.06, 18.89%) | -0.18 (0.25, 70.78%) | 0.08 (0.03, 2.21%) |
| **T2D** | -0.07 (0.04, 6.87%) | 0.12 (0.07, 79.16%) | -0.02 (0.02, 0.49%) | 0.13 (0.10, 86.92%) | -0.05 (0.06, 0.44%) | 0.08 (0.07, 66.13%) |
| **Hypertension** | -0.03 (0.02, 7.9%) | 0.05 (0.04, 81.36%) | - | 0.05 (0.03, 98.3%) | -0.03 (0.01, 4.57%) | 0.03 (0.02, 66.39%) |
| **Age*CKD** | - | - | - | - | - | - |
| **Age*T2D** | -0.06 (0.03, 26.37%) | 0.07 (0.01, 1.89%) | -0.06 (0.03, 21.86%) | 0.05 (0.02, 0.29%) | - | - |
| **Age*Hypertension** | -0.02 (0.01, 7.47%) | 0.03 (0.02, 36.34%) | -0.02 (0.01, 18.64%) | 0.02 (0.01, 9.13%) | - | - |
| **BMI*T2D** | - | - | -0.14 (0.06, 15.11%) | 0.13 (0.05, 10.62%) | -0.13 (0.07, 36.01%) | 0.13 (0.06, 2.29%) |
| **WHR*T2D** | -0.06 (0.03, 33.55%) | 0.06 (0.03, 23.58%) | -0.07 (0.06, 57.77%) | 0.06 (0.03, 15.59%) | -0.05 (0.02, 4.68%) | 0.05 (0.04, 52.7%) |

**Table S9.** Significance areas for covariates in the MUR model for the anthropometric variables for men (N=34,044) and women (N=36,226) of the model for the right kidney substructures (parenchyma, cortex and medulla). The total area has been split into areas of positive, negative associations. The standardised regression coefficients ($\hat{\beta}$) are presented as median (interquartile range - IQR) across all vertices of the right kidney substructure’s surface and the significance areas as a percentage (%) of the vertices with statistically significant associations.

| **Kidney Left** | **Parenchyma** | | **Cortex** | | **Medulla** | |
| --- | --- | --- | --- | --- | --- | --- |
|  | $\hat{\beta}<0$ | $\hat{\beta}\boldsymbol{>0}$ | $\hat{\beta}<0$ | $\hat{\beta}\boldsymbol{>0}$ | $\hat{\beta}<0$ | $\hat{\beta}\boldsymbol{>0}$ |
|  | Median beta coefficients (IQR, Significance area (%)) | | | | | |
| **All** | | | | | | |
| **Intercept** | - | 11.58 (8.75, 100%) | - | 7.15 (1.51, 100%) | - | 8.11 (3.63, 100%) |
| **Age at baseline** | -0.20 (0.13, 38.64%) | 0.13 (0.04, 0.97%) | -0.10 (0.06, 14.81%) | 0.09 (0.03, 3.49%) | -0.19 (0.16, 52.77%) | 0.13 (0.01, 0.26%) |
| **Age² at baseline** | - | - | - | - | - | - |
| **Sex** | -0.69 (0.51, 22.36%) | 0.50 (0.28, 29.33%) | -0.27 (0.05, 0.32%) | 0.65 (0.34, 91.9%) | -0.71 (0.56, 30.43%) | 0.44 (0.23, 22.45%) |
| **Race [Non-white]** | - | - | - | - | - | - |
| **Assessment Centre [Newcastle]** | - | - | - | - | - | - |
| **BMI** | -0.19 (0.11, 2.3%) | 0.26 (0.14, 47.36%) | -0.08 (0.02, 0.42%) | 0.28 (0.12, 84.99%) | -0.21 (0.08, 4.67%) | 0.18 (0.05, 13.67%) |
| **WHR** | -0.23 (0.07, 1.65%) | 0.18 (0.06, 0.87%) | -0.10 (0.03, 0.56%) | 0.12 (0.06, 59.73%) | - | - |
| **Alcohol intake** | - | - | - | - | - | - |
| **Smoking status** | - | - | - | - | - | - |
| **NSAIDs for Pain** | - | - | - | - | - | - |
| **Dominant HGS** | - | - | - | - | - | - |
| **Townsend deprivation index** | - | - | - | - | - | - |
| **Fluid intake** | - | - | - | - | - | - |
| **Mid-Thigh IMAT** | -0.24 (0.03, 0.4%) | 0.19 (0.05, 2.3%) | -0.11 (0.00, 0.12%) | 0.14 (0.08, 2.05%) | - | - |
| **CKD** | - | - | - | - | -0.96 (0.33, 14.85%) | - |
| **T2D** | - | 0.87 (0.17, 6.02%) | - | 0.53 (0.25, 0.52%) | - | - |
| **Hypertension** | - | - | - | - | - | - |
| **2nd Visit** | -0.25 (0.16, 17.08%) | 0.28 (0.12, 14.81%) | -0.13 (0.05, 11.02%) | 0.16 (0.04, 6.75%) | -0.23 (0.16, 8.16%) | 0.23 (0.06, 3.01%) |
| **CKD * 2nd Visit** | - | - | - | - | - | - |
| **T2D * 2nd Visit** | - | - | -0.42 (0.00, 0.04%) | 0.51 (0.20, 4.71%) | - | - |
| **Hypertension * 2nd Visit** | - | 0.67 (0.16, 0.2%) | - | - | -0.38 (0.07, 1.25%) | - |
| **Men** | | | | | | |
| **Intercept** | - | 11.82 (8.62, 100%) | - | 7.89 (1.52, 100%) | - | 8.00 (3.45, 100%) |
| **Age at baseline** | -0.25 (0.14, 37.67%) | 0.06 (0.00, 0.02%) | -0.14 (0.05, 5.06%) | 0.13 (0.03, 1.69%) | -0.22 (0.17, 57.41%) | 0.21 (0.04, 0.62%) |
| **Age² at baseline** | - | - | - | - | - | - |
| **Race [Non-white]** | - | - | - | - | - | - |
| **Assessment Centre [Newcastle]** | - | - | - | - | - | - |
| **BMI** | -0.31 (0.07, 0.69%) | 0.31 (0.15, 40.71%) | -0.13 (0.01, 0.06%) | 0.32 (0.11, 85.88%) | -0.24 (0.02, 0.33%) | 0.25 (0.07, 5.07%) |
| **WHR** | -0.27 (0.07, 1.71%) | 0.17 (0.00, 0.02%) | -0.11 (0.01, 0.56%) | 0.11 (0.03, 26.85%) | - | - |
| **Alcohol intake** | - | - | - | - | - | - |
| **Smoking status** | - | - | - | - | - | - |
| **NSAIDs for Pain** | - | - | - | - | - | - |
| **Dominant HGS** | - | - | - | - | - | - |
| **Townsend deprivation index** | - | - | - | - | - | - |
| **Fluid intake** | - | - | - | - | - | - |
| **Mid-Thigh IMAT** | -0.41 (0.05, 0.36%) | - | - | - | - | - |
| **CKD** | - | - | - | - | -1.17 (0.34, 16.17%) | - |
| **T2D** | - | 1.04 (0.35, 10.7%) | - | - | - | - |
| **Hypertension** | - | - | - | - | - | - |
| **2nd Visit** | -0.42 (0.19, 3.06%) | 0.44 (0.22, 0.64%) | - | 0.26 (0.05, 1.39%) | -0.30 (0.17, 3.64%) | 0.25 (0.02, 0.18%) |
| **CKD * 2nd Visit** | - | - | - | 1.03 (0.20, 0.3%) | - | - |
| **T2D * 2nd Visit** | - | - | -0.90 (0.02, 0.04%) | - | - | - |
| **Hypertension * 2nd Visit** | - | - | - | - | - | - |
| **Women** | | | | | | |
| **Intercept** | - | 11.54 (8.73, 100%) | - | 7.07 (1.47, 100%) | - | 8.04 (3.67, 100%) |
| **Age at baseline** | -0.23 (0.10, 18.13%) | 0.22 (0.08, 0.87%) | -0.11 (0.08, 12.69%) | 0.10 (0.03, 3.36%) | -0.24 (0.12, 28.34%) | 0.21 (0.03, 1.14%) |
| **Age² at baseline** | - | - | - | - | - | - |
| **Race [Non-white]** | - | - | - | - | - | - |
| **Assessment Centre [Newcastle]** | - | 0.33 (0.07, 0.12%) | -0.21 (0.02, 1.07%) | - | - | - |
| **BMI** | -0.23 (0.18, 4.29%) | 0.32 (0.14, 30.6%) | -0.14 (0.04, 2.12%) | 0.26 (0.13, 79.61%) | -0.27 (0.07, 2.06%) | 0.24 (0.05, 4.74%) |
| **WHR** | -0.20 (0.00, 0.02%) | - | -0.09 (0.03, 0.77%) | 0.10 (0.04, 61.06%) | - | - |
| **Alcohol intake** | - | - | - | - | - | - |
| **Smoking status** | - | 1.54 (0.00, 0.02%) | - | - | - | - |
| **NSAIDs for Pain** | - | - | - | - | - | - |
| **Dominant HGS** | - | - | - | - | - | - |
| **Townsend deprivation index** | - | - | - | - | - | - |
| **Fluid intake** | - | - | - | - | - | - |
| **Mid-Thigh IMAT** | - | - | - | - | - | - |
| **CKD** | - | 3.05 (0.00, 0.02%) | - | 0.90 (0.51, 0.36%) | -1.90 (0.08, 0.07%) | - |
| **T2D** | - | - | - | 0.65 (0.15, 6.69%) | - | - |
| **Hypertension** | - | - | - | - | - | - |
| **2nd Visit** | -0.34 (0.28, 14.79%) | 0.42 (0.15, 13.28%) | -0.18 (0.07, 14.36%) | 0.21 (0.04, 5.32%) | -0.36 (0.15, 5.7%) | 0.30 (0.08, 5.18%) |
| **CKD * 2nd Visit** | - | - | - | - | -1.43 (0.13, 0.26%) | - |
| **T2D * 2nd Visit** | - | - | - | 0.83 (0.40, 27.92%) | - | - |
| **Hypertension * 2nd Visit** | - | - | - | - | - | - |

**Table S10.** Significance areas for covariates in the mass univariate linear mixed-effect models for the anthropometric variables for all (N=2,610), men (N=1,305) and women (N=1,305) of the model for the left kidney substructures (parenchyma, cortex and medulla). The total area has been split into areas of positive, negative associations. The standardised regression coefficients ($\hat{\beta}$) are presented as median (interquartile range - IQR) across all vertices of the left kidney substructure’s surface and the significance areas as a percentage (%) of the vertices with statistically significant associations.

| **Kidney Right** | **Parenchyma** | | **Cortex** | | **Medulla** | |
| --- | --- | --- | --- | --- | --- | --- |
|  | $\hat{\beta}<0$ | $\hat{\beta}\boldsymbol{>0}$ | $\hat{\beta}<0$ | $\hat{\beta}\boldsymbol{>0}$ | $\hat{\beta}<0$ | $\hat{\beta}\boldsymbol{>0}$ |
|  | Median beta coefficients (IQR, Significance area (%)) | | | | | |
| **All** | | | | | | |
| **Intercept** | - | 9.99 (9.70, 100%) | - | 7.87 (2.55, 100%) | - | 1.41 (1.50, 100%) |
| **Age at baseline** | -0.40 (0.30, 55.41%) | 0.12 (0.06, 0.57%) | -0.12 (0.07, 2.03%) | 0.10 (0.05, 27.67%) | -0.13 (0.16, 43.27%) | 0.05 (0.04, 8.53%) |
| **Age² at baseline** | - | - | - | - | - | - |
| **Sex** | -0.75 (0.55, 20.92%) | 2.03 (2.37, 56.21%) | -0.34 (0.19, 4.45%) | 0.61 (0.55, 79.57%) | -0.20 (0.13, 2.9%) | 0.22 (0.34, 60.69%) |
| **Race [Non-white]** | - | - | - | - | - | - |
| **Assessment Centre [Newcastle]** | - | - | -0.18 (0.03, 2.13%) | 0.16 (0.02, 0.9%) | - | - |
| **BMI** | -0.10 (0.05, 3.57%) | 0.48 (0.40, 68.33%) | -0.11 (0.04, 3.32%) | 0.24 (0.15, 70.55%) | -0.08 (0.03, 0.98%) | 0.09 (0.11, 35.46%) |
| **WHR** | -0.28 (0.02, 0.37%) | 0.14 (0.01, 0.64%) | -0.13 (0.02, 0.8%) | 0.12 (0.05, 28.98%) | -0.06 (0.02, 0.29%) | 0.06 (0.05, 0.18%) |
| **Alcohol intake** | - | - | - | - | - | - |
| **Smoking status** | - | - | - | - | - | - |
| **NSAIDs for Pain** | - | - | - | - | - | - |
| **Dominant HGS** | - | - | - | - | - | 0.08 (0.04, 6.46%) |
| **Townsend deprivation index** | - | - | - | - | -0.08 (0.05, 2.69%) | - |
| **Fluid intake** | - | 0.30 (0.11, 2.75%) | - | 0.10 (0.03, 1.29%) | -0.05 (0.02, 0.07%) | 0.07 (0.04, 10.16%) |
| **Mid-Thigh IMAT** | - | - | -0.16 (0.01, 0.06%) | 0.17 (0.09, 4.08%) | - | 0.18 (0.16, 3.85%) |
| **CKD** | -1.15 (0.16, 0.41%) | - | -1.06 (0.07, 0.14%) | 0.68 (0.00, 0.02%) | -0.70 (0.58, 25.95%) | 0.25 (0.00, 0.04%) |
| **T2D** | -0.84 (0.15, 0.9%) | 1.69 (1.21, 16.35%) | - | 0.55 (0.26, 18.05%) | - | 0.64 (0.47, 5.19%) |
| **Hypertension** | -0.49 (0.10, 0.23%) | 0.32 (0.10, 1.6%) | - | - | - | - |
| **2nd Visit** | -0.11 (0.19, 0.27%) | 0.22 (0.05, 3.42%) | - | 0.18 (0.08, 5.5%) | -0.22 (0.22, 9.69%) | 0.13 (0.07, 6.72%) |
| **CKD * 2nd Visit** | - | - | - | - | - | - |
| **T2D * 2nd Visit** | - | - | -0.60 (0.12, 0.14%) | 0.71 (0.29, 0.74%) | - | - |
| **Hypertension * 2nd Visit** | - | - | - | - | - | - |
| **Men** | | | | | | |
| **Intercept** | - | 11.75 (11.23, 100%) | - | 8.32 (2.65, 100%) | - | 1.54 (1.78, 100%) |
| **Age at baseline** | -0.46 (0.39, 25.33%) | 0.11 (0.00, 0.04%) | -0.22 (0.08, 0.74%) | 0.16 (0.07, 25.23%) | -0.17 (0.19, 38.51%) | 0.07 (0.06, 8.28%) |
| **Age² at baseline** | - | - | - | - | -0.13 (0.09, 5.88%) | 0.04 (0.02, 0.15%) |
| **Race [Non-white]** | - | - | - | - | - | - |
| **Assessment Centre [Newcastle]** | - | 1.11 (0.20, 1.64%) | - | - | - | - |
| **BMI** | -0.67 (0.17, 4.94%) | 0.86 (0.96, 37.12%) | -0.18 (0.03, 0.37%) | 0.24 (0.12, 60.97%) | -0.16 (0.05, 0.94%) | 0.13 (0.14, 19.38%) |
| **WHR** | -0.37 (0.03, 0.1%) | 0.27 (0.10, 0.37%) | -0.18 (0.00, 0.23%) | 0.14 (0.04, 11.28%) | - | - |
| **Alcohol intake** | - | - | - | - | - | - |
| **Smoking status** | - | - | - | - | - | - |
| **NSAIDs for Pain** | - | - | - | 0.39 (0.00, 0.02%) | - | - |
| **Dominant HGS** | - | - | - | - | - | - |
| **Townsend deprivation index** | - | - | - | - | - | - |
| **Fluid intake** | - | - | - | - | - | - |
| **Mid-Thigh IMAT** | - | - | - | - | -0.03 (0.00, 0.04%) | 0.37 (0.17, 2.21%) |
| **CKD** | -1.52 (0.44, 2.69%) | - | - | - | -0.88 (0.52, 3.01%) | 0.31 (0.00, 0.04%) |
| **T2D** | -1.22 (0.21, 0.43%) | 2.58 (1.49, 8.1%) | - | 0.93 (0.66, 7.44%) | - | 0.43 (0.00, 0.04%) |
| **Hypertension** | - | - | - | - | - | 0.38 (0.04, 0.11%) |
| **2nd Visit** | -0.48 (0.00, 0.02%) | 0.27 (0.03, 1.27%) | -0.51 (0.10, 0.49%) | 0.26 (0.11, 6.28%) | -0.29 (0.11, 4.36%) | 0.15 (0.11, 0.33%) |
| **CKD * 2nd Visit** | - | - | - | - | - | - |
| **T2D * 2nd Visit** | - | - | -0.79 (0.05, 0.08%) | 1.08 (0.08, 0.14%) | - | - |
| **Hypertension * 2nd Visit** | - | - | - | - | -0.33 (0.00, 0.04%) | - |
| **Women** | | | | | | |
| **Intercept** | - | 9.74 (9.68, 100%) | - | 7.87 (2.56, 100%) | - | 1.38 (1.44, 100%) |
| **Age at baseline** | -0.57 (0.31, 32.59%) | 0.17 (0.03, 0.23%) | -0.14 (0.05, 3.71%) | 0.13 (0.06, 8.98%) | -0.13 (0.12, 36.04%) | 0.07 (0.03, 3.12%) |
| **Age² at baseline** | - | - | - | - | - | - |
| **Race [Non-white]** | - | - | - | - | - | - |
| **Assessment Centre [Newcastle]** | - | - | - | - | - | - |
| **BMI** | -0.13 (0.07, 3.69%) | 0.56 (0.38, 62.14%) | -0.14 (0.05, 3.12%) | 0.25 (0.17, 62.92%) | -0.05 (0.01, 0.15%) | 0.14 (0.13, 26.28%) |
| **WHR** | - | - | -0.14 (0.02, 1.27%) | 0.12 (0.05, 22.66%) | - | - |
| **Alcohol intake** | - | - | - | - | - | - |
| **Smoking status** | - | - | - | - | - | 0.26 (0.03, 0.07%) |
| **NSAIDs for Pain** | - | - | - | - | - | - |
| **Dominant HGS** | - | 0.33 (0.00, 0.02%) | - | - | -0.04 (0.01, 0.07%) | 0.09 (0.04, 2.36%) |
| **Townsend deprivation index** | - | - | - | - | - | - |
| **Fluid intake** | - | - | - | 0.11 (0.03, 2.58%) | - | 0.04 (0.00, 0.04%) |
| **Mid-Thigh IMAT** | - | - | - | 0.17 (0.03, 1.03%) | - | - |
| **CKD** | - | - | - | - | -1.01 (0.54, 7.19%) | 0.79 (0.00, 0.04%) |
| **T2D** | - | - | - | 0.92 (0.46, 1.78%) | - | 0.65 (0.00, 0.04%) |
| **Hypertension** | -0.51 (0.05, 0.14%) | 1.28 (0.79, 20.22%) | - | - | - | - |
| **2nd Visit** | -0.14 (0.01, 0.04%) | 0.24 (0.06, 1.09%) | -0.22 (0.03, 0.06%) | 0.21 (0.02, 0.53%) | -0.25 (0.23, 9.33%) | 0.17 (0.08, 2.83%) |
| **CKD * 2nd Visit** | - | - | - | - | - | - |
| **T2D * 2nd Visit** | - | - | -0.76 (0.19, 0.25%) | 0.89 (0.18, 6.75%) | - | - |
| **Hypertension * 2nd Visit** | - | - | - | - | - | - |

**Table S11.** Significance areas for covariates in the mass univariate linear mixed-effect models for the anthropometric variables for all (N=2,610), men (N=1,305) and women (N=1,305) of the model for the right kidney substructures (parenchyma, cortex and medulla). The total area has been split into areas of positive, negative associations. The standardised regression coefficients ($\hat{\beta}$) are presented as median (interquartile range - IQR) across all vertices of the left kidney substructure’s surface and the significance areas as a percentage (%) of the vertices with statistically significant associations.

Supplementary Figures

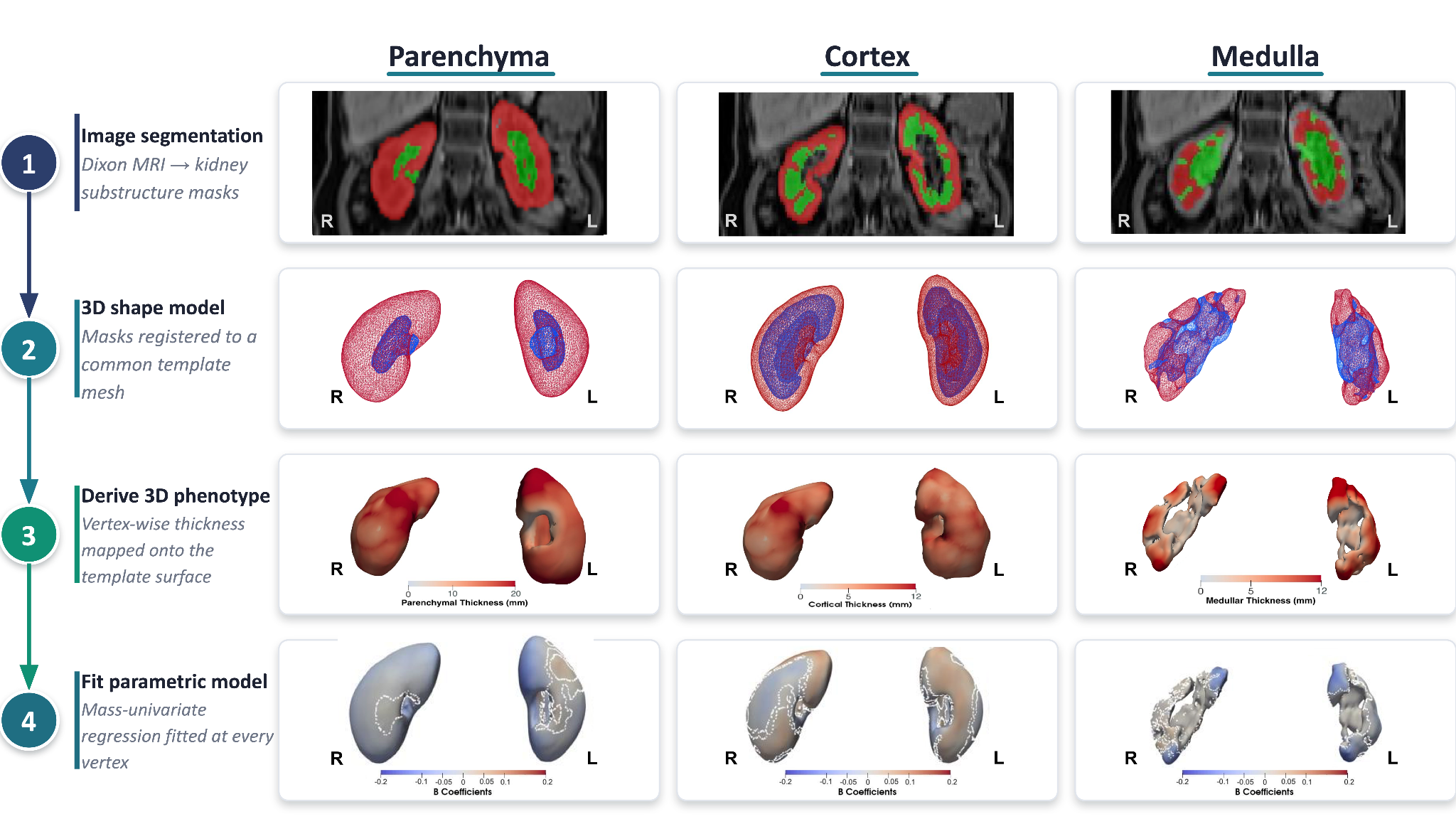

**Figure S1.** Overview of the 3D mesh construction, registration, and analysis workflow. Neck-to-knee Dixon MRI images from the UK Biobank are segmented to isolate the kidney substructures (cortex, medulla, parenchyma, vessels); a population template mesh is built and each participant's segmentation is registered to this common template using rigid, affine, and non-rigid registration; per-vertex thickness is computed for each substructure from the registered mesh; and the resulting thickness values are analysed cross-sectionally and longitudinally using mass univariate regression, with results visualised on the template mesh. Reproduced by kind permission of UK Biobank ©.

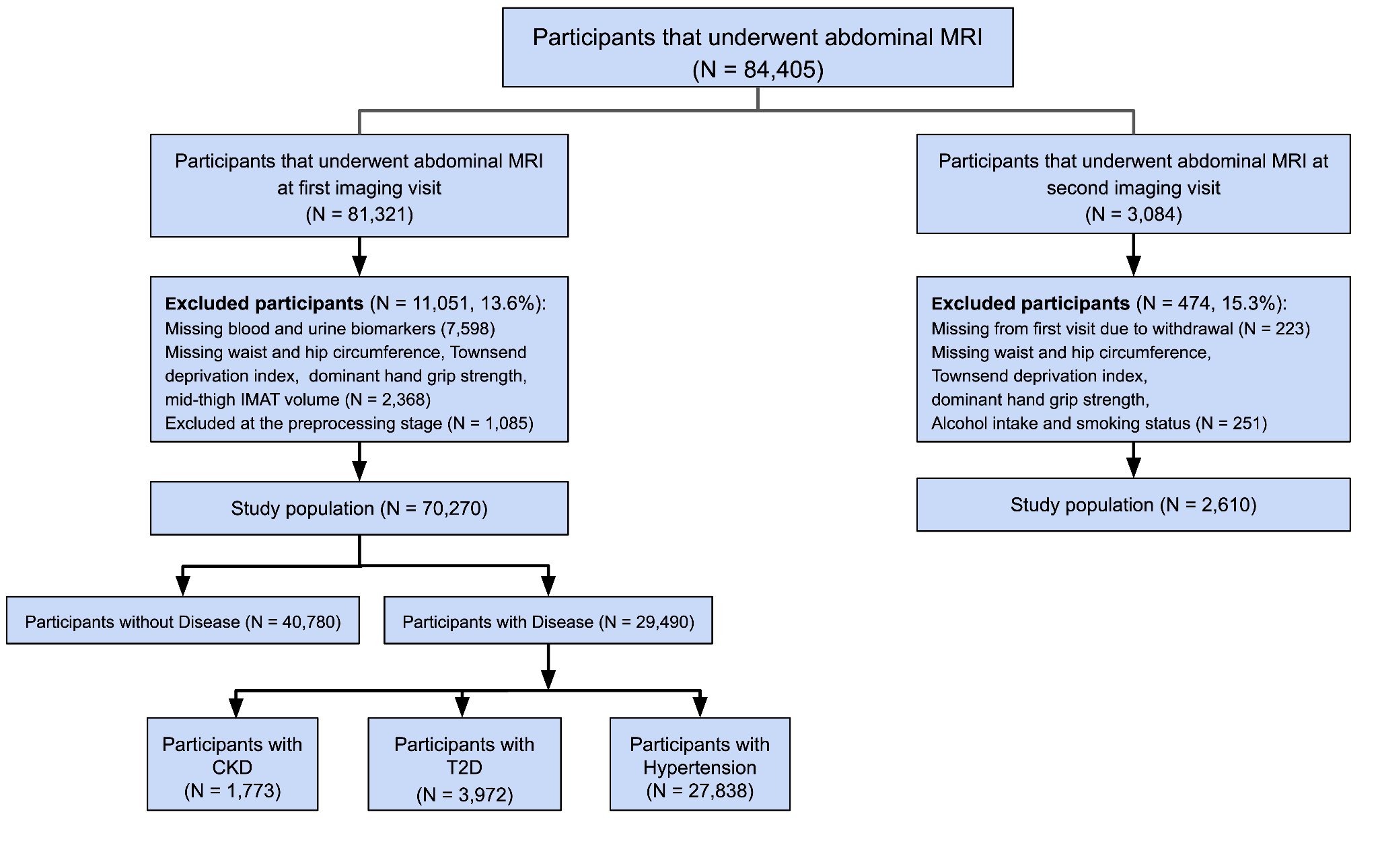
**Figure S2.** Flow diagram of participant selection. Of 84,405 UK Biobank participants who underwent abdominal MRI, 81,321 attended the first (baseline) imaging visit and 3,084 attended a second (follow-up) imaging visit. At baseline, 11,051 participants (13.6%) were excluded for missing blood or urine biomarkers, missing anthropometric or lifestyle covariates, or kidney segmentations that did not pass quality control, leaving a baseline study population of 70,270 participants (40,780 without CKD, T2D, or hypertension and 29,490 with at least one of these conditions: 1,773 with CKD, 3,972 with T2D, and 27,838 with hypertension). At follow-up, 474 participants (15.3%) were excluded for withdrawal or missing covariates, leaving a longitudinal study population of 2,610 participants.

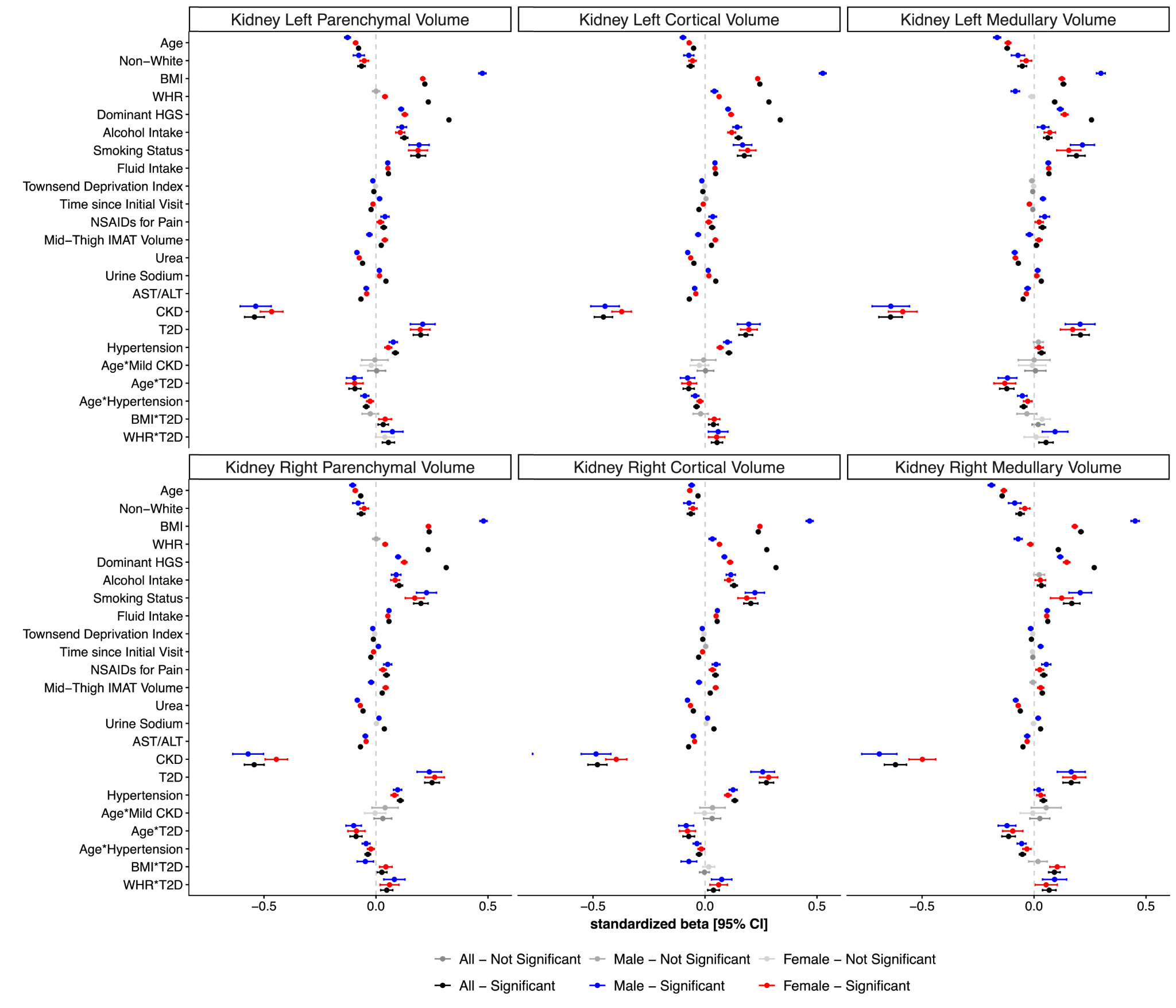

**Figure S3.** Standardised regression coefficients (β, 95% CI) from mass univariate regression models of kidney substructure volume (parenchyma, cortex, medulla; left and right) on anthropometric and clinical covariates (age, ethnicity, BMI, waist-to-hip ratio [WHR], dominant hand grip strength, alcohol intake, smoking status, fluid intake, Townsend deprivation index, time since initial assessment, NSAID use, mid-thigh intermuscular adipose tissue [IMAT] volume, urea, urine sodium, AST:ALT ratio, CKD, T2D, hypertension, and age- or adiposity-by-disease interaction terms), fitted in the full cohort and separately in men and women. Black points denote significant associations in the full cohort, blue in men, and red in women; grey points denote non-significant associations. Error bars show 95% confidence intervals.

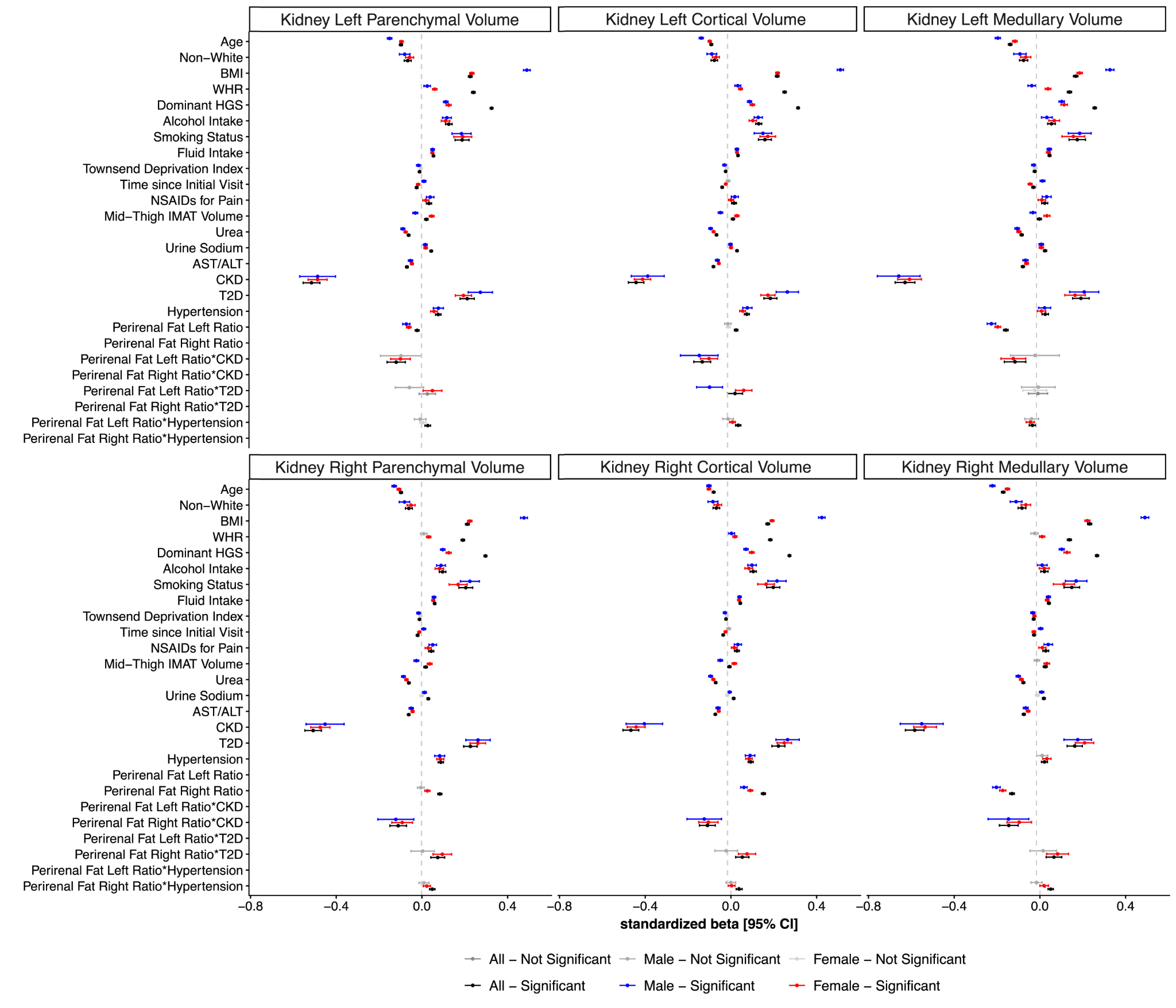

**Figure S4.** Standardised regression coefficients (β, 95% CI) from mass univariate regression models of kidney substructure volume (parenchyma, cortex, medulla; left and right) on the covariates in Supplementary Figure S3 plus left and right perirenal fat ratio and its interactions with CKD, T2D, and hypertension, fitted in the full cohort and separately in men and women. Black points denote significant associations in the full cohort, blue in men, and red in women; grey points denote non-significant associations. Error bars show 95% confidence intervals.

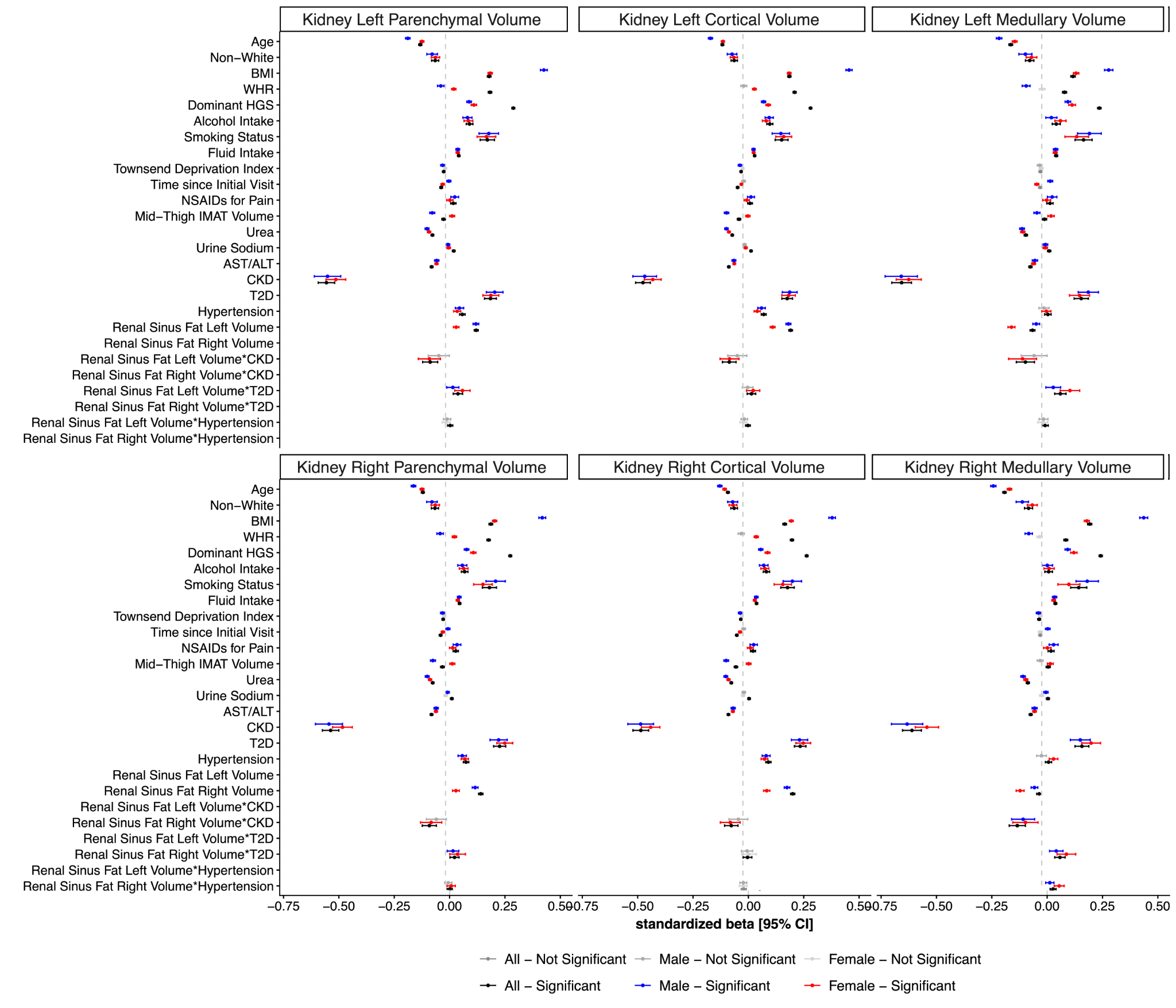

**Figure S5.** Standardised regression coefficients (β, 95% CI) from mass univariate regression models of kidney substructure volume (parenchyma, cortex, medulla; left and right) on the covariates in Supplementary Figure S3 plus left and right renal sinus fat volume and its interactions with CKD, T2D, and hypertension, fitted in the full cohort and separately in men and women. Black points denote significant associations in the full cohort, blue in men, and red in women; grey points denote non-significant associations. Error bars show 95% confidence intervals.

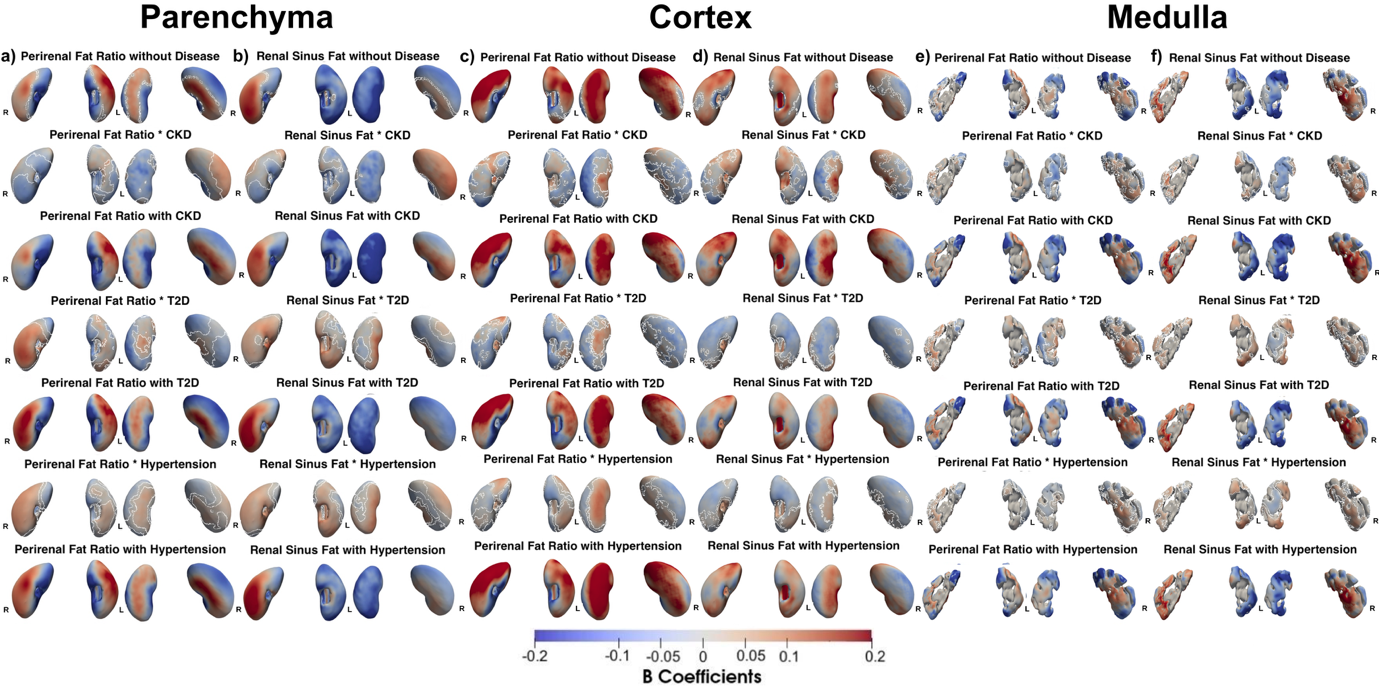
**Figure S6.** Vertex-wise associations between kidney-adjacent fat depots and disease status on kidney substructure thickness. Three-dimensional statistical parametric maps (SPMs) show, for the kidney parenchyma (a, b), cortex (c, d), and medulla (e, f), the local association with thickness for the left (L) and right (R) kidney (anterior views, left plots; posterior views, right plots), in the full cohort (N=70,270). Panels (a, c, e) show associations with perirenal fat ratio, and panels (b, d, f) with renal sinus fat. Within each panel, the top row shows the association in participants without chronic kidney disease (CKD), type-2 diabetes (T2D), or hypertension. The following rows show, for each disease in turn, the interaction effect alone, from the perirenal fat ratio×disease (or renal sinus fat×disease) regression coefficient, and the total association in participants with that disease, from the sum of the main and interaction coefficients for CKD, T2D, hypertension. Red indicates a positive (outward/thicker) association and blue a negative (inward/thinner) association. Regression coefficients ($\hat{\beta}$) are shown with units in standard deviations for each covariate.

**Men**

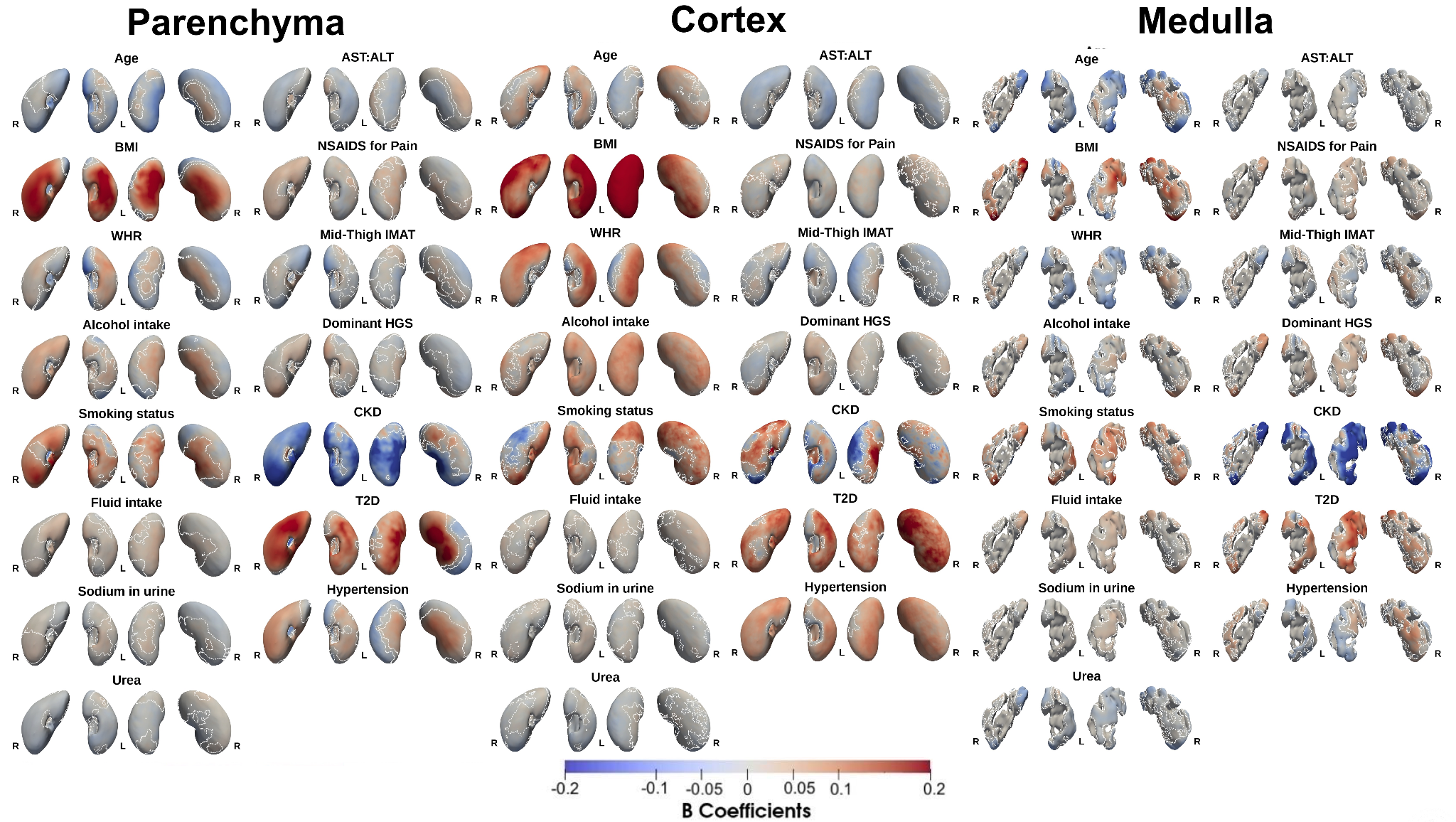

**Figure S7.** Three-dimensional statistical parametric maps (SPMs) show, for each covariate (age, BMI, waist-to-hip ratio (WHR), alcohol intake, smoking status, fluid intake, sodium in urine, urea, AST:ALT ratio, NSAID use, mid-thigh IMAT, dominant hand grip strength, chronic kidney disease (CKD), type-2 diabetes (T2D), and hypertension), the local strength of association with thickness at each vertex of the kidney parenchyma, cortex, and medulla surface meshes, from mass univariate regression models fitted in the men cohort (N=34,044). For each covariate, the anterior and posterior surfaces of the left (L) and right (R) kidney are shown (anterior views, left plots; posterior views, right plots). White contour lines mark the boundary of statistically significant regions (p < 0.05 after correction for multiplicity). Red indicates a positive (outward/thicker) association and blue a negative (inward/thinner) association. Colour intensity reflects the standardised regression coefficient ($\hat{\beta}$, in units of SD).

**Women**

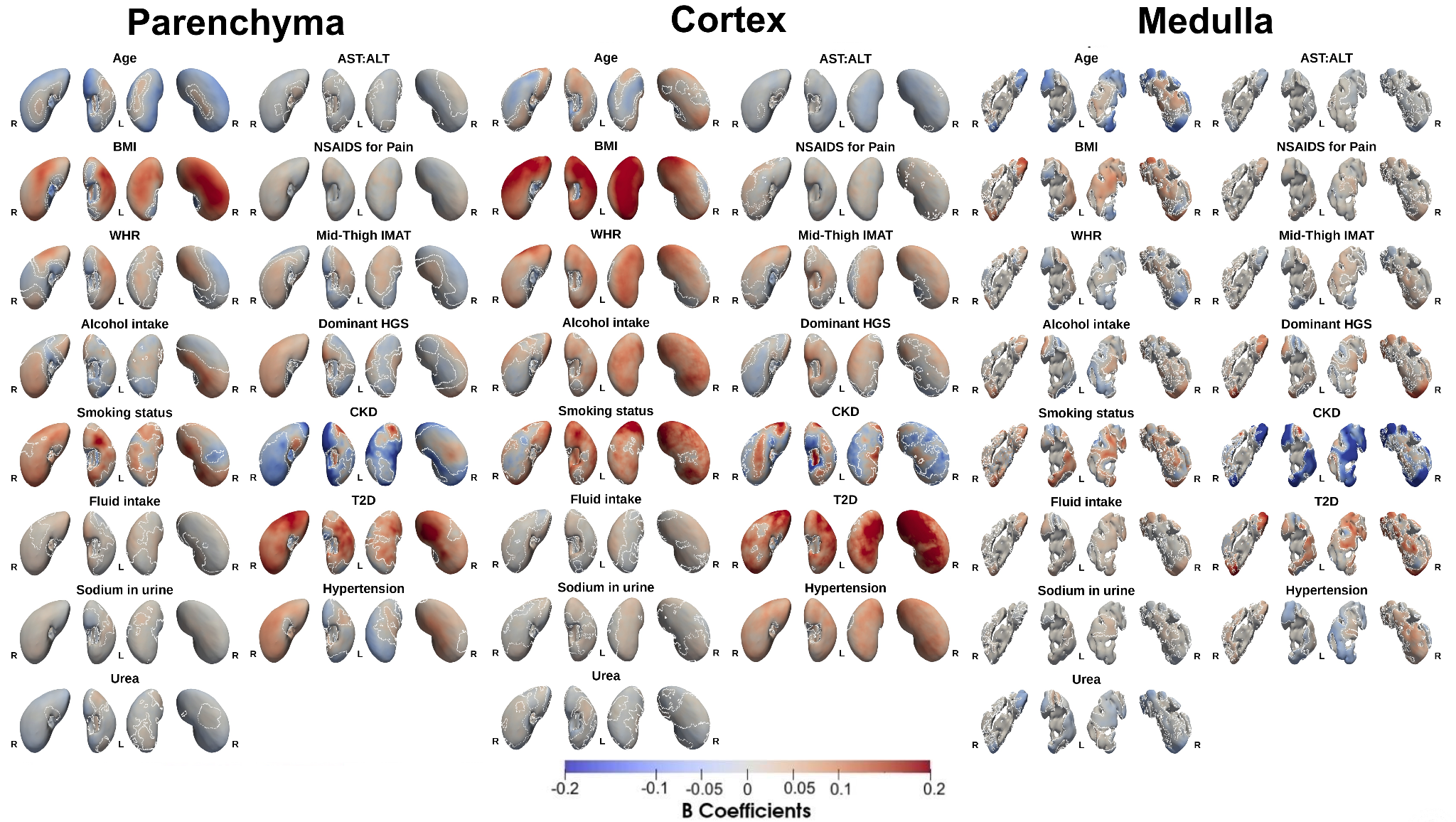

**Figure S8.** Three-dimensional statistical parametric maps (SPMs) show, for each covariate (age, BMI, waist-to-hip ratio (WHR), alcohol intake, smoking status, fluid intake, sodium in urine, urea, AST:ALT ratio, NSAID use, mid-thigh IMAT, dominant hand grip strength, chronic kidney disease (CKD), type-2 diabetes (T2D), and hypertension), the local strength of association with thickness at each vertex of the kidney parenchyma, cortex, and medulla surface meshes, from mass univariate regression models fitted in the women cohort (N=36,226). For each covariate, the anterior and posterior surfaces of the left (L) and right (R) kidney are shown (anterior views, left plots; posterior views, right plots). White contour lines mark the boundary of statistically significant regions (p < 0.05 after correction for multiplicity). Red indicates a positive (outward/thicker) association and blue a negative (inward/thinner) association. Colour intensity reflects the standardised regression coefficient ($\hat{\beta}$, in units of SD).

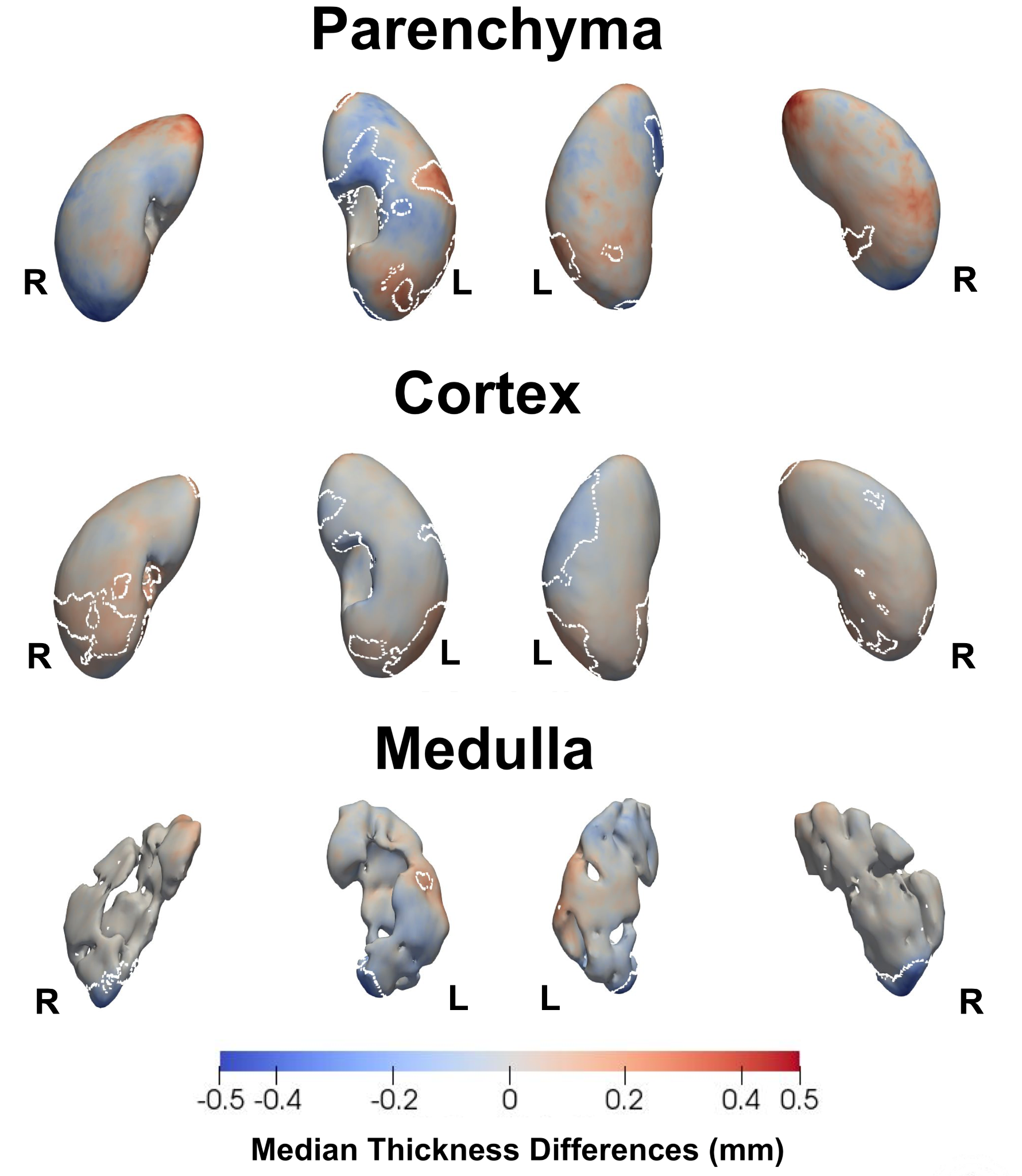

**Figure S9.** Median thickness differences (mm) between the baseline and follow-up imaging visits, mapped onto the kidney parenchyma, cortex, and medulla surface meshes for the left (L) and right (R) kidney, in the longitudinal cohort (N = 2,610). White contour lines mark the boundary of vertices with a statistically significant difference between visits (FDR-corrected p < 0.05); red indicates an outward (thicker) change at follow-up and blue an inward (thinner) change.

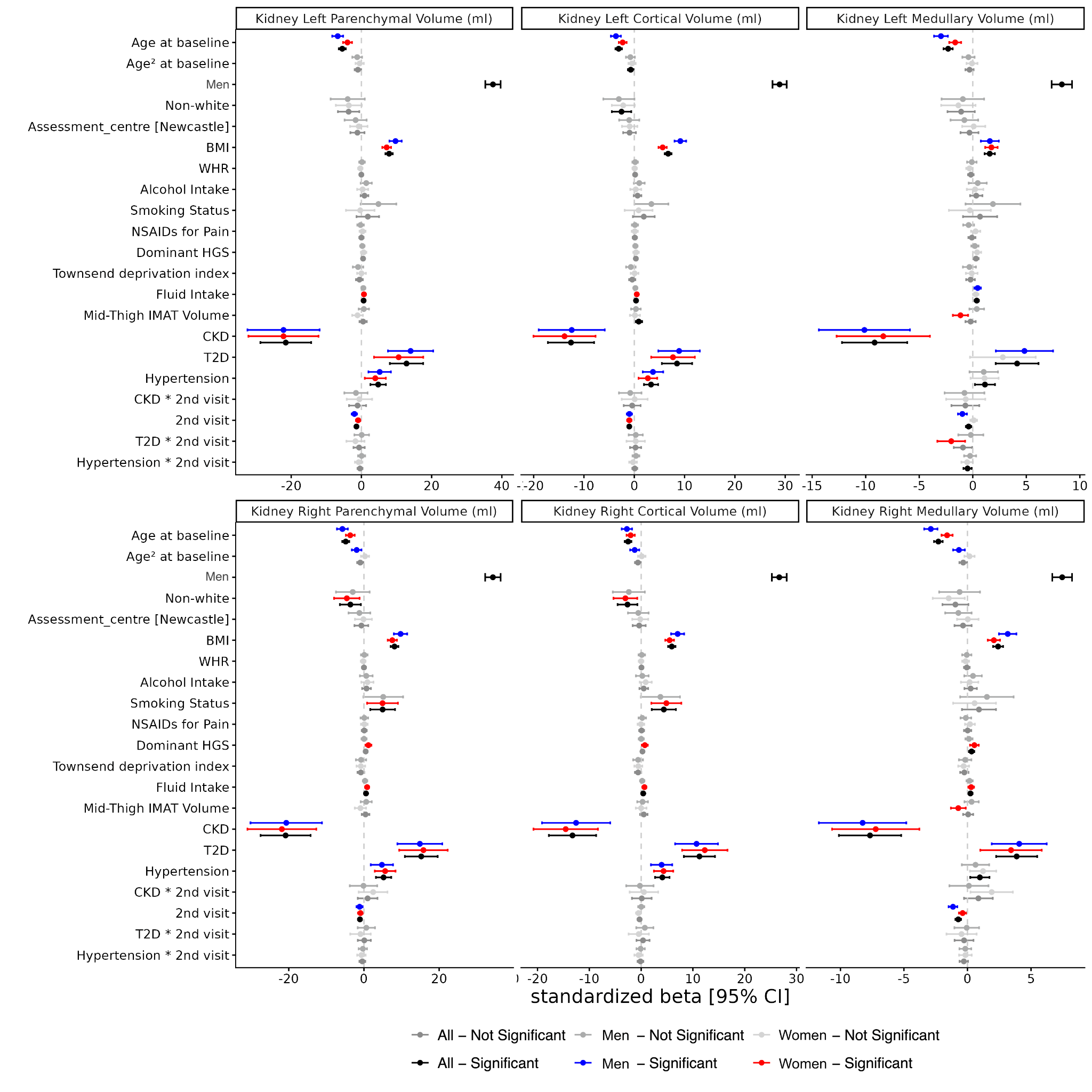

**Figure S10.** Summary of standardised regression coefficients from the longitudinal linear mixed-effects models of kidney substructure volume (image-derived phenotypes, IDPs; parenchyma, cortex, medulla, left and right), each measured in millilitres (mL), on imaging visit and the anthropometric and clinical covariates listed in Supplementary Tables S9-S10, fitted in the longitudinal cohort (N = 2,610).

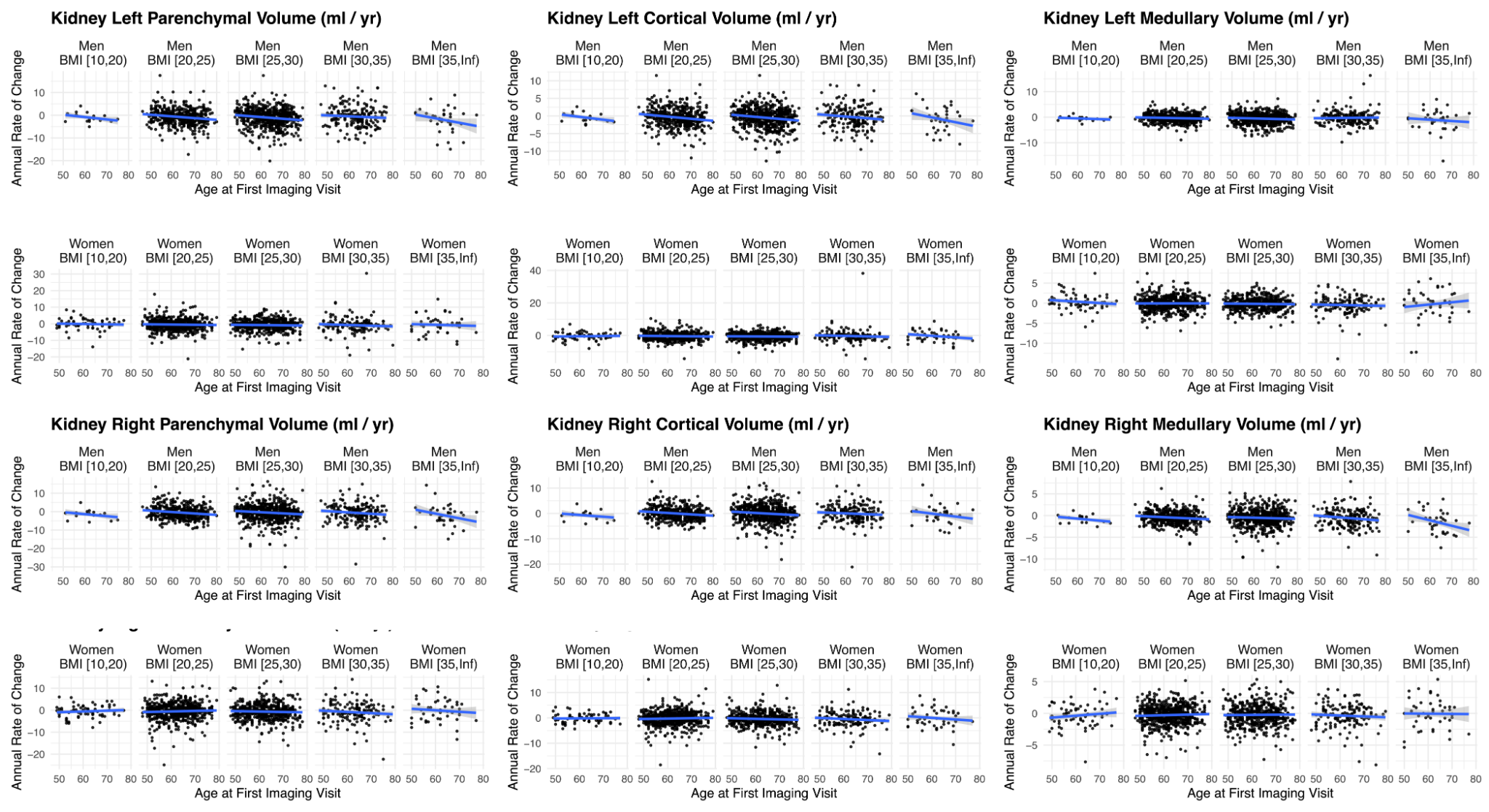

**Figure S11.** Annual rate of change in kidney substructure volume (mL/year) with age at the first imaging visit, stratified by sex and by baseline BMI category (10-20, 20-25, 25-30, 30-35, and >35 kg/m²), for kidney parenchyma, cortex, and medulla volume, left and right, in the longitudinal cohort (N = 2,610). Each point is one participant's annual rate of change between the two imaging visits; blue lines show the linear trend with age within each BMI-by-sex stratum, with shaded 95% confidence bands.

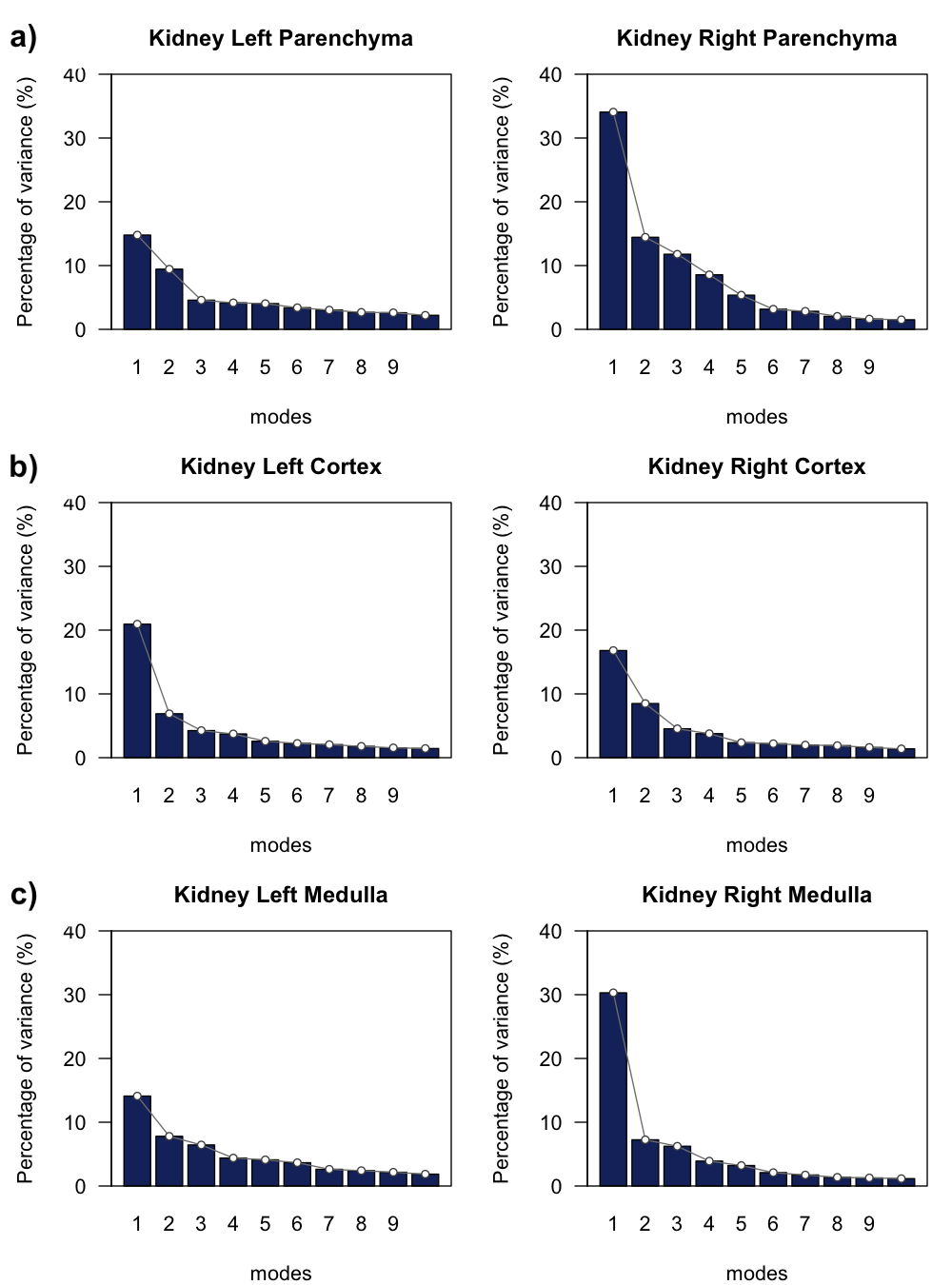
**Figure S12.** Percentage of shape variance explained by the first ten principal component (PC) modes from statistical shape analysis of the left and right kidney a) parenchyma, b) cortex, and c) medulla, in the full cohort (N = 70,270).

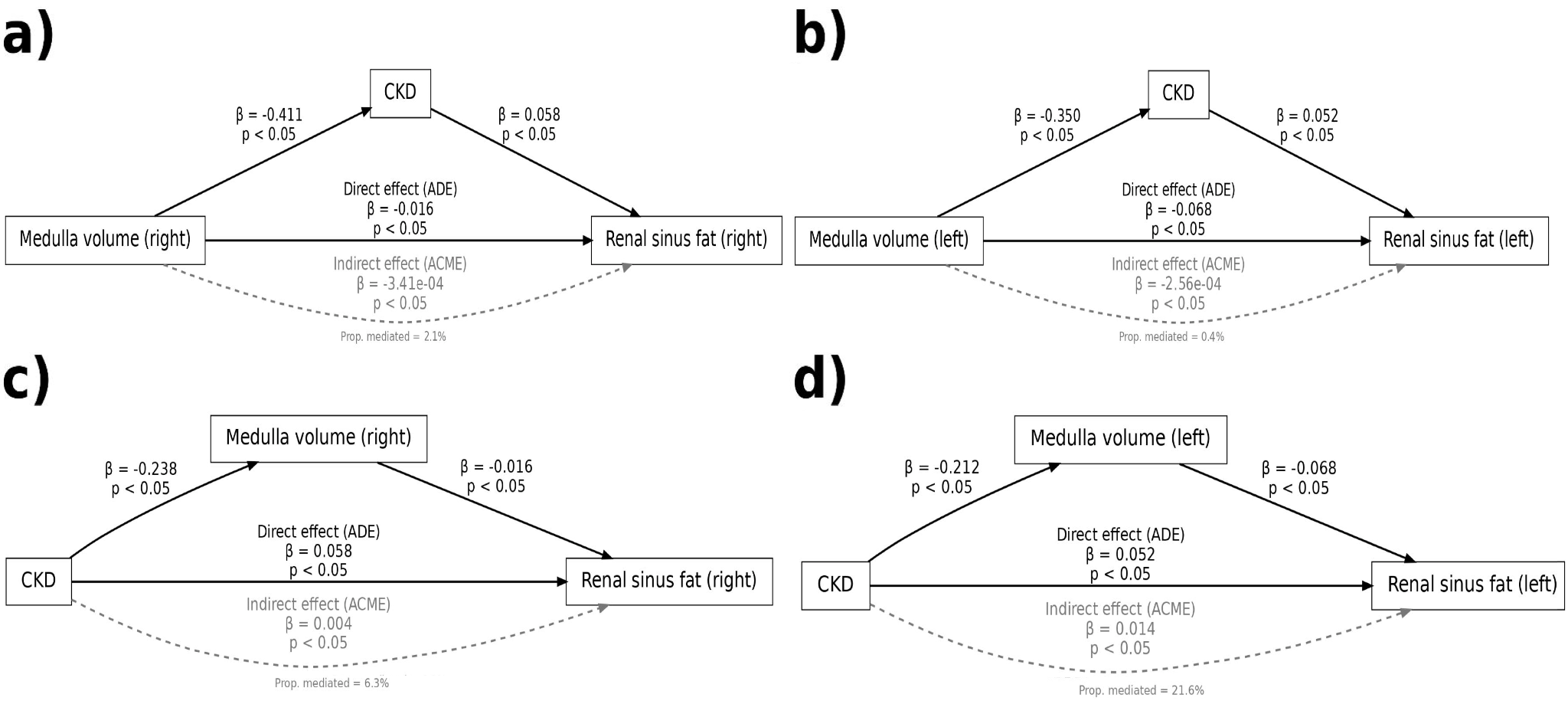
**Figure S13.** Causal mediation analysis on the relationship between kidney medulla volume, CKD and renal sinus fat in the overall cohort (N=70,270). Upper panels **a)** and **b)** show exposure kidney medulla left and right volume with mediator CKD. Lower panels **c)** and **d)** show exposure CKD with mediator kidney medulla left and right volume. Direct effects are indicated by solid black arrows, whereas mediation effects are indicated by dashed grey arrows. Labels indicate effect size and p-values.

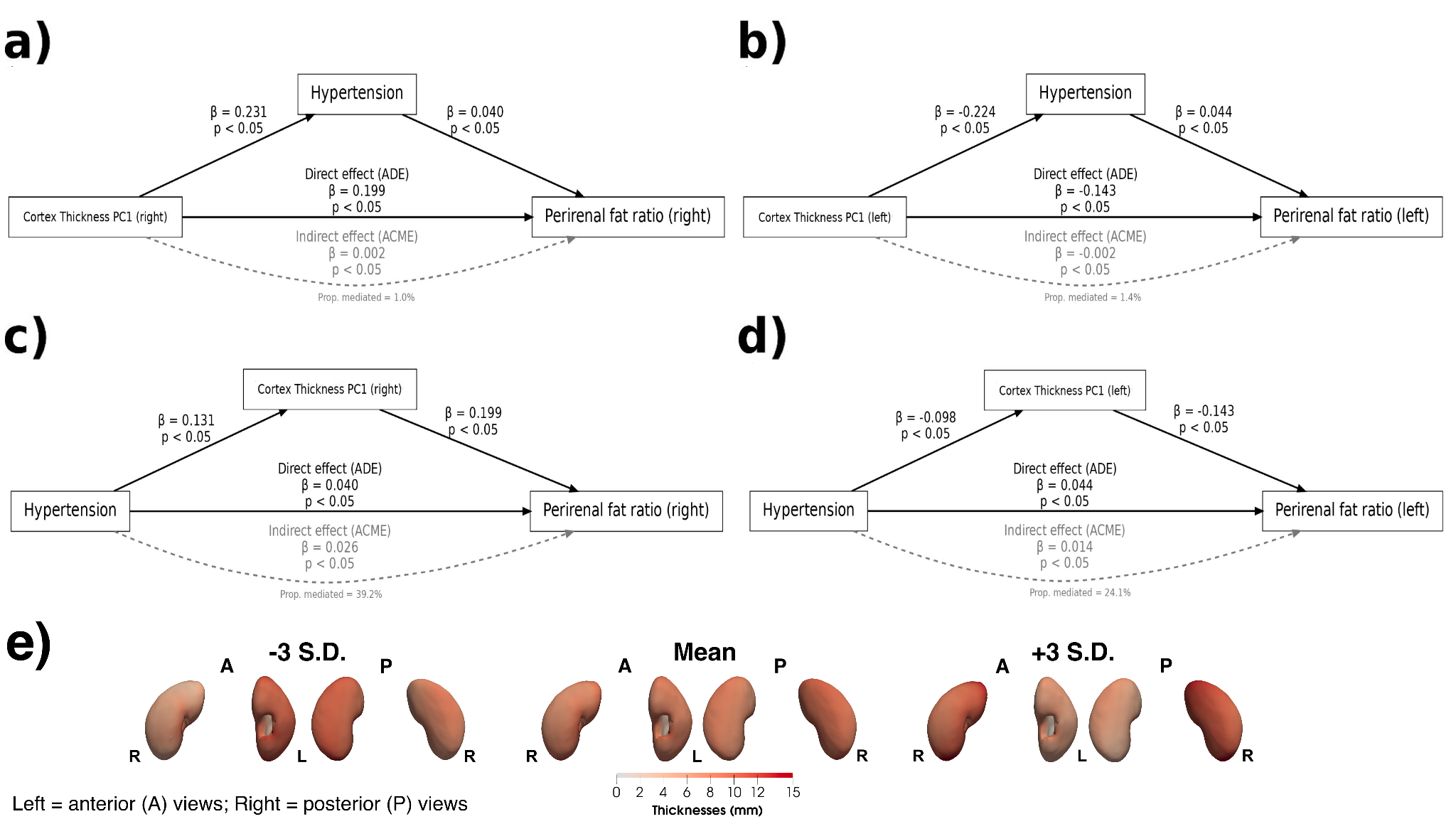

**Figure S14.** Causal mediation analysis on the relationship between kidney cortex thickness PC1, reflecting global cortical thickness variations, hypertension and perirenal fat ratio in the overall cohort (N=70,270). Upper panels **a)** and **b)** show exposure kidney cortex left and right thickness PC1 with mediator hypertension. Lower panels **c)** and **d)** show exposure hypertension with mediator kidney cortex left and right thickness PC1. **e)** The mean cortex thickness and the thickness at the ±3 SD are displayed for PC1, showing the thickness variation in mm. The thickness variations are shown in the anterior (A) and posterior (P) views of each kidney. Direct effects are indicated by solid black arrows, whereas mediation effects are indicated by dashed grey arrows. Labels indicate effect size and p-values.

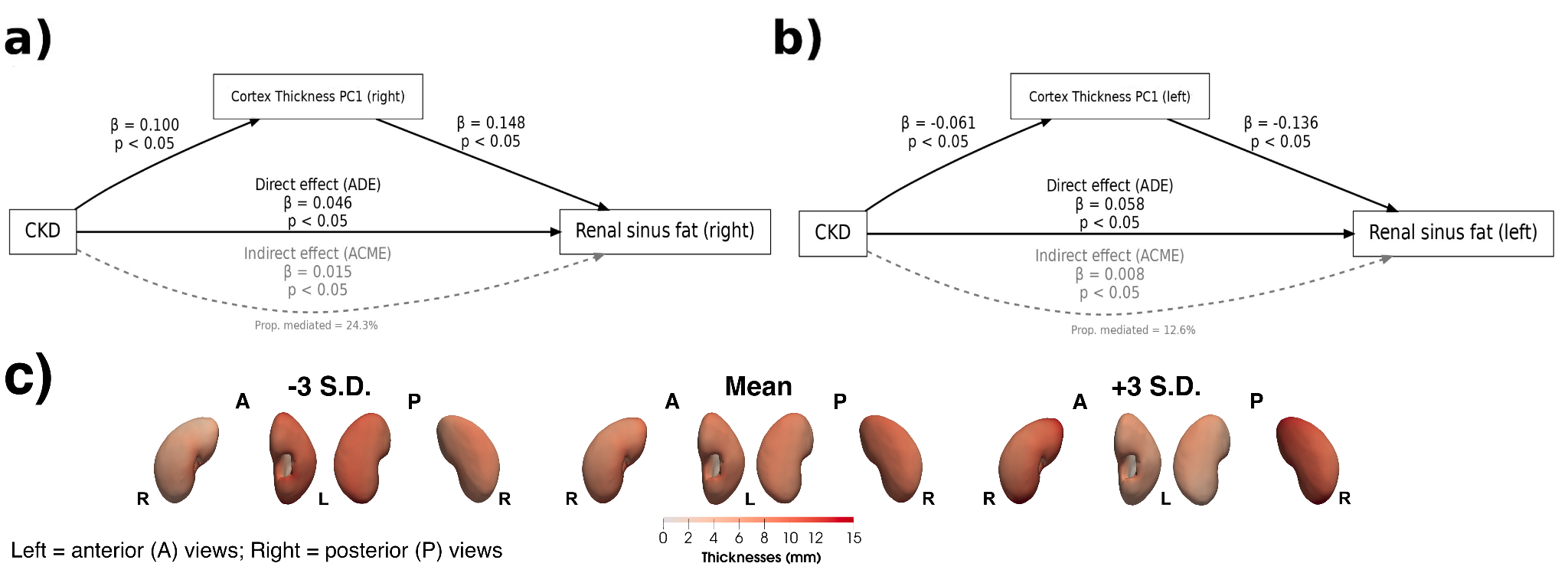
**Figure S15.** Causal mediation analysis on the relationship between kidney cortex thickness PC1, reflecting global cortical thickness variations, CKD and renal sinus fat in the overall cohort (N=70,270). **a)** and **b)** show exposure CKD with mediator kidney cortex left and right thickness PC1. **c)** The mean cortex thickness and the thickness at the ±3 SD are displayed for PC1, showing the thickness variation in mm. The thickness variations are shown in the anterior (A) and posterior (P) views of each kidney.Direct effects are indicated by solid black arrows, whereas mediation effects are indicated by dashed grey arrows. Labels indicate effect size and p-values.

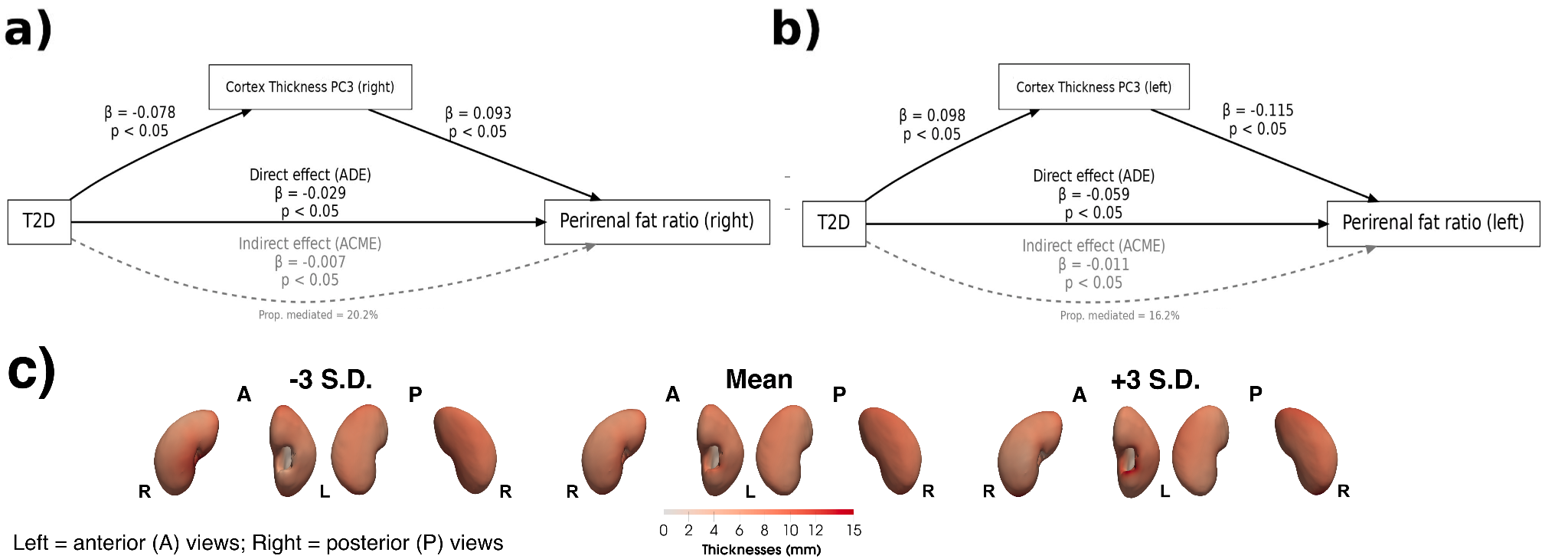

**Figure S16.** Causal mediation analysis on the relationship between kidney cortex thickness PC3, reflecting the variation at the inferior pole and medial border near the hilum, T2D and perirenal fat ratio in the overall cohort (N=70,270). **a)** and **b)** show exposure T2D with mediator kidney cortex left and right thickness PC3. **c)** The mean cortex thickness and the thickness at the ±3 SD are displayed for PC3, showing the thickness variation in mm. The thickness variations are shown in the anterior (A) and posterior (P) views of each kidney. Direct effects are indicated by solid black arrows, whereas mediation effects are indicated by dashed grey arrows. Labels indicate effect size and p-values.

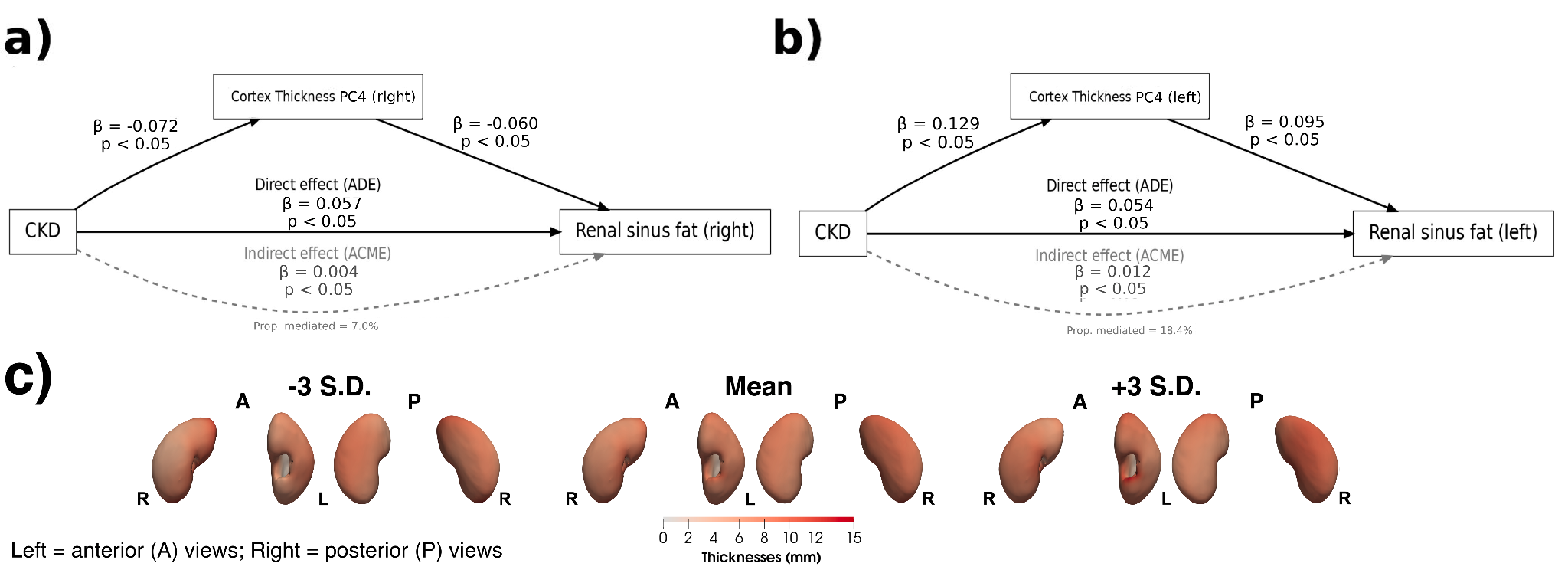

**Figure S17.** Causal mediation analysis on the relationship between kidney cortex thickness PC4, reflecting localised foci along the medial and hilar variations, CKD and renal sinus fat in the overall cohort (N=70,270). **a)** and **b)** show CKD exposure with mediator kidney cortex left and right thickness PC4. **c)** The mean cortex thickness and the thickness at the ±3 SD are displayed for PC4, showing the thickness variation in mm. The thickness variations are shown in the anterior (A) and posterior (P) views of each kidney.Direct effects are indicated by solid black arrows, whereas mediation effects are indicated by dashed grey arrows. Labels indicate effect size and p-values. Abbreviations: CKD: chronic kidney disease.

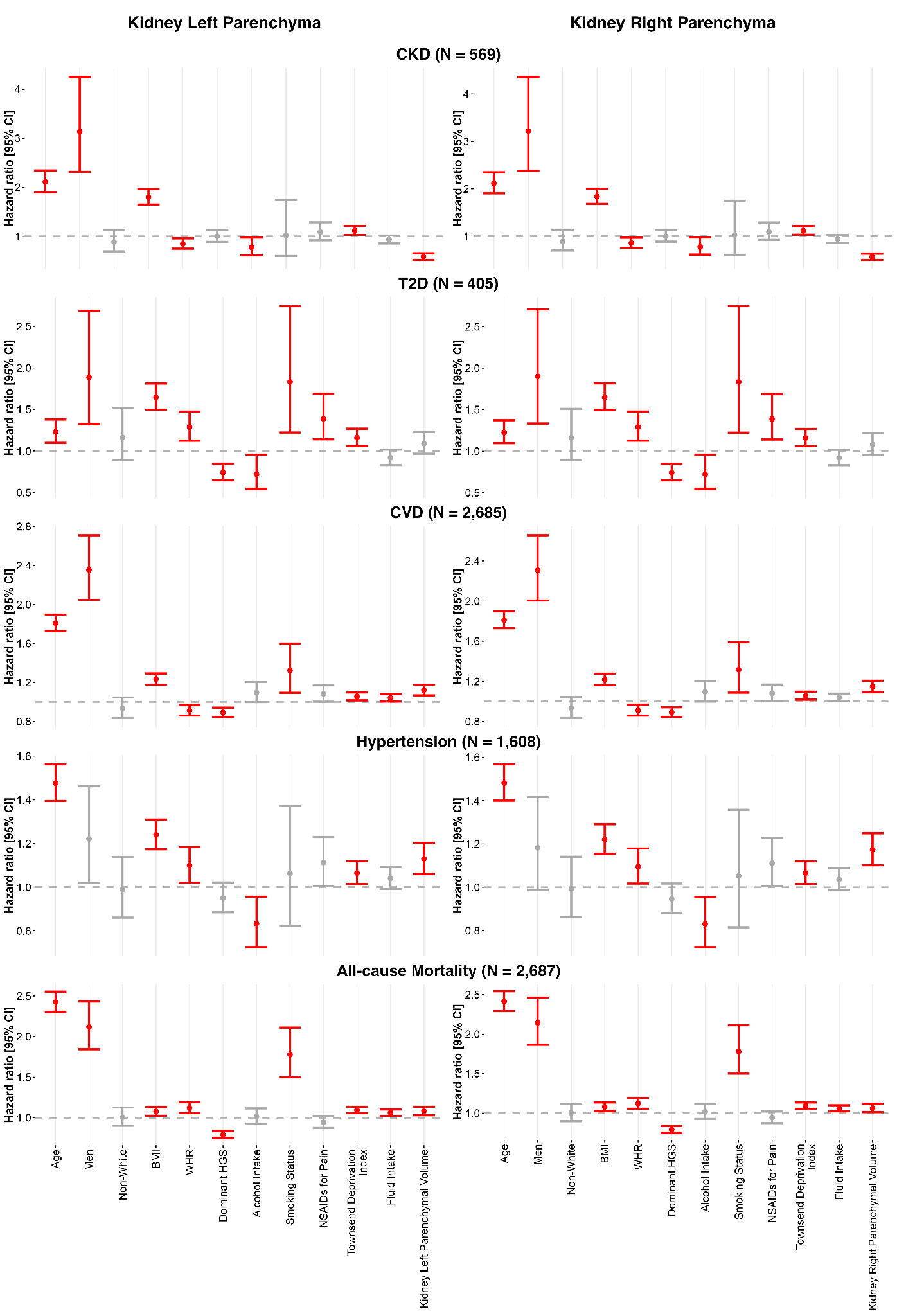

**Figure S18.** Hazard ratios (HR) and 95% confidence intervals (CI) for incident CKD, T2D, CVD, hypertension, and all-cause mortality associated with left and right kidney parenchymal volume, from Cox proportional hazards models adjusted for age, sex, ethnicity, BMI, WHR, dominant hand grip strength, alcohol intake, smoking status, NSAID use, Townsend deprivation index, and fluid intake. Significant associations (p < 0.05) are shown in red and non-significant associations in grey.

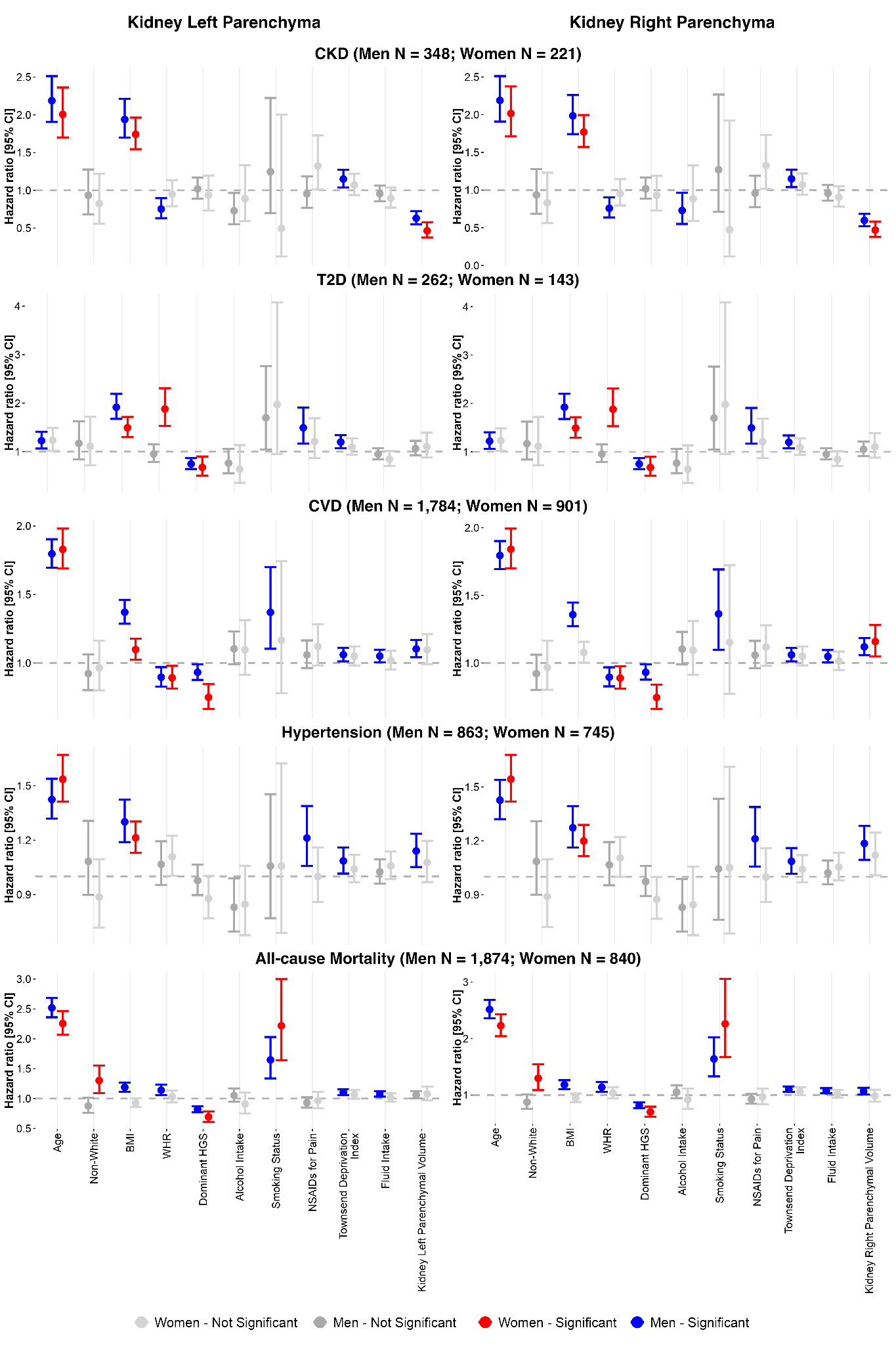

**Figure S19.** Sex-stratified hazard ratios (HR) and 95% confidence intervals (CI) for incident CKD, T2D, CVD, hypertension, and all-cause mortality associated with left and right kidney parenchymal volume, from Cox proportional hazards models adjusted for age, ethnicity, BMI, WHR, dominant hand grip strength, alcohol intake, smoking status, NSAID use, Townsend deprivation index, and fluid intake, fitted separately in men and women. Significant associations (p < 0.05) are shown in blue (men) and red (women); non-significant associations are shown in grey.

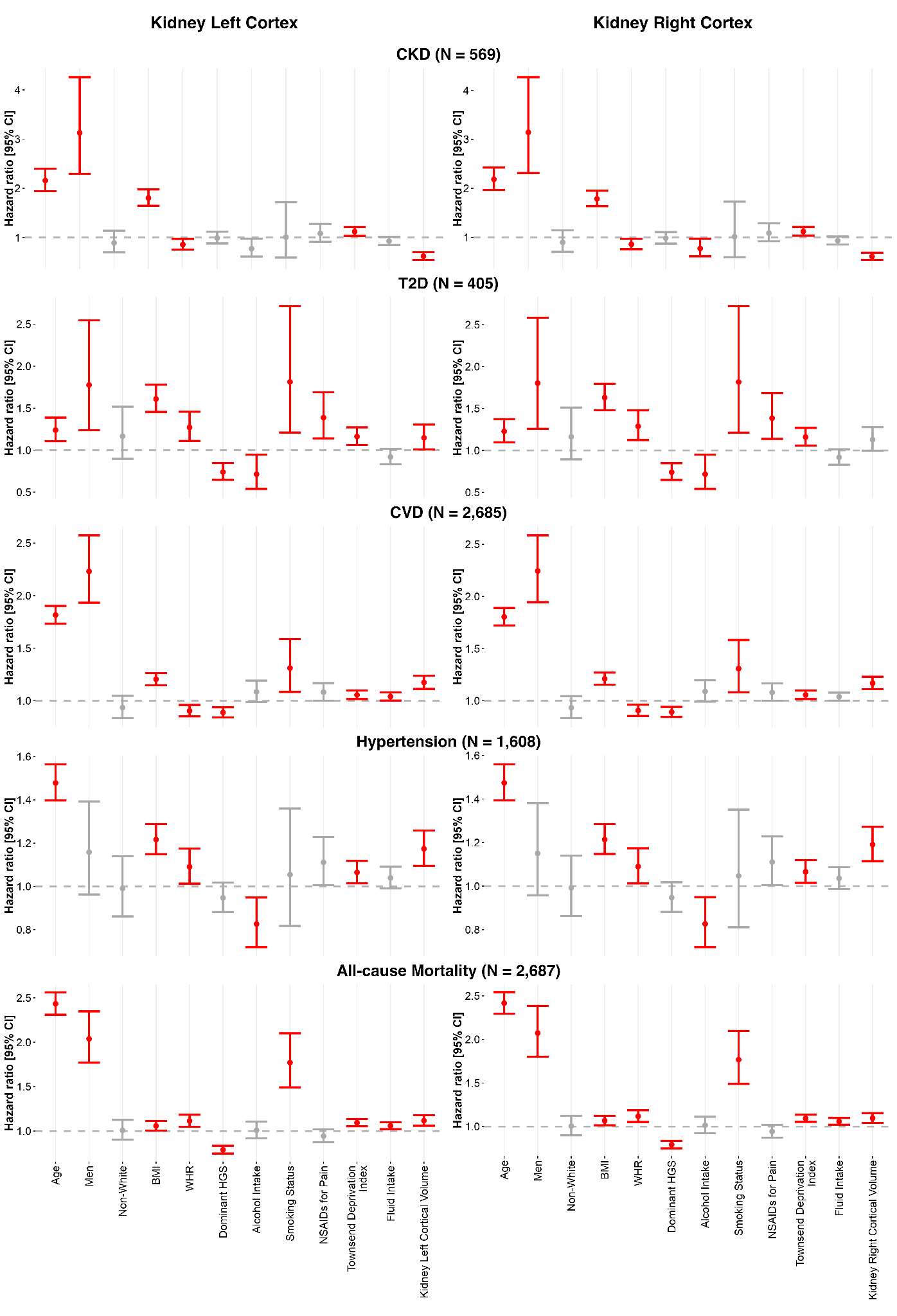

**Figure S20.** Hazard ratios (HR) and 95% confidence intervals (CI) for incident CKD, T2D, CVD, hypertension, and all-cause mortality associated with left and right kidney cortex volume, from Cox proportional hazards models adjusted for age, sex, ethnicity, BMI, WHR, dominant hand grip strength, alcohol intake, smoking status, NSAID use, Townsend deprivation index, and fluid intake. Significant associations (p < 0.05) are shown in red and non-significant associations in grey.

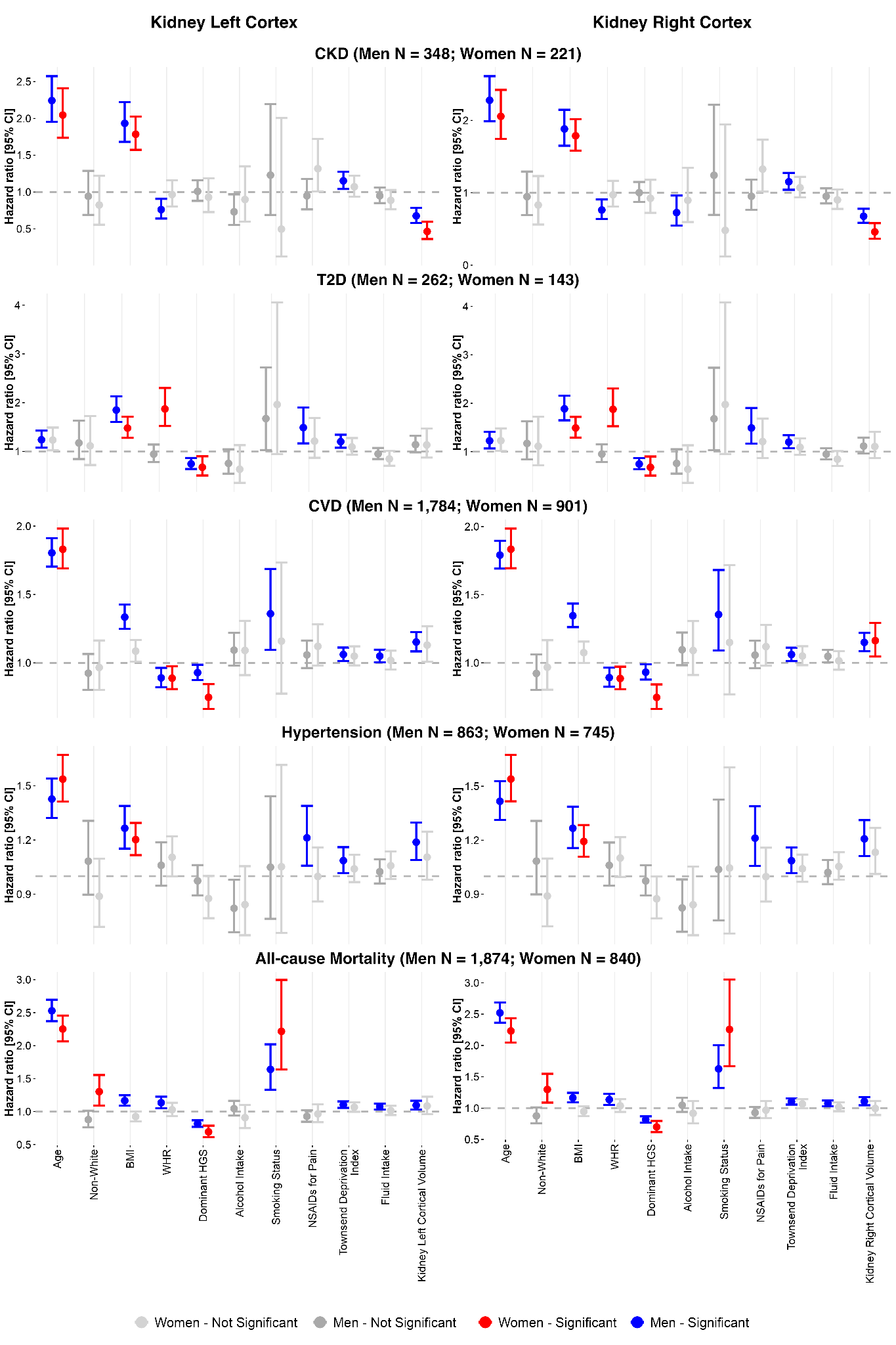

**Figure S21.** Sex-stratified hazard ratios (HR) and 95% confidence intervals (CI) for incident CKD, T2D, CVD, hypertension, and all-cause mortality associated with left and right kidney cortex volume, from Cox proportional hazards models adjusted for age, ethnicity, BMI, WHR, dominant hand grip strength, alcohol intake, smoking status, NSAID use, Townsend deprivation index, and fluid intake, fitted separately in men and women. Significant associations (p < 0.05) are shown in blue (men) and red (women); non-significant associations are shown in grey.

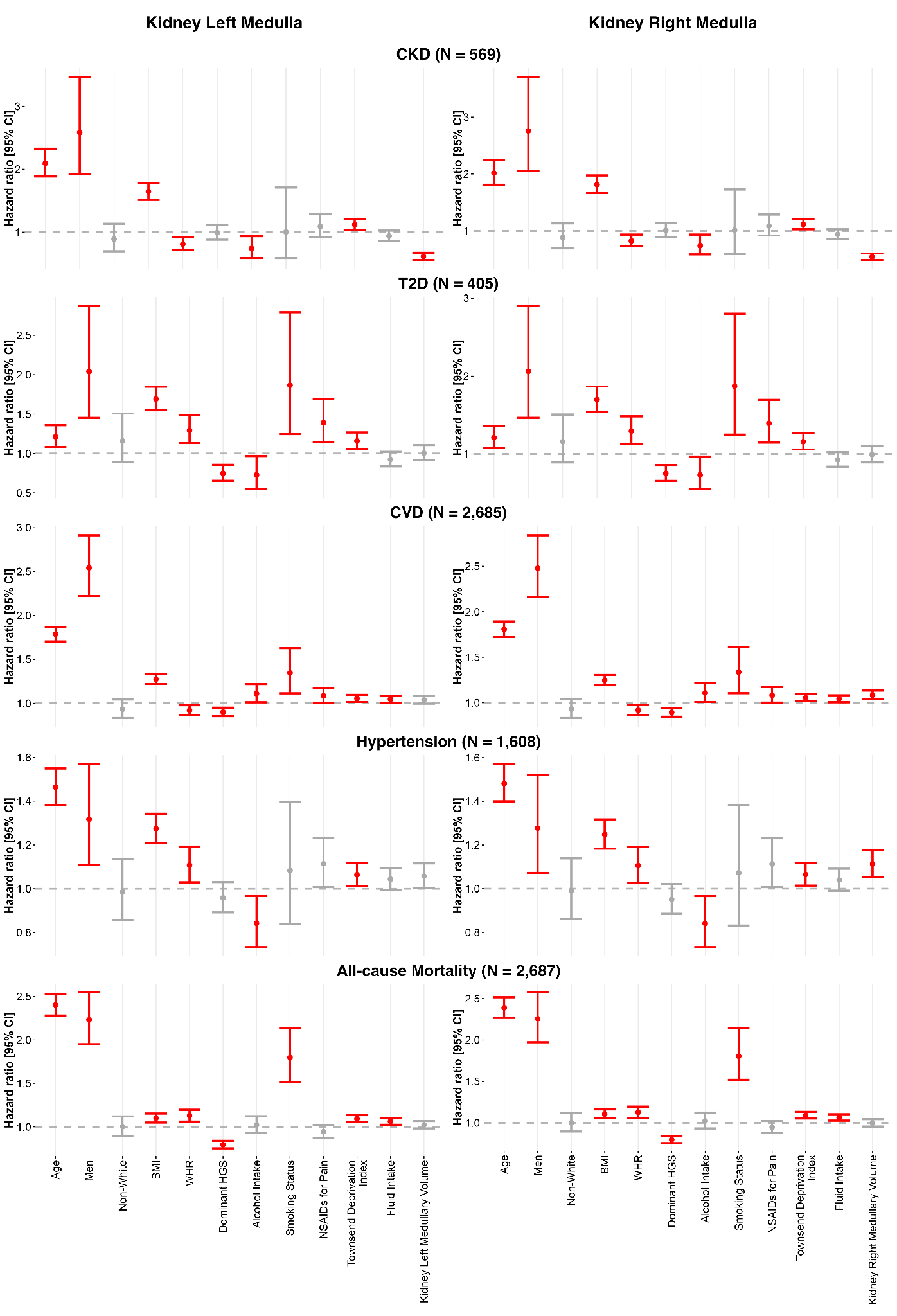

**Figure S22.** Hazard ratios (HR) and 95% confidence intervals (CI) for incident CKD, T2D, CVD, hypertension, and all-cause mortality associated with left and right kidney medulla volume, from Cox proportional hazards models adjusted for age, sex, ethnicity, BMI, WHR, dominant hand grip strength, alcohol intake, smoking status, NSAID use, Townsend deprivation index, and fluid intake. Significant associations (p < 0.05) are shown in red and non-significant associations in grey.

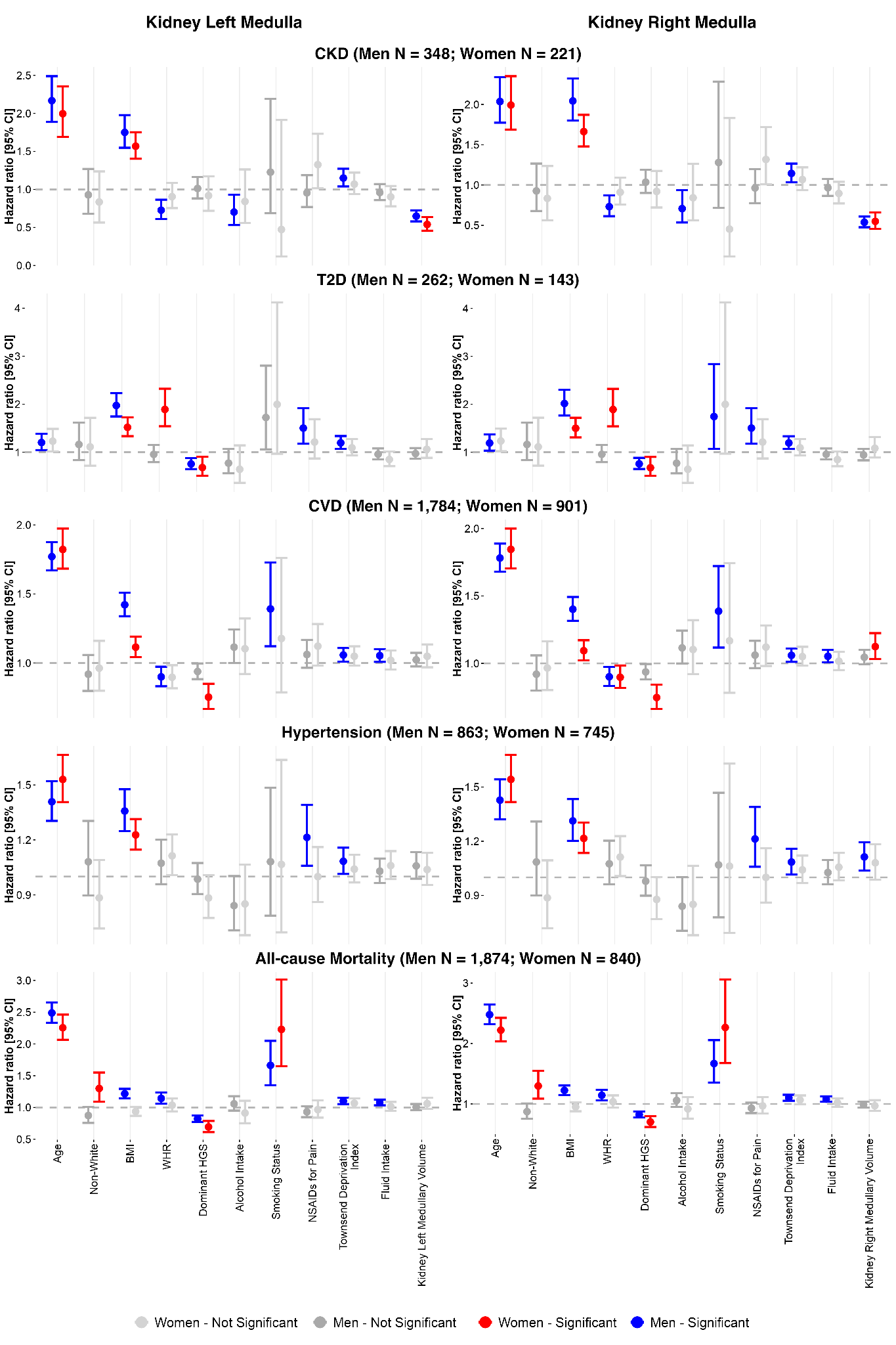

**Figure S23.** Sex-stratified hazard ratios (HR) and 95% confidence intervals (CI) for incident CKD, T2D, CVD, hypertension, and all-cause mortality associated with left and right kidney medulla volume, from Cox proportional hazards models adjusted for age, ethnicity, BMI, WHR, dominant hand grip strength, alcohol intake, smoking status, NSAID use, Townsend deprivation index, and fluid intake, fitted separately in men and women. Significant associations (p < 0.05) are shown in blue (men) and red (women); non-significant associations are shown in grey.

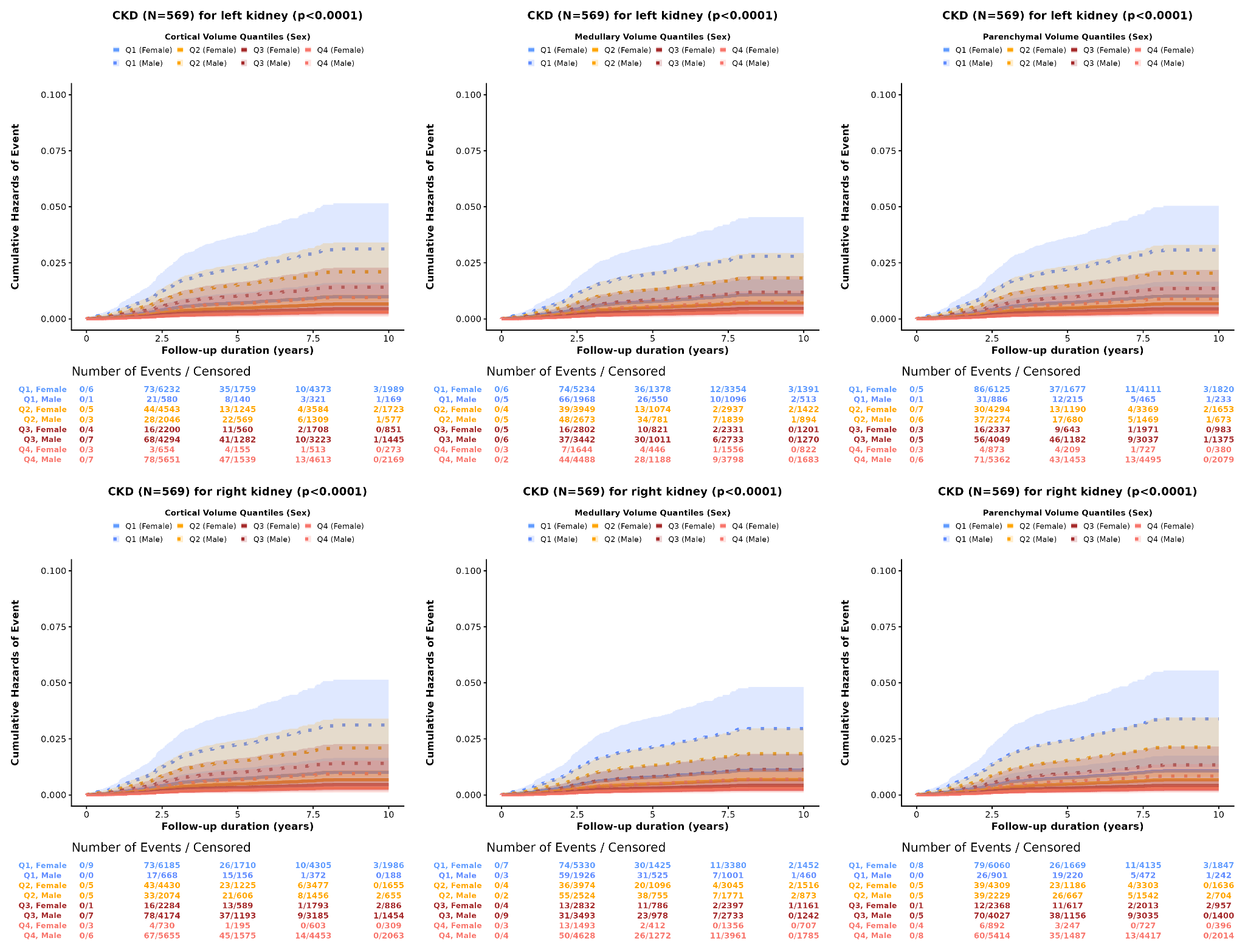

**Figure S24.** Cumulative hazard of incident chronic kidney disease (CKD) (N = 569) over up to 10 years of follow-up, stratified by sex and by quartile (Q1-Q4) of kidney cortical, medullary, and parenchymal volume, for the left and right kidney. Shaded bands show 95% confidence intervals; the number of events and censored participants at each follow-up interval is given below each panel.

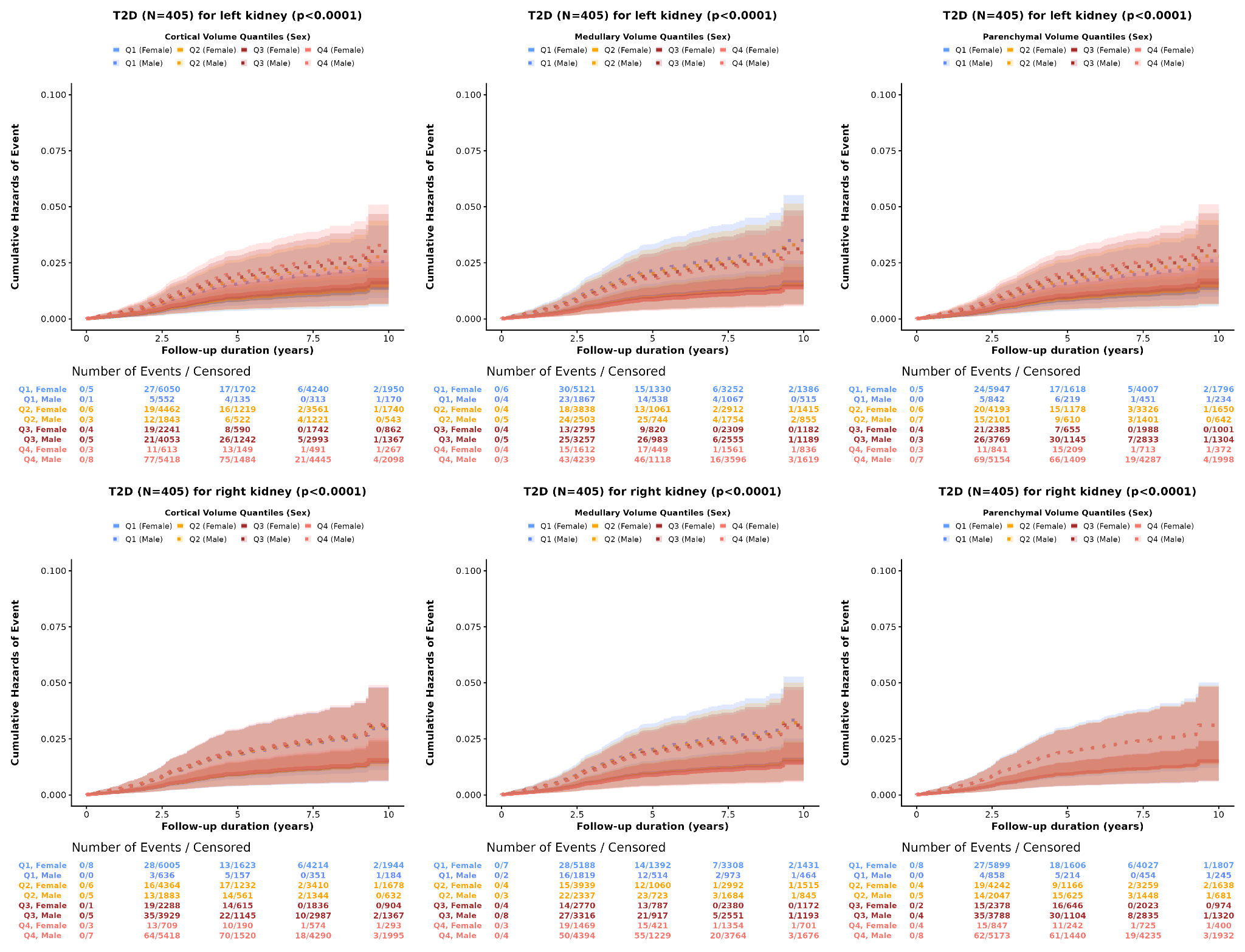

**Figure S25.** Cumulative hazard of incident type-2 diabetes (T2D) (N = 405) over up to 10 years of follow-up, stratified by sex and by quartile (Q1-Q4) of kidney cortical, medullary, and parenchymal volume, for the left and right kidney. Shaded bands show 95% confidence intervals; the number of events and censored participants at each follow-up interval is given below each panel.

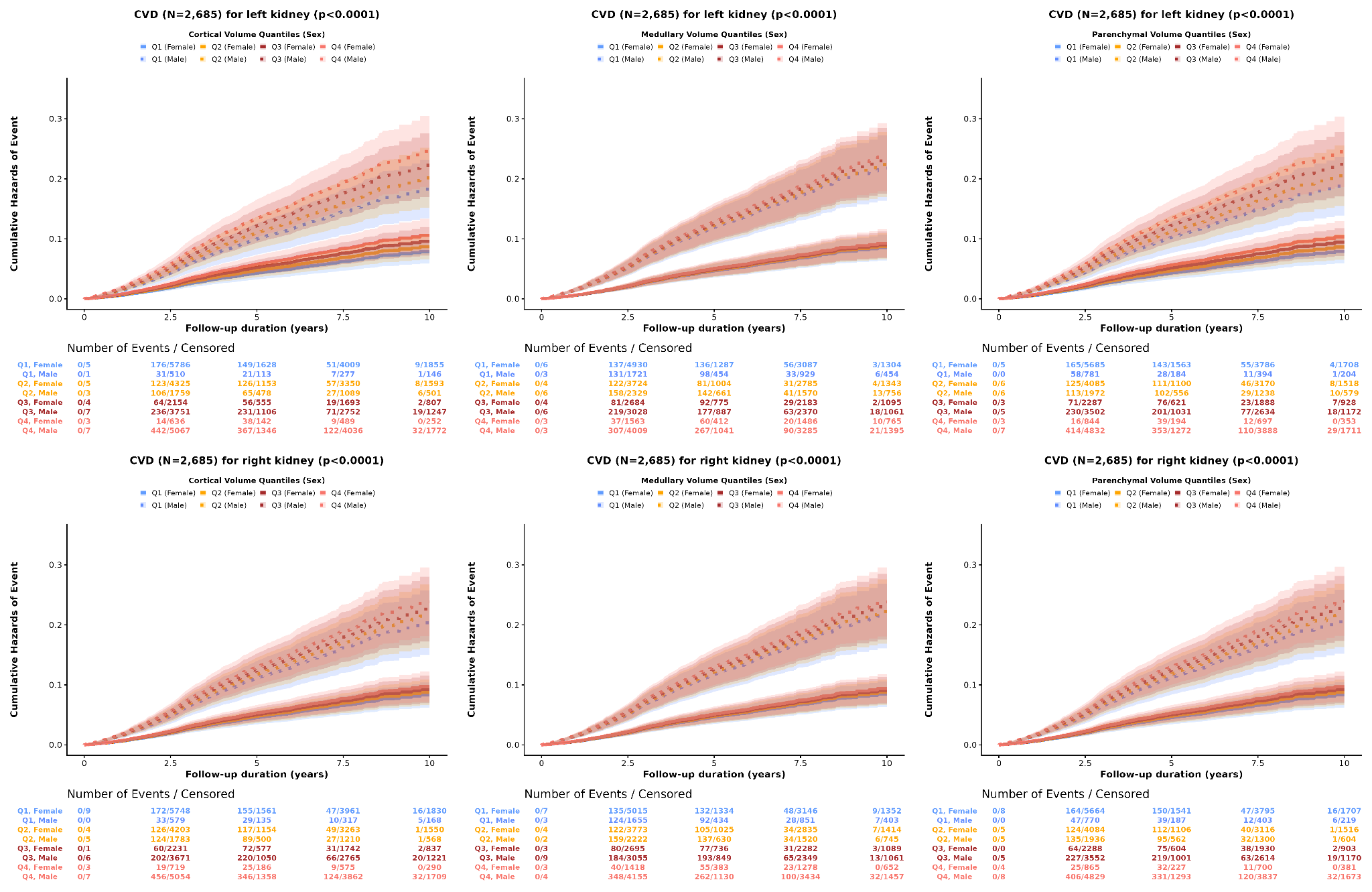

**Figure S26.** Cumulative hazard of incident cardiovascular disease (CVD) (N = 2,685) over up to 10 years of follow-up, stratified by sex and by quartile (Q1-Q4) of kidney cortical, medullary, and parenchymal volume, for the left and right kidney. Shaded bands show 95% confidence intervals; the number of events and censored participants at each follow-up interval is given below each panel.

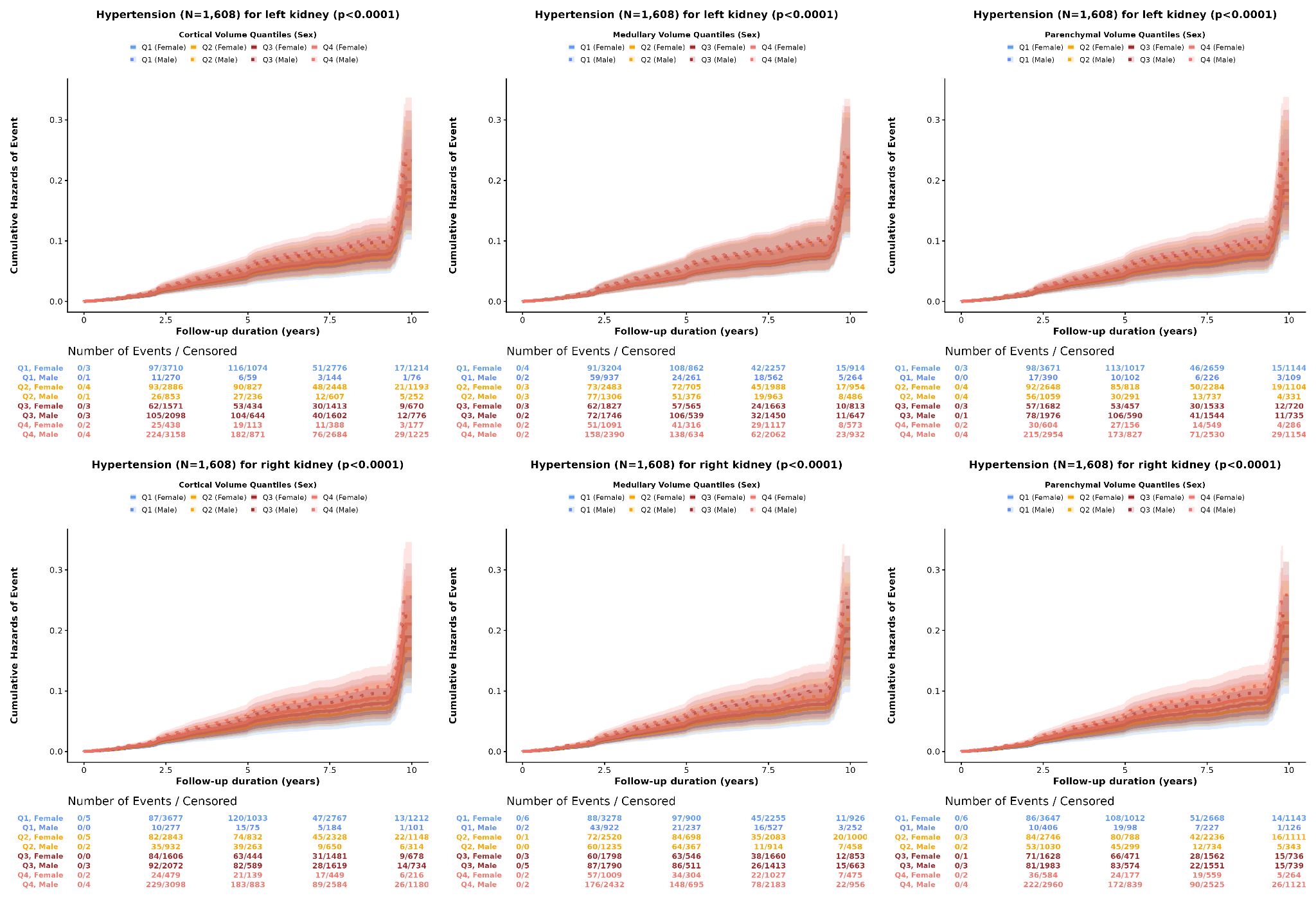

**Figure S27.** Cumulative hazard of incident hypertension (N = 1,608) over up to 10 years of follow-up, stratified by sex and by quartile (Q1-Q4) of kidney cortical, medullary, and parenchymal volume, for the left and right kidney. Shaded bands show 95% confidence intervals; the number of events and censored participants at each follow-up interval is given below each panel.

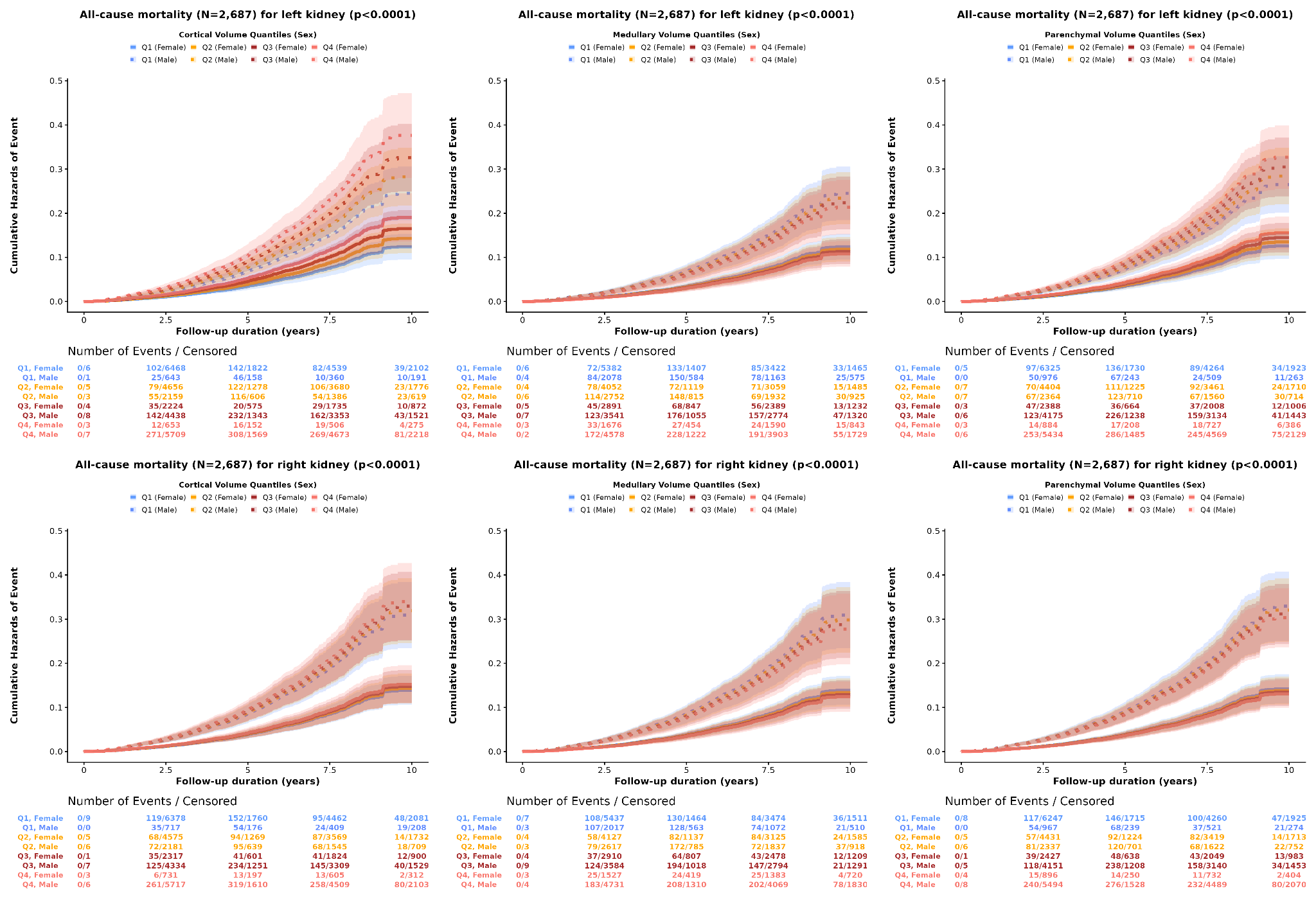

**Figure S28.** Cumulative hazard of incident all-cause mortality (N = 2,687) over up to 10 years of follow-up, stratified by sex and by quartile (Q1-Q4) of kidney cortical, medullary, and parenchymal volume, for the left and right kidney. Shaded bands show 95% confidence intervals; the number of events and censored participants at each follow-up interval is given below each panel.

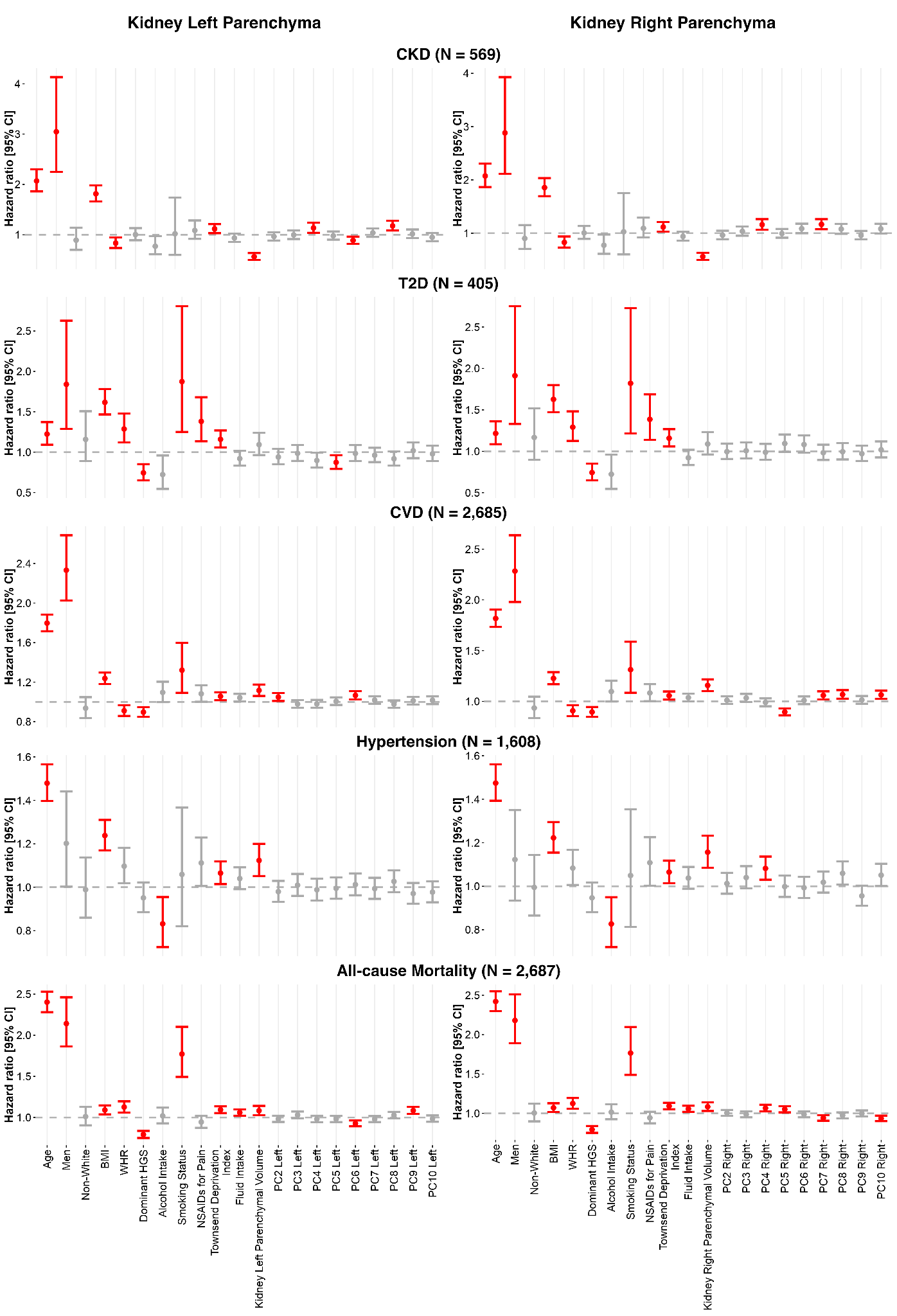

**Figure S29.** Hazard ratios (HR) and 95% confidence intervals (CI) for incident CKD, T2D, CVD, hypertension, and all-cause mortality associated with left and right kidney parenchyma, from Cox proportional hazards models adjusted for age, sex, ethnicity, BMI, WHR, dominant hand grip strength, alcohol intake, smoking status, NSAID use, Townsend deprivation index, fluid intake, kidney parenchymal volume, and the first ten thickness principal component (PC) scores. Significant associations (p < 0.05) are shown in red and non-significant associations in grey.

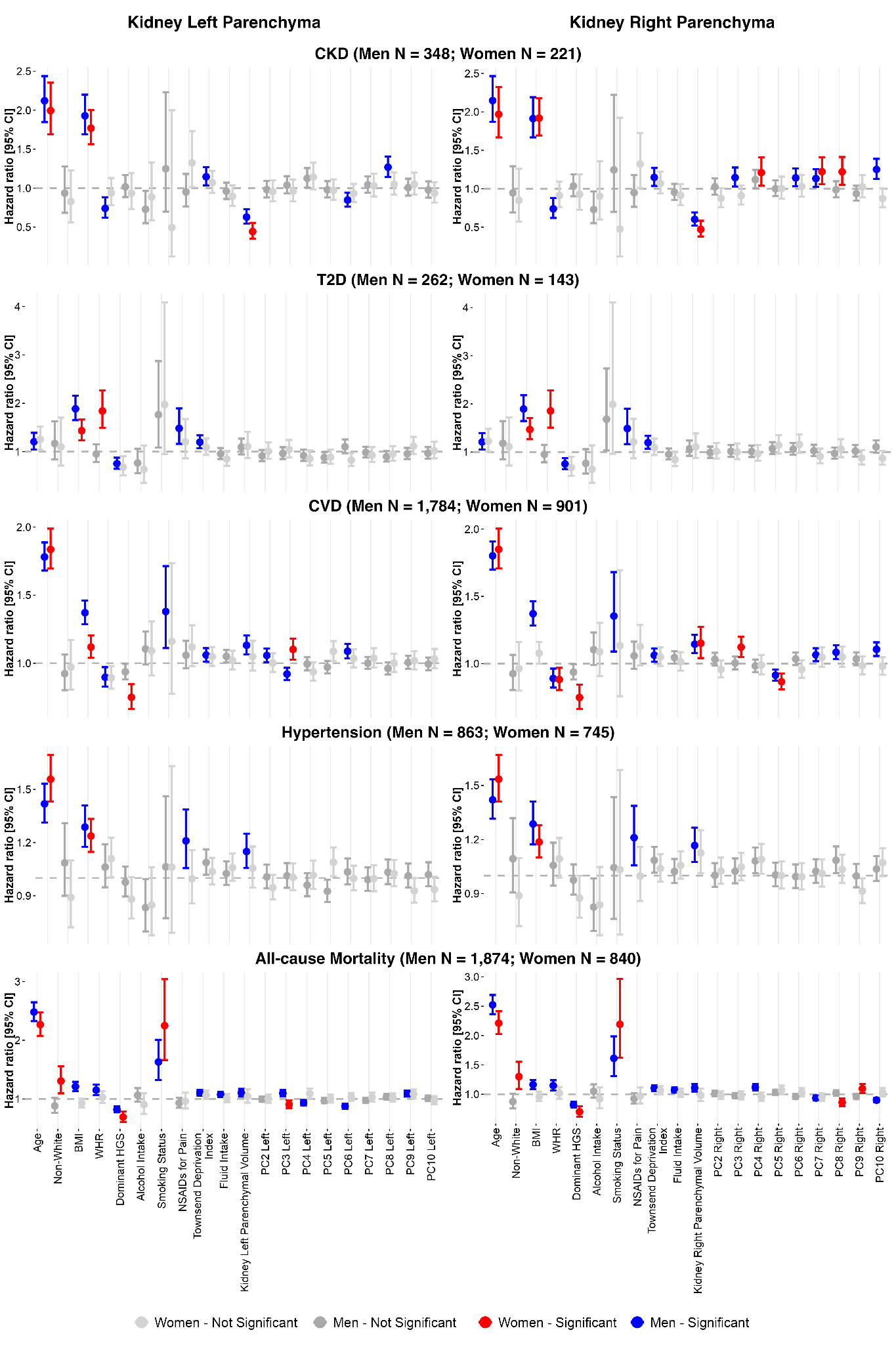
**Figure S30.** Sex-stratified hazard ratios (HR) and 95% confidence intervals (CI) for incident CKD, T2D, CVD, hypertension, and all-cause mortality associated with left and right kidney parenchyma, from Cox proportional hazards models as in Supplementary Figure S29, fitted separately in men and women. Significant associations (p < 0.05) are shown in blue (men) and red (women); non-significant associations are shown in grey.

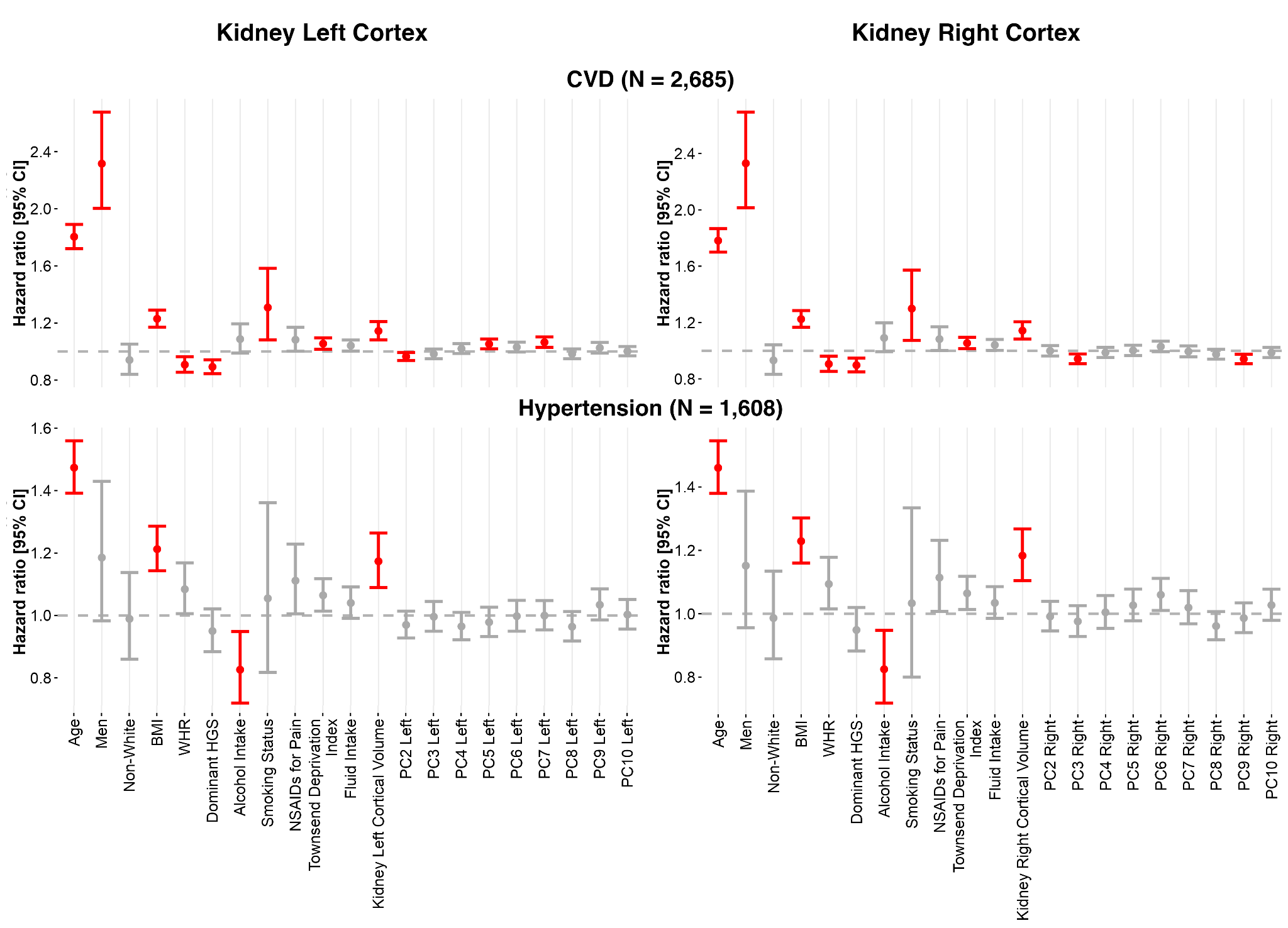

**Figure S31.** Hazard ratios (HR) and 95% confidence intervals (CI) for incident cardiovascular disease (CVD) and hypertension associated with left and right kidney cortex, from Cox proportional hazards models adjusted for age, sex, ethnicity, BMI, WHR, dominant hand grip strength, alcohol intake, smoking status, NSAID use, Townsend deprivation index, fluid intake, kidney cortical volume, and the first ten thickness principal component (PC) scores. Significant associations (p < 0.05) are shown in red and non-significant associations in grey.

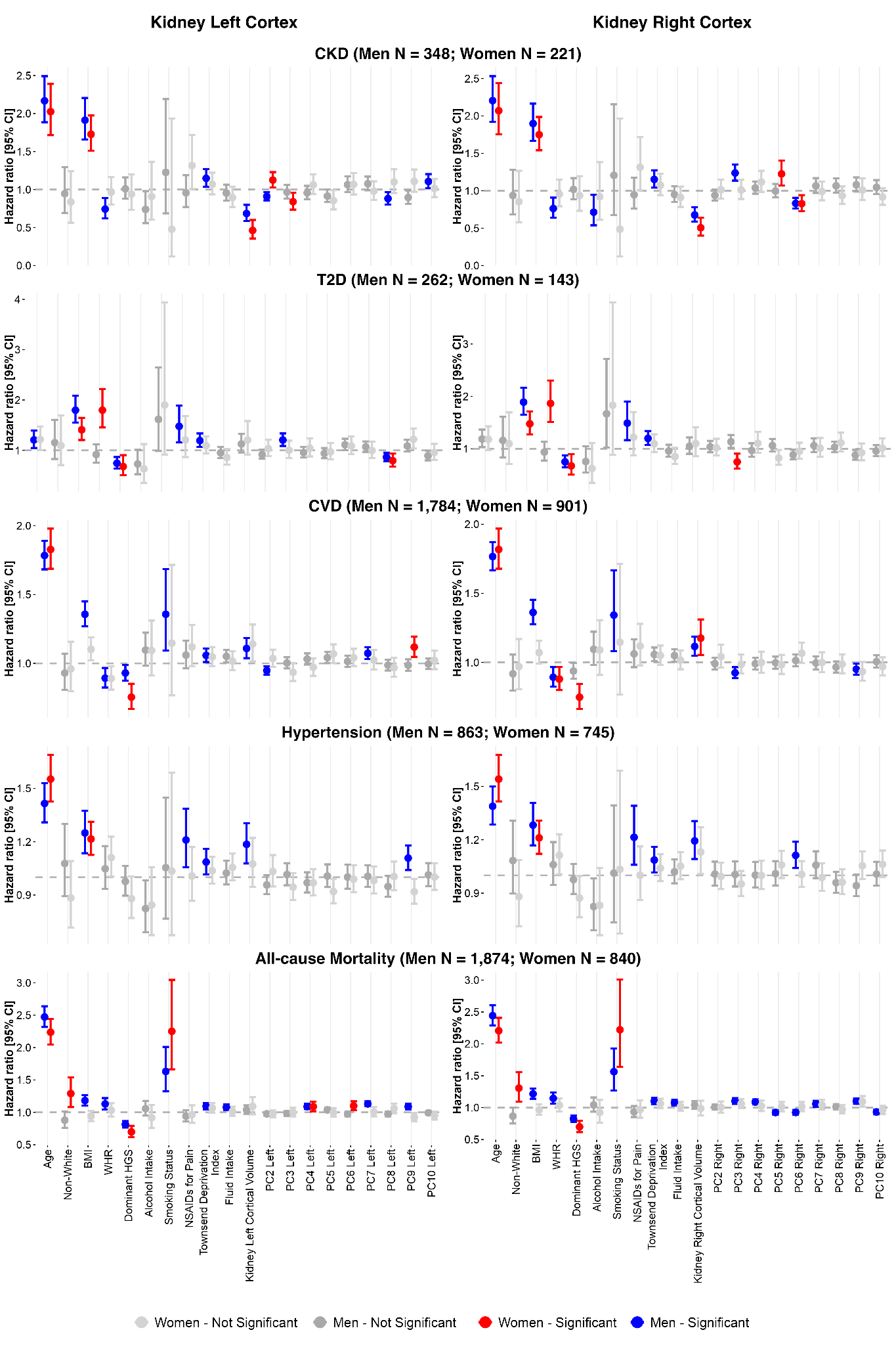

**Figure S32.** Sex-stratified hazard ratios (HR) and 95% confidence intervals (CI) for incident CKD, T2D, CVD, hypertension, and all-cause mortality associated with left and right kidney cortex, from Cox proportional hazards models adjusted for age, ethnicity, BMI, WHR, dominant hand grip strength, alcohol intake, smoking status, NSAID use, Townsend deprivation index, fluid intake, kidney cortical volume, and the first ten thickness principal component (PC) scores, fitted separately in men and women. Significant associations (p < 0.05) are shown in blue (men) and red (women); non-significant associations are shown in grey.

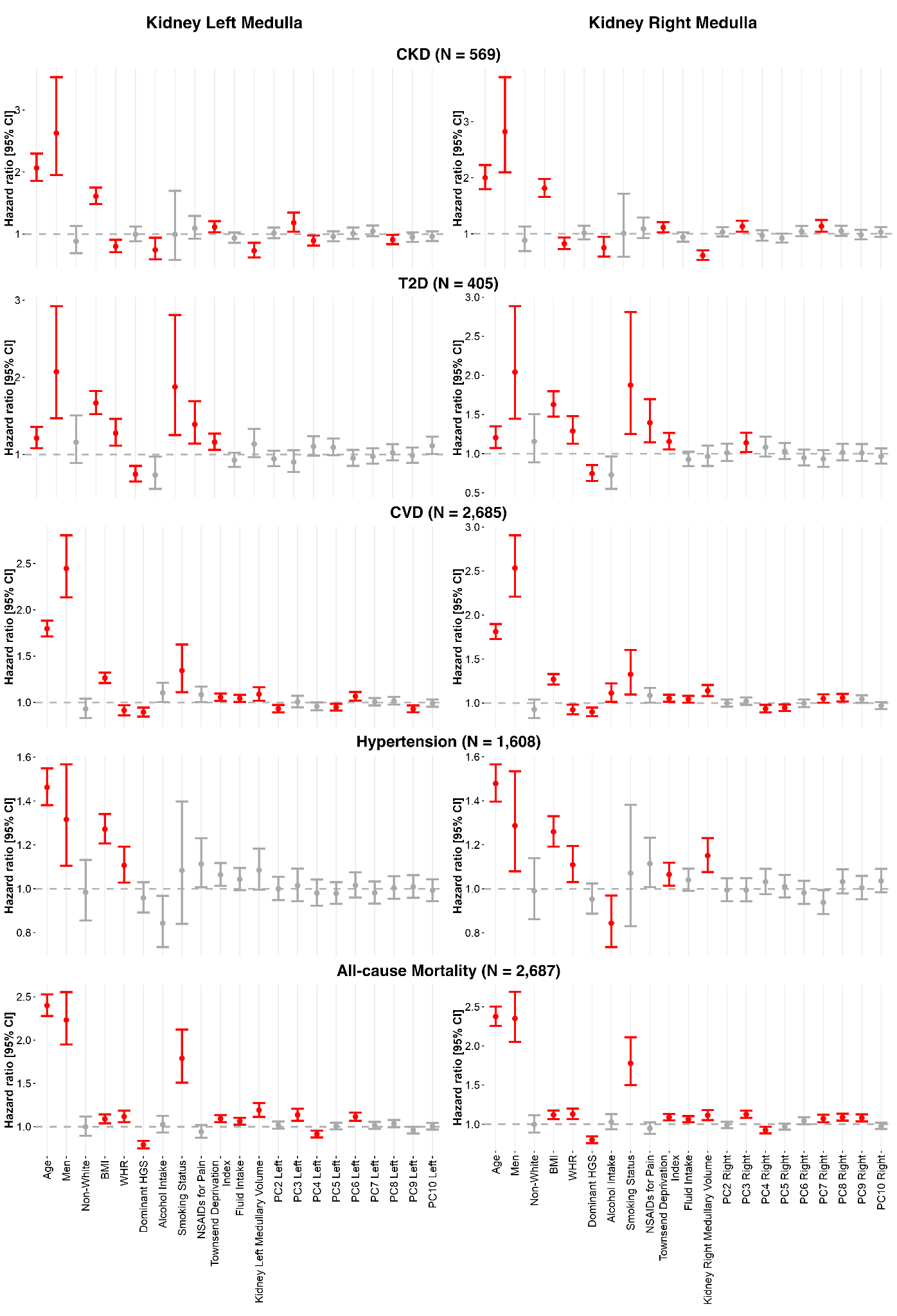

**Figure S33.** Hazard ratios (HR) and 95% confidence intervals (CI) for incident CKD, T2D, CVD, hypertension, and all-cause mortality associated with left and right kidney medulla, from Cox proportional hazards models adjusted for age, sex, ethnicity, BMI, WHR, dominant hand grip strength, alcohol intake, smoking status, NSAID use, Townsend deprivation index, fluid intake, kidney medullary volume, and the first ten thickness principal component (PC) scores. Significant associations (p < 0.05) are shown in red and non-significant associations in grey.

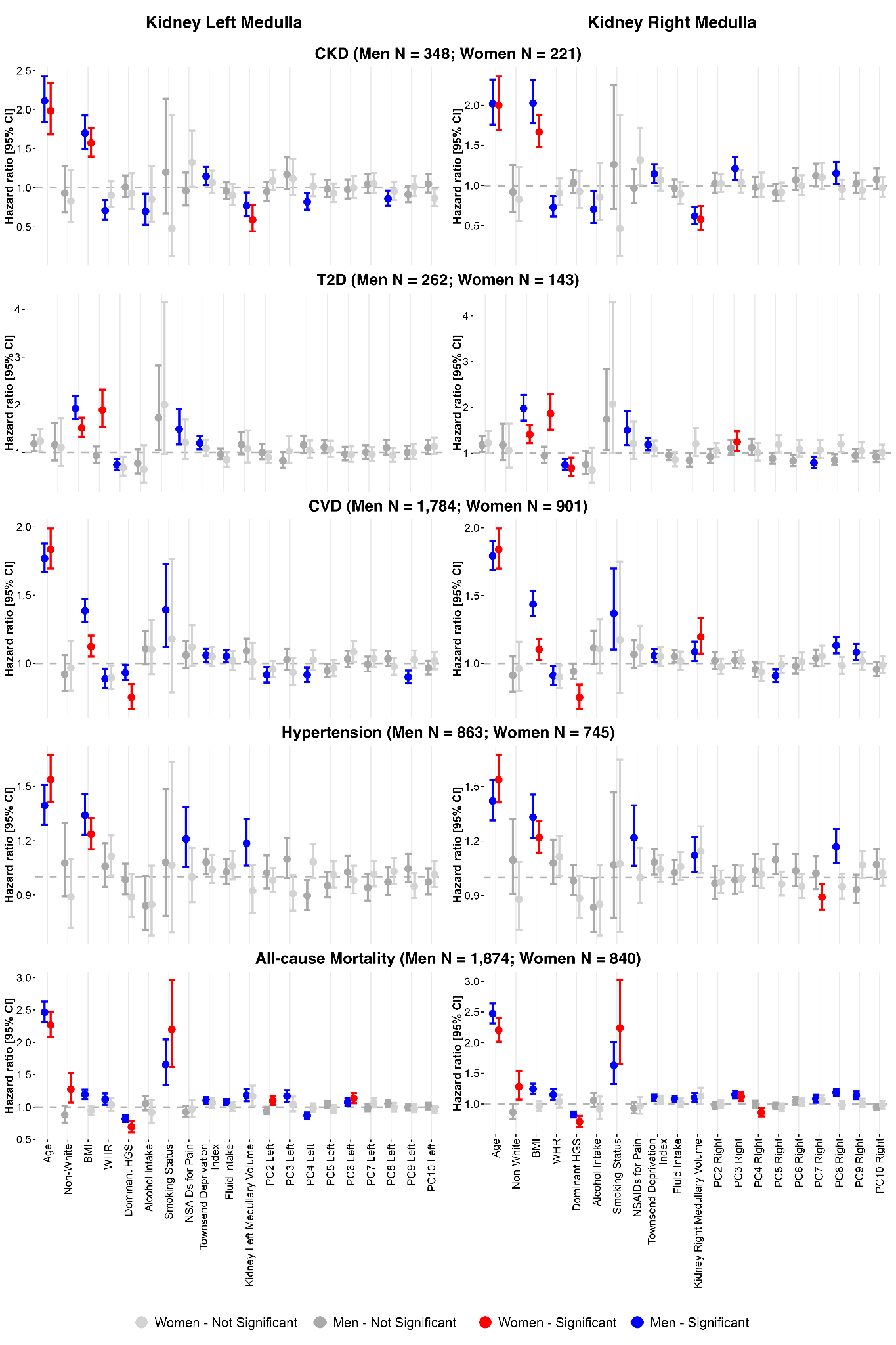

**Figure S34.** Sex-stratified hazard ratios (HR) and 95% confidence intervals (CI) for incident CKD, T2D, CVD, hypertension, and all-cause mortality associated with left and right kidney medulla, from Cox proportional hazards models adjusted for age, ethnicity, BMI, WHR, dominant hand grip strength, alcohol intake, smoking status, NSAID use, Townsend deprivation index, fluid intake, kidney medullary volume, and the first ten thickness principal component (PC) scores, fitted separately in men and women. Significant associations (p < 0.05) are shown in blue (men) and red (women); non-significant associations are shown in grey.

Supplementary Videos

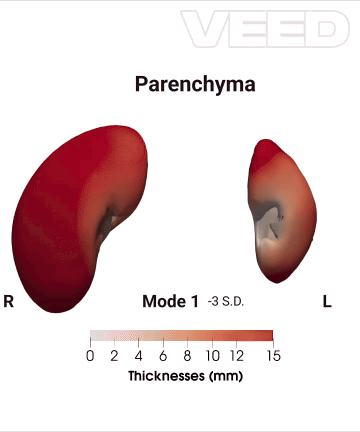

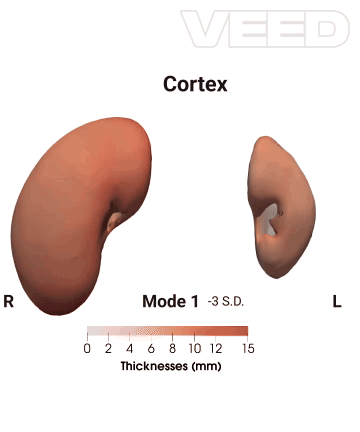

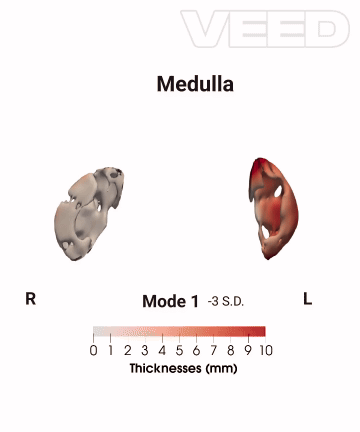

**Video S1.** The first 10 modes of shape variation for the kidney parenchyma, cortex and medullary thicknesses, showing anterior views for both left and right kidney on the full cohort (N=70,270). The mean shape and the shape at the +/- 3 standard deviations are displayed for each mode, showing the kidney thickness change in mm. For visualisation purposes, SPCA was applied jointly to two complementary representations of shape: the 3D coordinates, capturing physical shape variation, and the thicknesses, capturing computational shape differences relative to a reference surface. Together, these two components allow the modes to reflect both the physical deformation of the structure and the surface-level morphological variation.
